# Distinguishing different psychiatric disorders using DDx-PRS

**DOI:** 10.1101/2024.02.02.24302228

**Authors:** Wouter J. Peyrot, Georgia Panagiotaropoulou, Loes M. Olde Loohuis, Mark J. Adams, Swapnil Awasthi, Tian Ge, Andrew M. McIntosh, Brittany L. Mitchell, Niamh Mullins, Kevin S O’Connell, Brenda W.J.H. Penninx, Danielle Posthuma, Stephan Ripke, Douglas M. Ruderfer, Emil Uffelmann, Bjarni J. Vilhjalmsson, Zhihong Zhu, Schizophrenia Working Group of the Psychiatric Genomics Consortium, Bipolar Disorder Working Group of the Psychiatric Genomics Consortium, Major Depressive Disorder Working Group of the Psychiatric Genomics Consortium, Jordan W. Smoller, Alkes L. Price

## Abstract

Despite great progress on methods for case-control polygenic prediction (e.g. schizophrenia vs. control), there remains an unmet need for a method that genetically distinguishes clinically related disorders (e.g. schizophrenia (SCZ) vs. bipolar disorder (BIP) vs. depression (MDD) vs. control); such a method could have important clinical value, especially at disorder onset when differential diagnosis can be challenging. Here, we introduce a method, Differential Diagnosis-Polygenic Risk Score (DDx-PRS), that jointly estimates posterior probabilities of each possible diagnostic category (e.g. SCZ=50%, BIP=25%, MDD=15%, control=10%) by modeling variance/covariance structure across disorders, leveraging case-control polygenic risk scores (PRS) for each disorder (computed using existing methods) and prior clinical probabilities for each diagnostic category. DDx-PRS uses only summary-level training data and does not use tuning data, facilitating implementation in clinical settings. In simulations, DDx-PRS was well-calibrated (whereas a simpler approach that analyzes each disorder marginally was poorly calibrated), and effective in distinguishing each diagnostic category vs. the rest. We then applied DDx-PRS to Psychiatric Genomics Consortium SCZ/BIP/MDD/control data, including summary-level training data from 3 case-control GWAS (*N*=41,917-173,140 cases; total *N*=1,048,683) and held-out test data from different cohorts with equal numbers of each diagnostic category (total *N*=11,460). DDx-PRS was well-calibrated and well-powered relative to these training sample sizes, attaining AUCs of 0.66 for SCZ vs. rest, 0.64 for BIP vs. rest, 0.59 for MDD vs. rest, and 0.68 for control vs. rest. DDx-PRS produced comparable results to methods that leverage tuning data, confirming that DDx-PRS is an effective method. True diagnosis probabilities in top deciles of predicted diagnosis probabilities were considerably larger than prior baseline probabilities, particularly in projections to larger training sample sizes, implying considerable potential for clinical utility under certain circumstances. In conclusion, DDx-PRS is an effective method for distinguishing clinically related disorders.

## Introduction

Great progress has been made in methods for case-control polygenic prediction (e.g. schizophrenia vs. control)^1–8^. However, in clinical practice the most pressing challenge is often to distinguish related disorders from each other as well as controls (e.g. schizophrenia (SCZ) vs. bipolar disorder (BIP) vs. depression (MDD) vs. control), especially at disorder onset when differential diagnosis can be challenging. The importance of maximizing the utility of genetic predictions for clinical practice is well-recognized^9^, but progress on genetically distinguishing related disorders has been limited; a recent study proposed a method to distinguish inflammatory-arthritis causing diseases using genome-wide significant variants^10^, but this approach is unlikely to perform well for highly polygenic disorders such as psychiatric disorders, for which the contribution of genome-wide significant variants is limited^1,2,8,11,12^.

Here, we introduce a new method, Differential Diagnosis-Polygenic Risk Score (DDx-PRS), to distinguish related disorders from each other and from controls. DDx-PRS models the variance/covariance structure of disorder liabilities^13^ and polygenic risk scores (PRS) across disorders, leveraging case-control PRS computed using all genome-wide variants^3,5–7^. DDx-PRS uses prior disorder probabilities (which can be flexibly specified by a clinician) and applies Bayes’ Theorem to compute posterior probabilities of each diagnostic category (e.g. SCZ=50%, BIP=25%, MDD=15%, control=10%). We apply DDx-PRS to predict SCZ vs. BIP vs. MDD vs. control in simulated data and in empirical data from the Psychiatric Genomics Consortium^8,14–16^. We also apply DDx-PRS to simulated data at larger training sample sizes to obtain projections of future clinical utility.

## Results

### Overview of methods

DDx-PRS distinguishes cases from clinically related disorders (e.g. SCZ vs. BIP vs. MDD vs. control), estimating posterior probabilities of each diagnostic category using case-control GWAS summary statistics for each disorder (as training data) and prior clinical probabilities (e.g. a unform prior of 25% for each diagnostic category). DDx-PRS consists of 4 steps: (1) estimate prior probabilities of each possible configuration of liabilities; (2) compute case-control PRS for each disorder; (3) estimate analytical variances and covariances across disorders of liabilities and case-control PRS (overall and for each configuration of liabilities); and (4) estimate posterior probabilities for each test sample. Each step is summarized below; details are provided in the Methods section. We have publicly released open-source software implementing DDx-PRS (see Code Availability). DDx-PRS has low computational cost, excluding the time required to compute case-control PRS (see Methods).

In step 1, DDx-PRS relies on the fact that, for *n* disorders, there exist 2*^n^* possible configurations of liabilities (above or below the liability threshold for each disorder^13^), e.g. 8 possible configurations for 3 disorders; the mapping from configurations of liabilities to diagnostic categories is informed by the PGC classifications used in ref. ^8,14–16^ (Table 1). Given prior clinical probabilities of each diagnostic category, DDx-PRS estimates prior probabilities of each configuration of liabilities by assuming random sampling of liabilities within each diagnostic category. We note that the prior clinical probability for help-seeking individuals will typically be much larger than the population prevalence. For simplicity, we assume a clinical prior probability of 25% for each of SCZ, BIP, MDD and control for the main analyses presented in this paper; however, in application of DDx-PRS, the prior can be specified flexibly by the physician based on e.g. psychiatric examination, questionnaires and/or disorder prevalence in a specific clinical setting.

**Table 1.**
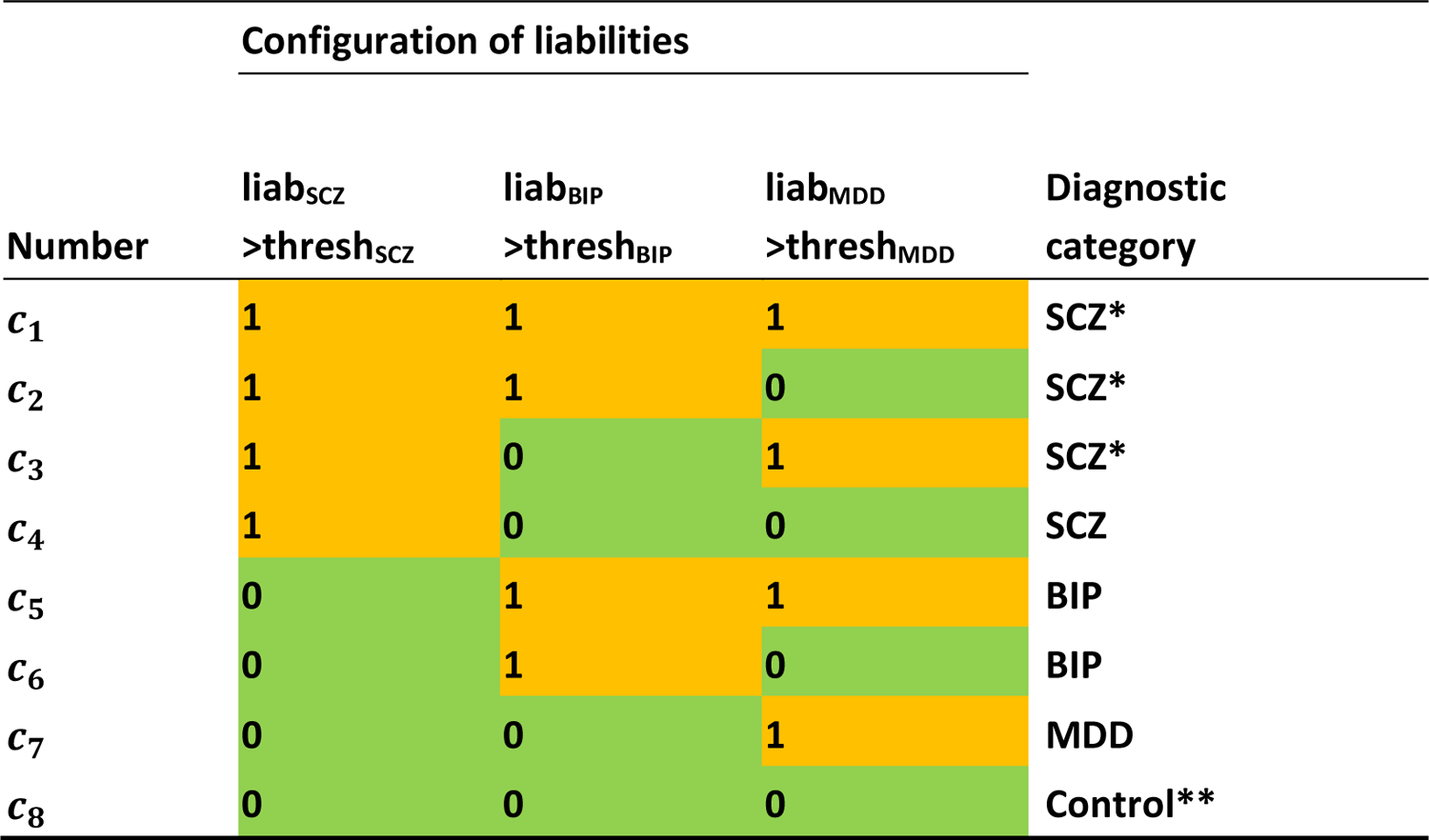
Overview of configurations of liabilities and disorder classification. For schizophrenia (SCZ), bipolar disorder (BIP) and major depressive disorder (MDD), we list the eight configurations of liabilities (i.e. liability (liab) above or below the liability threshold (thresh) for each disorder) and the corresponding diagnostic category. *These would likely be diagnosed as schizoaffective disorder following DSM-5 classification criteria (or as comorbid SCZ and MDD for configuration *c*_3_), but are labelled here as SCZ in line with ref.^8^. **Controls are unaffected by SCZ, BIP and MDD but could potentially have other disorders.

In step 2, DDx-PRS computes case-control PRS for each disorder (on the observed scale) using an existing method (in this paper, we use PRS-CS^5^) and transforms the PRS to their respective liability scales^17^. Case-control PRS are computed for both the test samples and samples from an external reference panel such as 1000 Genomes^18^; the external reference panel does not include phenotypes, and as such does not constitute a tuning data set.

In step 3, DDx-PRS estimates (i) overall variances and covariances of liabilities for each disorder using cross-trait LDSC^19^, (ii) overall variances and covariances of liability-scale case-control PRS for each disorder using the external reference panel (e.g. 1000 Genomes^18^)—in particular, this accounts for sample overlap across controls for different disorders in GWAS training data increasing correlation between the case-control PRS^19,20^, and (iii) overall covariances between liabilities and case-control PRS, based on analytical computations. DDx-PRS then estimates truncated multivariate normal probability densities of liabilities and case-control PRS for each configuration of liabilities using analytical derivations.

In step 4, DDx-PRS applies Bayes’ Theorem to estimate posterior probabilities of each configuration of liabilities for each test sample, conditional on their case-control PRS. Letting *c*_*j*_ denote configuration *j* and 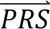_*i*_ denote the case-control PRS for each disorder of individual *i*, these posterior probabilities can be expressed as

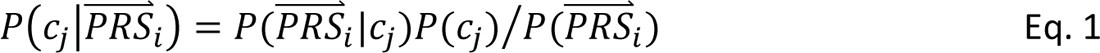

where *P*(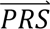_*i*_|*c*_*j*_) denotes the probability density of 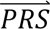_*i*_ given configuration *c*_*j*_ (computed from the output of step 3); *P*(*c*_*j*_) denotes the prior probability of configuration *c*_*j*_ (specified in step 1); and *P*(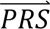*S* _*i*_) denotes the probability density of 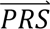_*i*_ summed across all configurations weighted by the prior probabilities, i.e. *P*^(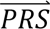^*S* _*i*_) = ∑_*j*′_ *P*^(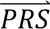^*S* _*i*_ |*c*_*j*′_)*P*(*c*_*j*′_). The posterior probabilities of each configuration of liabilities are subsequently combined into posterior probabilities of each diagnostic category using the information in Table 1.

We compare the results of DDx-PRS to the results of a simpler method, Marginal-PRS (Table 2). Briefly, Marginal-PRS consists of 3 steps: (1) compute the marginal disorder probabilities *P*(*SCZ*), *P*(*BIP*) and *P*(*MDD*) using univariate densities for each disorder separately^17^; (2) compute the control probability as *P*(*control*) = (1 − *P*(*SCZ*))(1 − *P*(*BIP*))(1 − *P*(*MDD*)); and (3) rescale these four probabilities to add up to 1. We note two important limitations of Marginal-PRS compared to DDx-PRS. First, Marginal-PRS does not model the covariance structure between the respective case-control PRS. Second, when computing marginal disorder probabilities, Marginal-PRS does not account for the other disorders (e.g., when computing *P*(*SCZ*), Marginal-PRS incorrectly assumes that non-SCZ individuals are all controls).

**Table 2.**
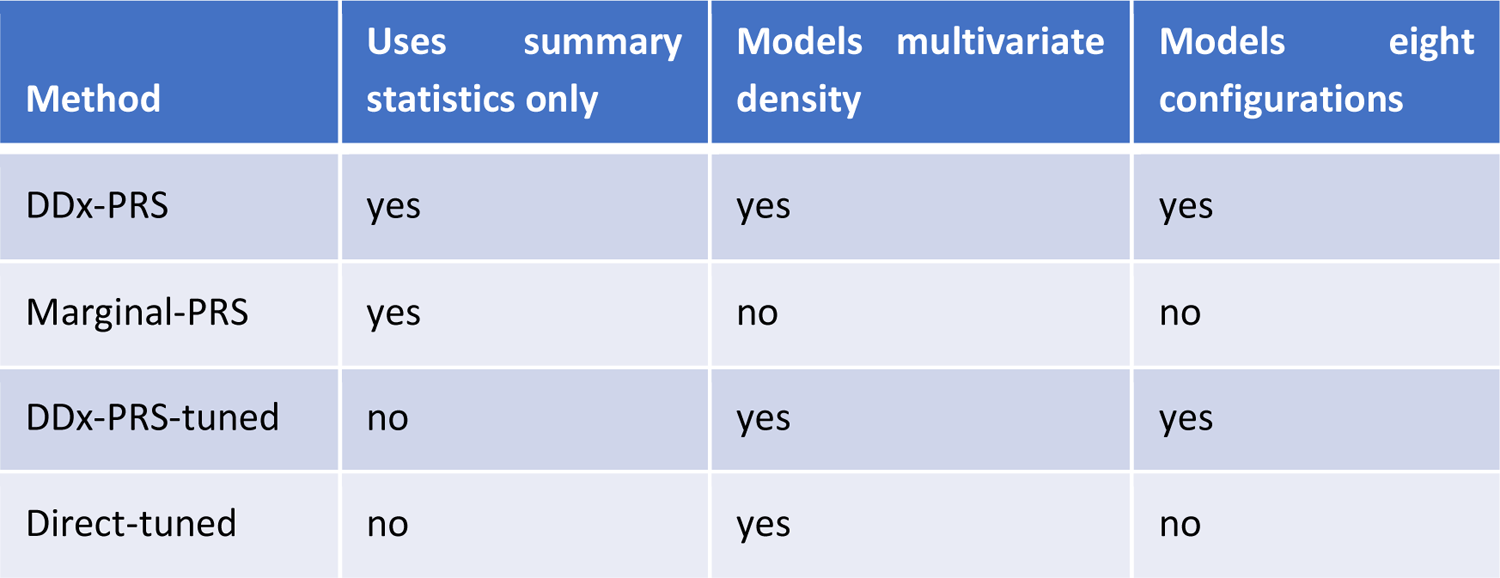
List of methods for genetically distinguishing different disorders. We list the methods for genetically distinguishing different disorders evaluated in this paper, and indicate whether they use summary statistics only (as opposed to summary statistics plus tuning data), whether they model multivariate density (across four diagnostic categories or eight configurations from Table 1), and whether they specifically model the all configurations from Table 1.

| Method | Uses summary<br>statistics only | Models multivariate<br>density | Models eight<br>configurations |
| --- | --- | --- | --- |
| DDx-PRS | yes | yes | yes |
| Marginal-PRS | yes | no | no |
| DDx-PRS-tuned | no | yes | yes |
| Direct-tuned | no | yes | no |

DDx-PRS and Marginal-PRS do not require additional tuning data to train parameters connecting case-control PRS to probabilities of each diagnostic category. Although methods that do not require tuning data are more practical, for completeness we also considered two methods that do require tuning data: DDx-PRS-tuned and Direct-tuned (Table 2 and Methods). DDx-PRS-tuned uses a tuning dataset to estimate the multivariate probability densities of 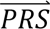in step 2 above more directly (while still modelling the eight configurations *c*_*j*_). Direct-tuned applies Bayes’ Theorem as in Eq. 1 to the four diagnostic categories directly (i.e. using four diagnostic categories instead of the eight configurations in Table 1), estimating the multivariate probability densities of 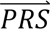in these four categories directly from the tuning data. In our main analyses, we use cross-cohort tuning data (we also investigated the performance of these methods using within-cohort tuning data, although we caution that it is much less likely that this type of data - matched with respect to sampling location, ancestry, and genotyping platform - will be available in practice).

We assessed the performance of each method in classifying each diagnostic category vs. the rest (SCZ vs. rest, BIP vs. rest, MDD vs. rest, and control vs. rest). Calibration was assessed using the Integrated Calibration Index (ICI)^21^, defined as the weighted absolute difference between the predicted probability and the true probability. Briefly, the ICI is computed by estimating the true probabilities compared to the predicted probability using a locally estimated scatterplot smoothing (loess)-based smoothing function, and subsequently assessing the average absolute difference between the predicted and true probabilities, weighted by the density of predicted probabilities; it thus reflects the average error of the predicted probabilities (ICI=0 reflects perfect calibration). Prediction accuracy was assessed using the area under the ROC curve (AUC)^22^.

We evaluated the performance of each method using data from the SCZ, BIP and MDD working groups of the Psychiatric Genomics Consortium (PGC; see Data Availability). The full PGC data that we analyzed consisted of the European-ancestry case-control GWAS summary statistics for SCZ^8^ (*N*=53,386 cases and 77,258 controls), BIP^14^ (*N*=41,917 cases and 371,549 controls), and MDD^15,16^ (*N*=173,140 cases and 331,433 controls) (Methods and Table S1). The test data consisted of merged PGC case-control cohorts comprising 11,460 individuals of European ancestry (2,865 of each of SCZ, BIP, MDD, control), subdivided into 4 test cohorts within which SCZ, BIP, MDD, and control samples were matched with respect to country and genotyping platform (Methods and Table S2). For each disorder-specific PGC case-control cohort that was included in one of the 4 merged test cohorts, we constructed corresponding training data sets (used to compute case-control PRS for each disorder) that excluded that PGC case-control cohort.

### Simulations

We simulated data based on the genetic architectures of SCZ^8^, BIP^14^ and MDD^15,16^, with liability-scale SNP-heritabilities^23^ of 0.24, 0.19 and 0.09, population prevalences of 1%, 2% and 16%, liability-scale case-control PRS *r*^2^ (ref.^24^) of 0.10, 0.09 and 0.04, respectively, and genetic correlations of 0.70 for SCZ-BIP, 0.35 for SCZ-MDD and 0.45 for BIP-MDD (collectively matching our empirical findings; see Methods). We simulated individual-level data for 1,000 SNPs (of which 50% were causal) in training samples whose sample size was selected to attain the specified case-control PRS *r*^2^ and 1,000 test samples for each diagnostic category (SCZ, BIP, MDD, controls). We introduced sample overlap of controls in SCZ, BIP and MDD training data to mimic empirical data, which includes substantial overlap of controls^20^. We simulated a small number of SNPs (*M*) and training samples (*N*) to limit computational cost, while noting that PRS *r*^2^ primarily depends on *M*/*N*, so that simulations at reduced values of both *M* and *N* are appropriate^3^. Because linkage disequilibrium (LD) does not impact cross-disorder architectures conditional on the above parameter values, we simulated genotypes without LD. Case-control PRS were computed using Bpred, which analytically computes posterior mean causal effects sizes under a point-normal prior in the special case of no LD^3^. We simulated individual-level SNP data (instead of directly simulating case-control PRS values in test samples) to also assess the performance of the complexities of transforming case-control PRS values to the liability scale (see Methods). We analyzed 50 simulation replicates for each simulation. We assessed the performance of DDx-PRS and Marginal-PRS (Table 2) in classifying each diagnostic category vs. the rest (SCZ vs. rest, BIP vs. rest, MDD vs. rest, and control vs. rest). Calibration was assessed using the ICI^21^, defined as the weighted absolute difference between the predicted probability and the true probability, and prediction accuracy was assessed using the area under the ROC curve (AUC)^22^.

The calibration of DDx-PRS and Marginal-PRS are reported in Figure 1a and Table S3. DDx-PRS was well-calibrated with a mean ICI (±SE) of 0.024±0.001 (averaged across the four comparisons), whereas Marginal-PRS was substantially less well-calibrated with a mean ICI of 0.055±0.002. Marginal-PRS suffered particularly poor calibration in the comparison of control vs. rest (mean ICI of 0.108±0.006) because Marginal-PRS systematically underestimates the three disorder probabilities and overestimates the control probability, as observed in comparisons of true probability vs. predicted probability for each diagnostic category (Figure 2). In addition to computing the mean ICI across simulation replicates, we computed concatenated ICI for simulated data that was concatenated across simulation replicates (Figure 1a and Table S3). The concatenated ICI (averaged across the four comparisons) was much lower than the mean ICI for DDx-PRS (0.010 vs. 0.024), likely because the concatenated results were less noisy due to 50 times larger test sample size. However, the concatenated ICI was only very slightly smaller than the mean ICI for Marginal-PRS (0.054 vs. 0.055), which we interpret as follows. The ICI reflects prediction error due to both noise and method miscalibration, and the concatenated ICI reduces the noise (most important for DDx-PRS) but not the method miscalibration (most important for Marginal-PRS). The absolute difference between the mean ICI and concatenated ICI is larger for DDx-PRS than for Marginal PRS, because noise for a well-calibrated method always increases the ICI whereas noise for a miscalibrated method can either increase or decrease the ICI.

**Figure 1.**
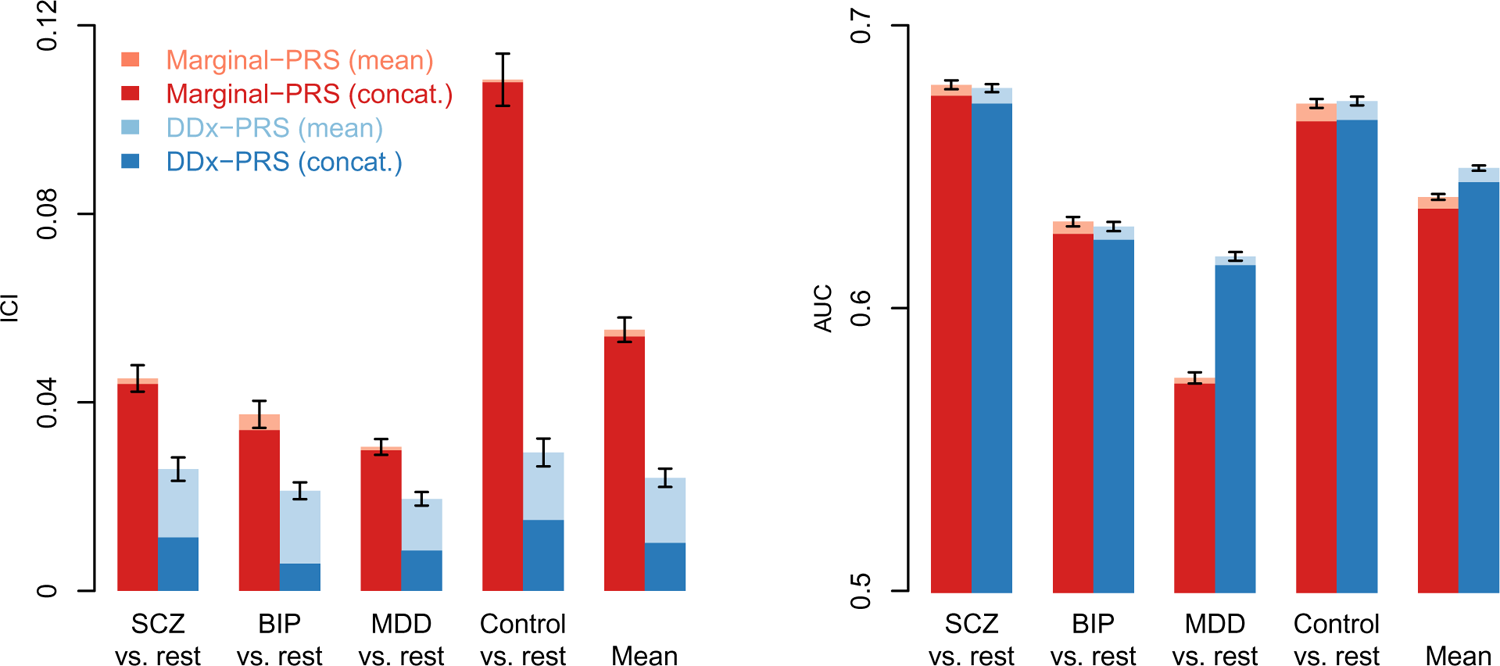
Calibration and accuracy of DDx-PRS and Marginal-PRS in simulations. We report the calibration (Integrated Calibration Index; ICI) and accuracy (area under the ROC curve; AUC) for the comparisons of schizophrenia (SCZ) vs. rest, bipolar disorder (BIP) vs. rest, major depressive disorder (MDD) vs. rest, control vs. rest and the mean across these four comparisons, for two methods: Marginal-PRS (red) and DDx-PRS (blue). Results are based on 50 simulation replicates; mean values are displayed in light red and light blue, and results based on data concatenated across 50 replicates are displayed in dark red and dark blue. Error bars denote standard errors. Numerical results are reported in Table S3.

**Figure 2.**
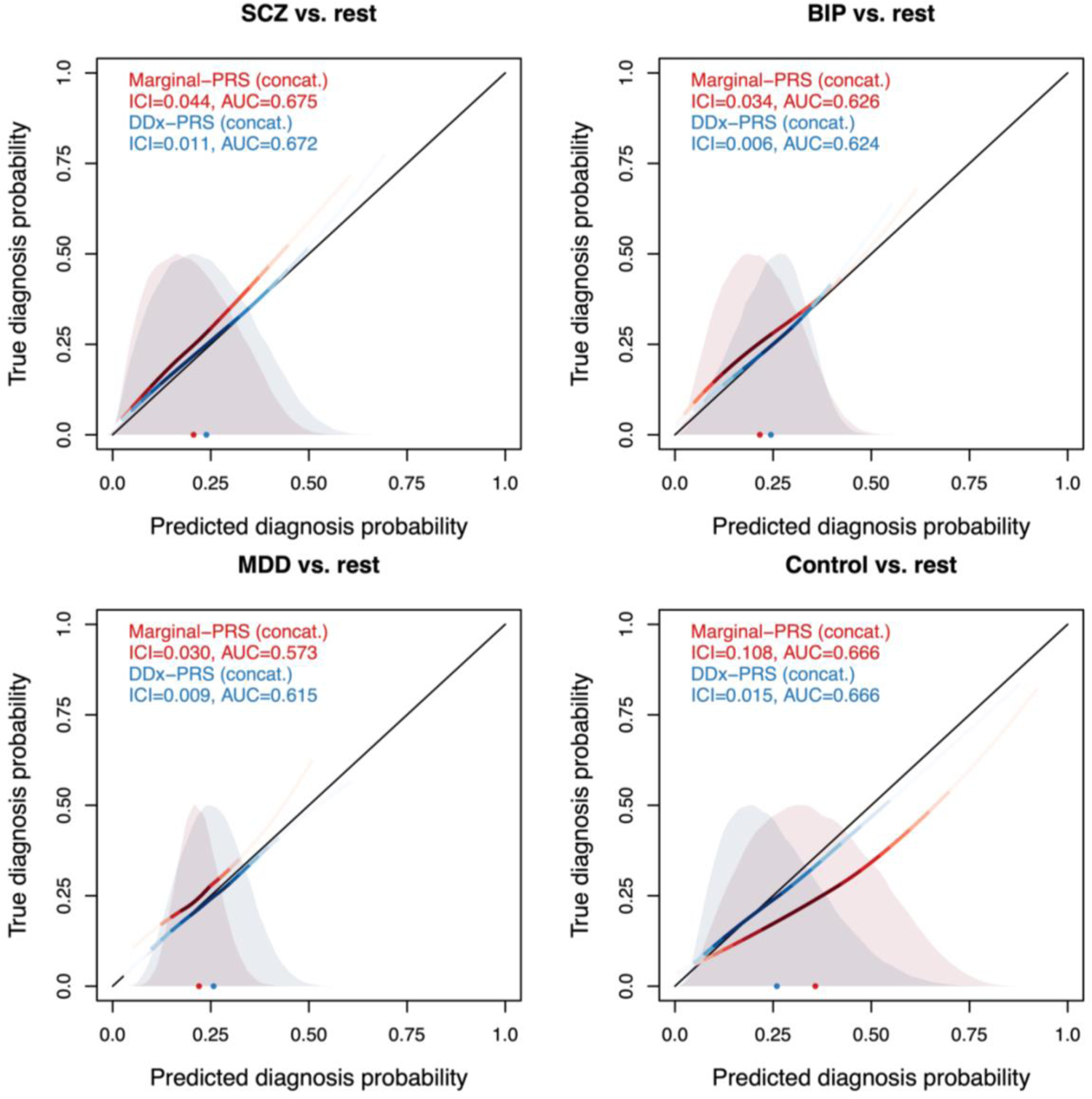
Calibration curves of DDx-PRS and Marginal-PRS in simulations. We report the true diagnosis probability vs. the predicted diagnosis probability for the comparisons of schizophrenia (SCZ) vs. rest, bipolar disorder (BIP) vs. rest, major depressive disorder (MDD) vs. rest, and control vs. rest, for two methods: Marginal-PRS (red) and DDx-PRS (blue). Results are based on concatenated data across 50 simulation replicates. The calibration curves are plotted using a locally estimated scatterplot smoothing (loess)-based smoothing function. The Integrated Calibration Index (ICI) equals the average absolute difference between the calibration curve and the line y=x (plotted in black), weighted by the density of the predicted diagnosis probabilities (displayed in the shaded histograms and in the color-intensity of the calibration curves). AUC, area under the ROC curve.

The prediction accuracy of DDx-PRS and Marginal-PRS are reported in Figure 1b and Table S3. The accuracy of DDx-PRS averaged across the four comparisons (mean AUC of 0.650±0.001) was slightly higher than the accuracy of Marginal-PRS (average AUC of 0.639±0.001). The difference in AUC was most pronounced for the comparison of MDD vs. rest (0.618±0.002 for DDx-PRS vs. 0.575±0.002 for Marginal-PRS) because DDx-PRS explicitly models the other disorders, whereas Marginal-PRS incorrectly assumes that non-MDD individuals are all controls. This has most impact for MDD because the genetic distance(MDD,control) is much smaller than distance(SCZ,control) or distance(BIP,control) due to the higher prevalence of MDD (Figure 1 of ref.^20^). The concatenated AUC (averaged across the four comparisons) were slightly smaller than the mean AUC (0.645 vs. 0.650 for DDx-PRS and 0.635 vs. 0.639 for Marginal-PRS), likely due to very slight heterogeneity across simulation replicates that slightly reduced the strength of the association between the genetic risk scores and the diagnostic category in the concatenated data (see Methods).

As an additional point of reference, we also compared the accuracy of DDx-PRS in case vs. rest comparisons to the accuracy of DDx-PRS in case vs. control comparisons (Table S4). (We note that the power to distinguish SCZ vs. control, BIP vs. control, and MDD vs. control was comparable for DDx-PRS, Marginal-PRS and the standard case-control PRS; Table S4.) We anticipated that distinguishing case vs. rest would be a harder problem than distinguishing case vs. control, given the positive genetic correlations between disorders. Indeed, case vs. rest accuracy was lower than case vs. control accuracy for SCZ (AUC of 0.678±0.001 for SCZ vs. rest, 0.745±0.002 for SCZ vs. control) and BIP (0.629±0.002 for BIP vs. rest, 0.701±0.002 for BIP vs. control)—but slightly higher than case vs. control accuracy for MDD (0.618±0.002 for MDD vs. rest, 0.610±0.002 for MDD vs. control), because MDD cases are genetically more similar to controls than to SCZ cases and BIP cases (Figure 1 of ref.^20^). Interestingly, we observed that the AUC of SCZ vs. rest (0.678±0.001) was lower than the average of the AUCs of SCZ vs. control, SCZ vs. BIP, and SCZ vs. MDD (0.695), consistent with the fact that a sample with equal numbers of controls, BIP cases and MDD cases and controls is more genetically similar to SCZ cases than each population considered separately (because averaging populations reduces genetic distance^25^) and hence more difficult to distinguish from SCZ cases. We also note that, despite the large genetic correlation between SCZ and BIP (0.70), there is significant potential to distinguish SCZ vs. BIP (AUC 0.632±0.002).

We performed six secondary analyses. First, we compared DDx-PRS to Marginal-PRS across all 6 pairs of diagnostic categories (SCZ vs. BIP, SCZ vs. MDD, BIP vs. MDD, SCZ vs. control, BIP vs. control, MDD vs. control), instead of 4 pairs of one diagnostic category vs. rest. Analogous to Figure 1, DDx-PRS attained substantially better calibration and slightly higher accuracy than Marginal-PRS (Figure S1 and Table S5). Second, we assessed the calibration of DDx-PRS and Marginal-PRS using calibration slope^3^, instead of ICI. Analogous to Figure 1a, DDx-PRS attained substantially better calibration than Marginal-PRS (e.g. 0.073±0.004 vs. 0.151±0.006 for mean absolute difference of slope from 1; Figure S2 and Table S6). Third, we repeated our simulations while doubling the GWAS training sample size. Once again, DDx-PRS attained substantially better calibration than Marginal-PRS (with ICI values similar to Figure 1a) and slightly higher accuracy than Marginal-PRS (with AUC values larger than Figure 1b for both methods, as expected) (Figure S3 and Table S7). Fourth, we repeated our simulations while decreasing the genetic correlation for SCZ-BIP from 0.7 to 0.3. Once again, we observed that DDx-PRS attained better calibration and slightly higher accuracy than Marginal-PRS, with both methods attaining higher accuracy than in our main simulations due to better discrimination between SCZ and BIP (Figure S4 and Table S8). Fifth, we repeated our simulations without sample overlap among controls of SCZ, BIP and MDD. We determined that our results were largely unchanged (Figure S5 and Table S9). Sixth, we repeated our simulations with unequal test sample sizes for the four diagnostic categories. We determined that our results were largely unchanged, with DDx-PRS remaining well-calibrated when the assumed prior probabilities were equal to the true prior probabilities (Figure S6a-e); when the prior probabilities were misspecified, results were miscalibrated in the expected direction (Figure S6f-m). We conclude that DDx-PRS was well-calibrated, and well-powered in distinguishing each diagnostic category vs. rest for the simulated training sample sizes.

### DDx-PRS distinguishes SCZ, BIP, MDD and controls in PGC data

We applied DDx-PRS to distinguish 4 diagnostic categories (SCZ, BIP, MDD, control) in data from the Psychiatric Genomics Consortium (PGC). The full PGC data that we analyzed consisted of European-ancestry case-control GWAS summary statistics for SCZ^8^ (*N*=53,386 cases and 77,258 controls), BIP^14^ (*N*=41,917 cases and 371,549 controls), and MDD^15,16^ (*N*=173,140 cases and 331,433 controls) (Methods and Table S1). The test data consisted of merged PGC case-control cohorts comprising 11,460 individuals of European ancestry (2,865 of each of SCZ, BIP, MDD, control), subdivided into 4 test cohorts within which SCZ, BIP, MDD, and control samples were matched with respect to country and genotyping platform (GER (N=3,368), UK1 (N=1,136), UK2 (N=6,080), USA (N=876); see Methods and Table S2). For each disorder-specific PGC case-control cohort that was included in one of the 4 merged test cohorts, we constructed corresponding training data sets (used to compute case-control PRS for each disorder) that excluded that PGC case-control cohort. We computed standard errors (SE) on our results based on variation across our four test cohorts, while noting that this does not account for variation in the training data. (We note that there is substantial sample overlap between controls of each disorder in training data^20^, but that DDx-PRS is robust to this overlap; see Simulations). We assessed the performance of DDx-PRS and Marginal-PRS in classifying each diagnostic category vs. the rest, assessing calibration using ICI^21^ (where 0 is optimal) and prediction accuracy using AUC^22^.

The calibration of DDx-PRS and Marginal-PRS are reported in Figure 3a and Table S10. DDx-PRS was well-calibrated with a mean ICI (±SE) of 0.026±0.003 (averaged across the four comparisons), whereas Marginal-PRS was substantially less well-calibrated with a mean ICI of 0.060±0.003. Analogous to simulations (Figure 1a), Marginal-PRS suffered particularly poor calibration in the comparison of control vs. rest (ICI of 0.118±0.003), because Marginal-PRS systematically underestimates the three disorder probabilities and overestimates the control probability, as observed in comparisons of true probability vs. predicted probability for each diagnostic category (Figure 4). We also computed concatenated ICI for PGC data that was concatenated across the 4 test cohorts (Figure 3a and Table S10). The concatenated ICI (averaged across the four comparisons) was slightly smaller than the mean ICI for DDx-PRS (0.023 vs. 0.026), likely because the concatenated results were slightly less noisy due to 4 times larger test sample size. However, the concatenated ICI was only very slightly smaller than the mean ICI for Marginal-PRS (0.059 vs. 0.060), analogous to simulations.

**Figure 3.**
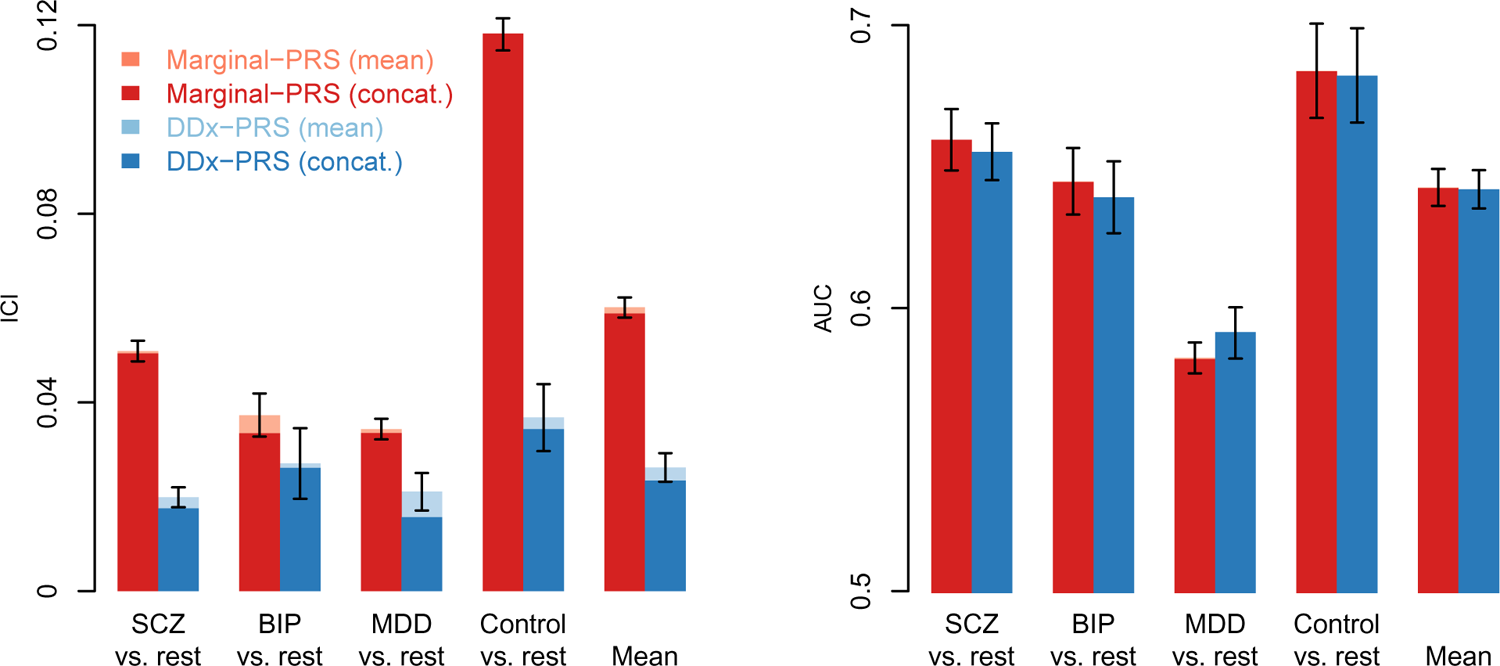
Calibration and accuracy of DDx-PRS and Marginal-PRS in PGC data. We report the calibration (Integrated Calibration Index; ICI) and accuracy (area under the ROC curve; AUC) for the comparisons of schizophrenia (SCZ) vs. rest, bipolar disorder (BIP) vs. rest, major depressive disorder (MDD) vs. rest, control vs. rest and the mean across these four comparisons, for two methods: Marginal-PRS (red) and DDx-PRS (blue). The training data consisted of case-control GWAS summary statistics for SCZ, BIP and MDD; the test data consisted of independent PGC samples subdivided in four test cohorts that were matched with respect to country and genotyping platform (total N=11,460, with 2,865 for each of SCZ, BIP, MDD and control). The mean values across these four test cohorts are displayed in light red and light blue, and results based on data concatenated across these four test cohorts are displayed in dark red and dark blue. Error bars denote standard errors. Numerical results are reported in Table S10.

**Figure 4.**
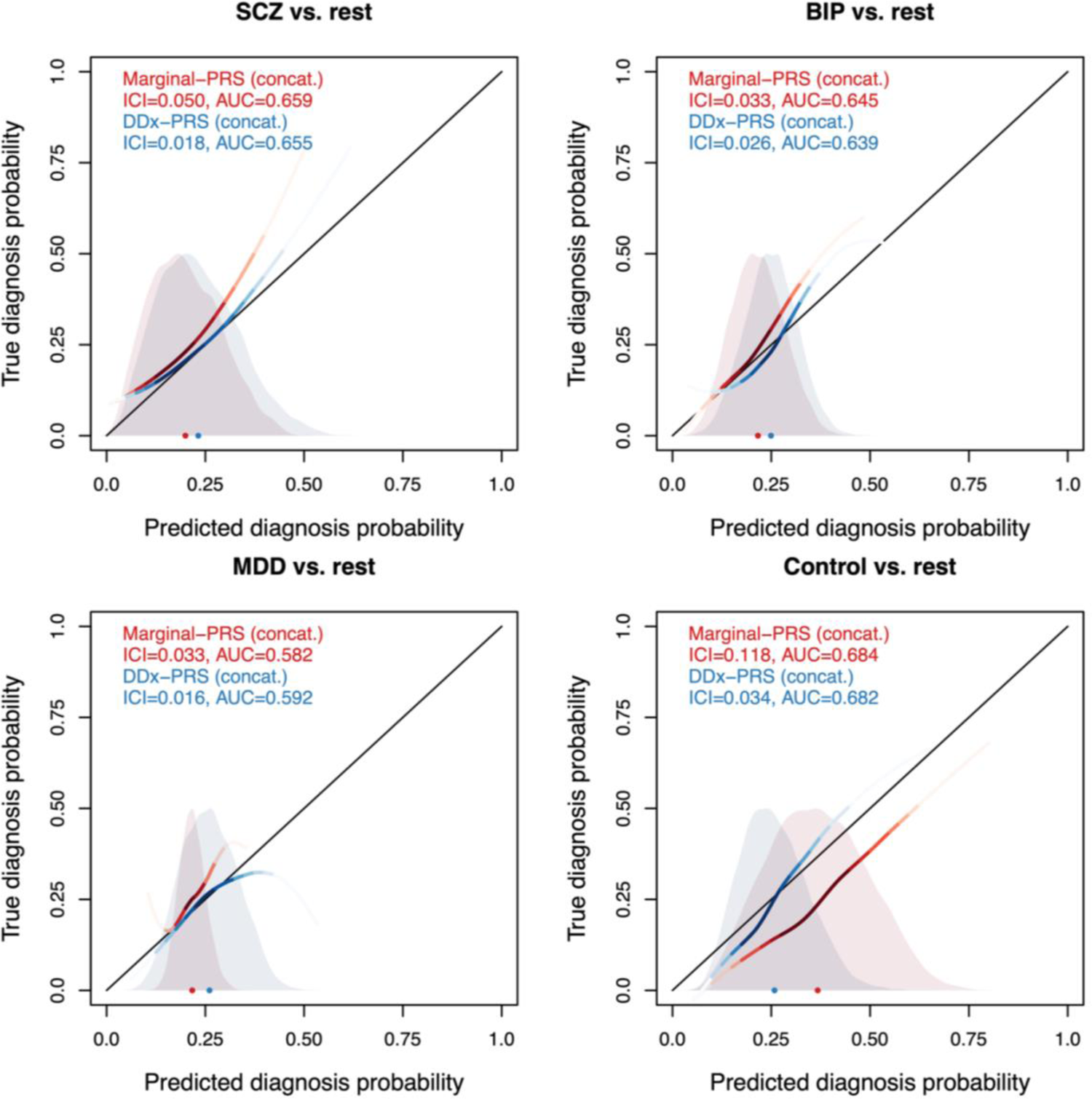
Calibration curves of DDx-PRS and Marginal-PRS in PGC data. We report the true diagnosis probability vs. the predicted diagnosis probability for the comparisons of schizophrenia (SCZ) vs. rest, bipolar disorder (BIP) vs. rest, major depressive disorder (MDD) vs. rest, and control vs. rest, for two methods: Marginal-PRS (red) and DDx-PRS (blue). The calibration curves are plotted using a locally estimated scatterplot smoothing (loess)-based smoothing function. Results are based on the concatenated data across the four PGC test cohorts (total N=11,460, with 2,865 for each of SCZ, BIP, MDD and control). The Integrated Calibration Index (ICI) equals the average absolute difference between the calibration curve and the line y=x (plotted in black), weighted by the density of the predicted diagnosis probabilities (displayed in the shaded histograms and in the color-intensity of the calibration curves). AUC, area under the ROC curve.

The prediction accuracy of DDx-PRS and Marginal-PRS are reported in Figure 3b and Table S10. DDx-PRS attained similar prediction accuracy (mean AUC of 0.642±0.007) as Marginal-PRS (mean AUC of 0.643±0.007), including similar prediction accuracy for each diagnostic category vs. rest (Figure 3b). Analogous to simulations (Figure 1b), the relative performance of DDx-PRS vs. Marginal-PRS was strongest for the MDD vs. rest comparison. Results for concatenated AUC (concatenated across the 4 test cohorts) were very similar to results for mean AUC.

We also compared the accuracy of DDx-PRS in case vs. rest comparisons to the accuracy of DDx-PRS in case vs. control comparisons (Table S11). Case vs. rest accuracy was lower than case vs. control accuracy for SCZ (AUC of 0.655±0.010 for SCZ vs. rest, 0.728±0.016 for SCZ vs. control), BIP (0.639±0.013 for BIP vs. rest, 0.720±0.013 for BIP vs. control), and MDD (0.591±0.009 for MDD vs. rest, 0.618±0.017 for MDD vs. control; non-significant difference), similar to results in simulations. We also observed significant potential to distinguish SCZ vs. BIP (AUC 0.633±0.003; Table S12), despite the large genetic correlation between SCZ and BIP (0.70).

We performed four secondary analyses. First, we repeated our analyses while analysing the four test cohorts separately. We observed similar results for all four test cohorts individually as for the mean results in Figure 3 (Figure S7). Second, we compared DDx-PRS to Marginal-PRS across all 6 pairs of diagnostic categories (SCZ vs. BIP, SCZ vs. MDD, BIP vs. MDD, SCZ vs. control, BIP vs. control, MDD vs. control), instead of 4 pairs of each diagnostic category vs. rest. Analogous to Figure 3, DDx-PRS attained substantially better calibration and similar accuracy as Marginal-PRS (Figure S8 and Table S12). Third, we assessed the calibration of DDx-PRS and Marginal-PRS using calibration slope^3^, instead of ICI. Analogous to Figure 3, DDx-PRS attained better calibration than Marginal-PRS (Figure S9 and Table S13). Fourth, we repeated our analyses at different QC thresholds (less strict QC for SNPs and/or stricter QC for individuals; see Methods). Results were similar to our primary results reported in Figure 3 (Figure S10 and Table S14).

We conclude that DDx-PRS was well-calibrated, and well-powered in distinguishing each diagnostic category vs. rest relative to the PGC training sample sizes analysed.

### Comparison of DDx-PRS to methods that require tuning data

An advantage of DDx-PRS is that it does not require additional tuning data to train parameters connecting case-control PRS to probabilities of each diagnostic category, offering practical advantages for implementation. However, for completeness, we also compared the performance of DDx-PRS to two methods that do require tuning data: DDx-PRS-tuned and Direct-tuned (Table 2 and Methods).

We primarily focused on cross-cohort tuning, in which the tuning data set is from a different cohort than test data (but we also report results for within-cohort tuning). For every possible choice of tuning cohort and test cohort in PGC data (4×3=12 analyses, as we analysed four tuning/test cohorts; Table S2), we restricted tuning data to 200 samples from each of the four diagnostic categories (total *N*_tuning_=800) and evaluated the performance of each method in test data.

The calibration of DDx-PRS, DDx-PRS-tuned and Direct-tuned are reported in Figure 5a and Table S15. The three methods attained similar calibration with mean ICIs (±SE) averaged across the four comparisons of 0.026±0.002, 0.032±0.004, and 0.030±0.002 respectively (Figure 5a and Table S15). A comparison of true vs. predicted probabilities further illustrated that the three methods had similar calibration (Figure 6). We also computed concatenated ICI for PGC data that was concatenated across 4 test cohorts for DDx-PRS (which does not use tuning data) or 12 pairs of tuning-test cohorts for DDx-PRS-tuned and Direct-tuned. The concatenated ICI (averaged across the four comparisons) were slightly smaller than the mean ICI, analogous to Figure 3a.

**Figure 5.**
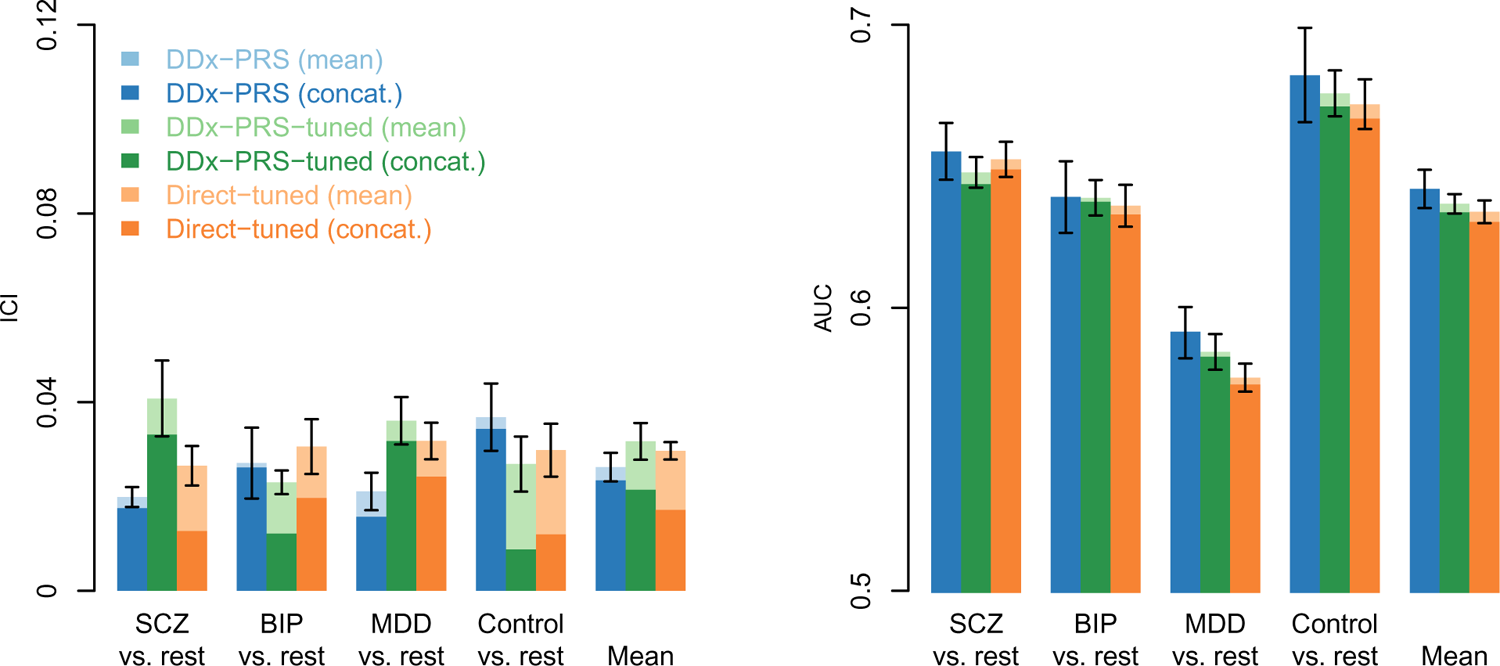
Calibration and accuracy of DDx-PRS, DDx-PRS-tuned and Direct-tuned in PGC data. We report the calibration (Integrated Calibration Index; ICI) and accuracy (area under the ROC curve; AUC) for the comparisons of schizophrenia (SCZ) vs. rest, bipolar disorder (BIP) vs. rest, major depressive disorder (MDD) vs. rest, control vs. rest and the mean across these four comparisons, for three methods: DDx-PRS (blue), DDx-PRS-tuned (green) and Direct-tuned (orange). The training data consisted of case-control GWAS summary statistics for SCZ, BIP and MDD; the test data consisted of independent PGC samples subdivided in four test cohorts that were matched with respect to country and genotyping platform (total N=11,460, with 2,865 for each of SCZ, BIP, MDD and control). For tuning for DDx-PRS-tuned and Direct-tuned, 4×3=12 tuning/test cohort-pairs were analysed with tuning data restricted to 200 samples from each of SCZ, BIP, MDD and control (total *N*_tuning_=800). The mean values across these four test cohorts (DDx-PRS) resp. 12 tuning/test cohort-pairs (DDx-PRS-tuned and Direct-tuned) are displayed in light blue, light green and light orange, and results based on data concatenated across these four test cohorts (resp. 12 tuning/test cohort-pairs) are displayed in dark blue, dark green and dark orange. Error bars denote standard errors. Numerical results are reported in Table S15.

**Figure 6.**
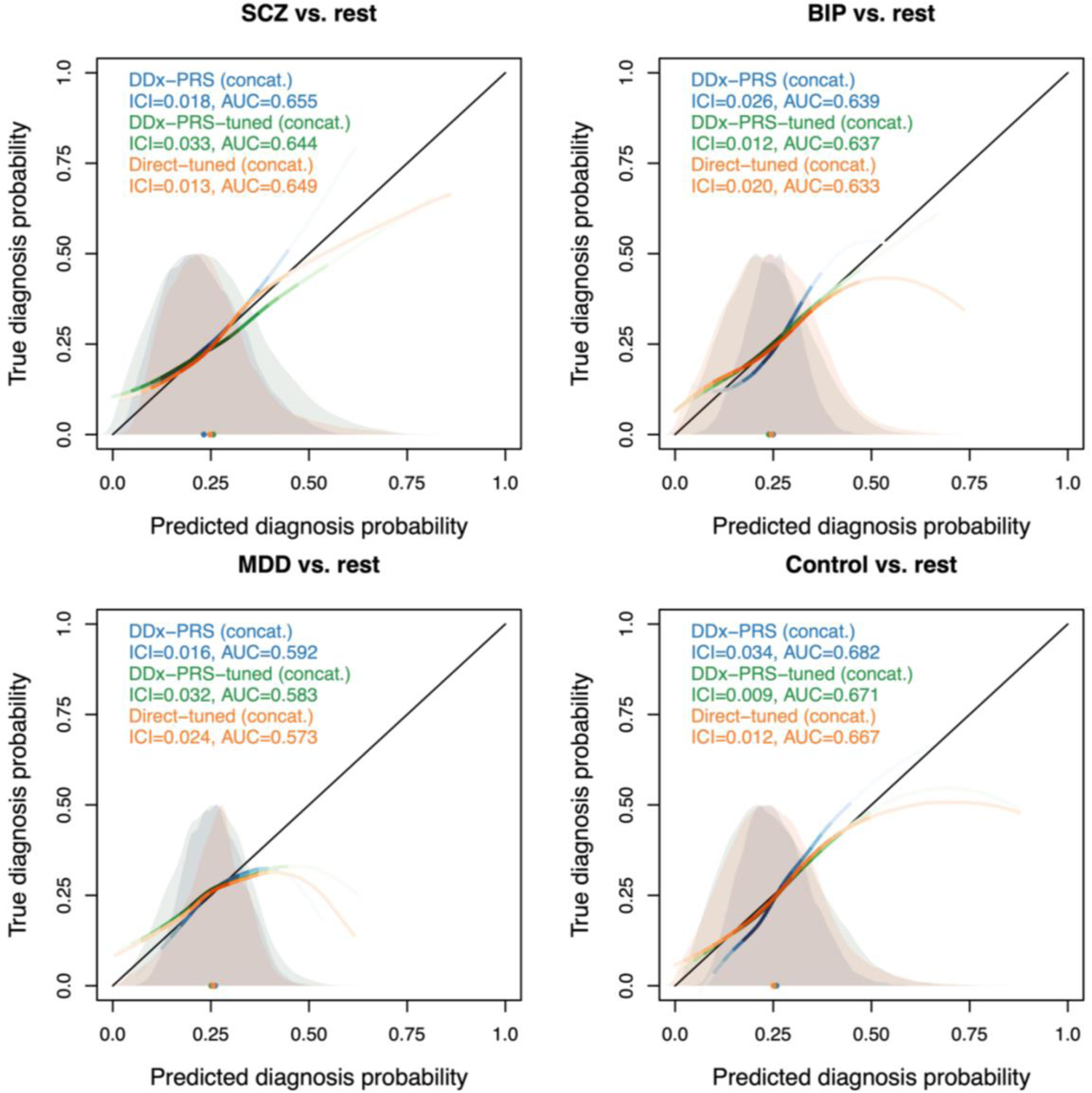
Calibration curves of DDx-PRS, DDx-PRS-tuned and Direct-tuned in PGC data. We report the true diagnosis probability vs. the predicted diagnosis probability for the comparisons of schizophrenia (SCZ) vs. rest, bipolar disorder (BIP) vs. rest, major depressive disorder (MDD) vs. rest, and control vs. rest, for three methods: DDx-PRS (blue), DDx-PRS-tuned (green) and Direct-tuned (orange). Results are based on the concatenated data across the four PGC test cohorts (DDx-PRS) resp. 12 tuning/test cohort-pairs (DDx-PRS-tuned and Direct-tuned). The calibration curves are plotted using a locally estimated scatterplot smoothing (loess)-based smoothing function. The Integrated Calibration Index (ICI) equals the average absolute difference between the calibration curve and the line y=x (plotted in black), weighted by the density of the predicted diagnosis probabilities (displayed in the shaded histograms and in the color-intensity of the calibration curves). AUC, area under the ROC curve.

The prediction accuracy of DDx-PRS, DDx-PRS-tuned and Direct-tuned are reported in Figure 5b and Table S15. The three methods attained comparable prediction accuracies with mean AUCs (±SE) of 0.0642±0.0039, 0.0637±0.0034, and 0.0634±0.0040, respectively. Results for concatenated AUC were very similar to results for mean AUC, analogous to Figure 3b.

We performed three secondary analyses. First, we compared DDx-PRS to DDx-PRS-tuned and Direct-tuned in simulations (Methods). We determined that DDx-PRS, DDx-PRS-tuned and Direct-tuned yielded comparable results (Figure S11 and Table S16). Second, we repeated our PGC analyses with smaller tuning data sets (*N*_tuning_=4×100 and *N*_tuning_=4×50 respectively). We determined that the results were minimally affected by smaller tuning sample size (Figure S12 and Table S17). Third, we compared DDx-PRS to DDx-PRS-tuned and Direct-tuned using within-cohort tuning (*N*_tuning_=4×200), in which the tuning data set is from the same cohort as test data (although this type of tuning data is much less likely to be available in clinical settings). We observed minimal performance improvement of DDx-PRS-tuned and Direct-tuned using within-cohort tuning, and that results for DDx-PRS, DDx-PRS-tuned and Direct-tuned remained comparable (Figure S13 and Table S18).

We conclude that DDx-PRS produces comparable results to DDx-PRS-tuned and Direct-tuned; thus, DDx-PRS is likely to be preferred in most settings (as it does not require tuning data).

### Projections of clinical utility at larger training sample sizes

We sought to assess the clinical utility of DDx-PRS in PGC data and in projections to larger training sample sizes. For each diagnostic category (SCZ, BIP, MDD, control), we computed the true diagnosis probability in each decile of predicted diagnosis probability (similar to previous studies of clinical utility^26–29^.

Results for PGC data are reported in Figure 7 (dashed black curve) and Table S19. The top decile of predicted diagnosis probability yielded true diagnosis probabilities of 0.49 for SCZ, 0.44 for BIP, 0.33 for MDD and 0.47 for controls (close to the predicted diagnosis probabilities; Table S20), substantially above baseline probabilities of 0.25.

**Figure 7.**
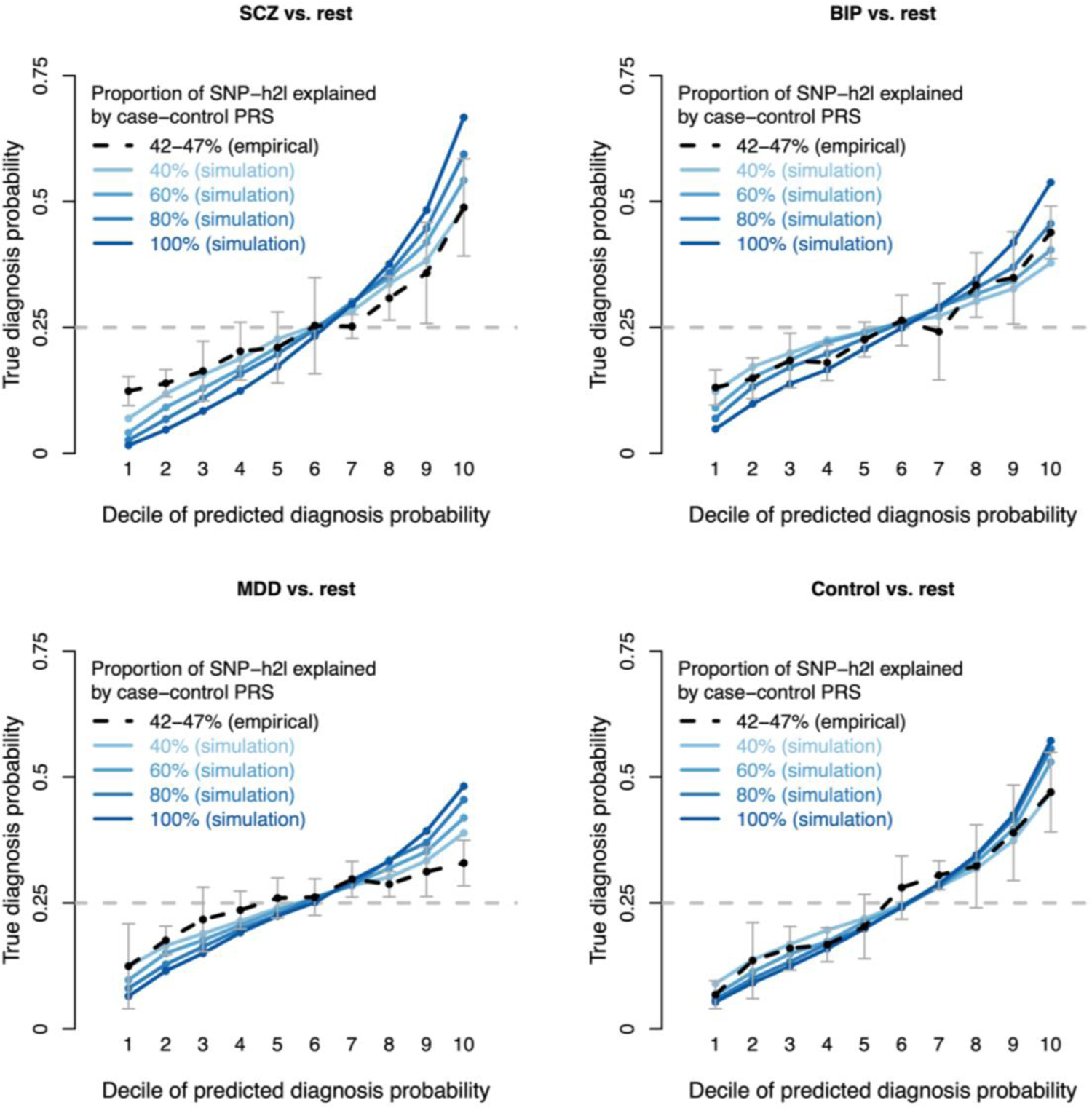
Projections of clinical utility at larger training sample sizes. We report the true diagnosis probability per decile of predicted diagnosis probability for the comparisons of schizophrenia (SCZ) vs. rest, bipolar disorder (BIP) vs. rest, major depressive disorder (MDD) vs. rest, and control vs. rest, for one method: DDx-PRS. Empirical results in PGC data are reported as black points (error bars in grey) connected with dashed black lines; error bars denote 1.96 times the SE across the four PGC test cohorts. Simulations results at various values of SNP-heritability captured by case-control PRS (40%, 60%, 80% and 100%) are reported as blue points connected with solid blue lines, with varying colour intensity. The baseline diagnosis probability (i.e. prior clinical probability) equals 0.25 for each of SCZ, BIP, MDD and control (dashed grey horizontal lines). PGC results for DDx-PRS-tuned and Direct-tuned are reported in Figure S14 and Table S21. Numerical results, including error bars for all data points, are reported in Table S19. The values of the predicted diagnosis probabilities are reported in Table S20.

We computed projections to larger training sample sizes by directly simulating case-control PRS (instead of simulating individual-level SNP data; Figure 1 and Figure 2) at different values of % of SNP-heritability captured by case-control PRS, ranging from 40% (similar to PGC data; 42-47%) to 100% (a best-case scenario, but already approximately attained for height^30^) (Methods). Results for projections to larger training samples sizes are reported in Figure 7 (solid blue curves) and Table S19. In simulations at SNP-heritability captured by case-control PRS equal to 40%, the top decile of predicted diagnosis probability yielded true diagnosis probabilities of 0.49 for SCZ, 0.38 for BIP, 0.39 for MDD and 0.47 for controls, approximately recapitulating analyses of PGC data. In simulations at SNP-heritability captured by case-control PRS equal to 100%, the top decile of predicted diagnosis probability yielded true diagnosis probabilities of 0.67 for SCZ, 0.54 for BIP, 0.48 for MDD and 0.57 for controls, implying considerable potential for clinical utility.

We performed two secondary analyses. First, we repeated the above analyses for DDx-PRS-tuned and Direct-tuned. We determined that results were similar to DDx-PRS (Figure S14 and Table S21). (We did not perform these analyses for Marginal-PRS, as this method is not well-calibrated; Figure 1a and Figure 3a.) Second, we performed simulations at the exact empirical values of % of SNP-heritability captured by case-control PRS, yielding results that were close to the empirical results (Figure S15 and Table S22).

We conclude that DDx-PRS has considerable potential for clinical utility at larger training sample sizes under certain circumstances (see Discussion).

## Discussion

Differential diagnosis is a common clinical challenge, specifically for disorders that are phenotypically (and genetically) similar; in particular, psychiatric disorders often present with non-specific symptoms at disorder onset, but tools to support differential diagnosis are limited. Here, we developed a new method, DDx-PRS, to distinguish related disorders from each other as well as controls by modeling variance/covariance structure across disorders, leveraging case-control PRS and prior clinical probabilities for each diagnostic category. We determined that DDx-PRS is well-calibrated and well-powered based on the training sample sizes analysed, outperforming a simpler method (Marginal-PRS) in both simulated data and empirical PGC data. A strength of DDx-PRS is that is does not use additional tuning data to train parameters connecting case-control PRS to probabilities of each diagnostic category. When comparing DDx-PRS to methods that do require additional tuning data (DDx-PRS-tuned and Direct-tuned), we determined that DDx-PRS produced comparable results, confirming that DDx-PRS is an effective method that is likely to be preferred in most settings as it does not require tuning data. We also assessed the clinical utility of DDx-PRS in projections to larger training sample sizes, indicating considerable potential for future clinical utility under certain circumstances (see below).

We are currently aware of only one previous method for distinguishing different disorders using genetic data (G-PROB)^10^. Unlike DDx-PRS, G-PROB uses only genome-wide significant variants. This may be suitable for distinguishing inflammatory arthritis-causing diseases for which G-PROB was developed, as these diseases are relatively less polygenic^11,12^; however, this approach will have limited utility for highly polygenic disorders such as psychiatric disorders, for which the contribution of genome-wide significant variants is limited^1,2,8,11,12^ (e.g., for SCZ, 2.4% of liability-scale variance is explained by genome-wide significant variants vs. 7.3% explained by a PRS using all genome-wide SNPs^8^). As G-PROB computes risk based on log(OR) effect-sizes, it is unclear how to extend this method to a genome-wide prediction framework. Furthermore, G-PROB is similar to Marginal-PRS in that the disorder probabilities are computed separately for every diagnostic category and later combined into a multi-diagnostic outcome, thus lacking the advantages obtained by modeling the variance/covariance structure across disorders (as in DDx-PRS). Finally, G-PROB aims to distinguish different disorders but does not additionally distinguish disorder cases from controls, the problem that we focus on here.

DDx-PRS has considerable potential for clinical utility, subject to several important considerations. First, increasing the probability for a diagnostic category in differential diagnosis can be clinically useful; for example, for SCZ/BIP/MDD/control, increasing probability from a prior baseline risk of 25% to genetically informed risk of ∼50% may guide clinicians in selecting interventions such as preventive psychotherapy, targeted follow-up, and psychoeducation pertaining to risk factors and early symptoms, which can provide important benefits^31,32^. However, we caution that a diagnostic probability of ∼50% for psychiatric disorders does not justify starting psychiatric medication due to significant potential side effects^33,34^. Second, for clinical utility it is important to select individuals at moderate disorder risk (e.g. 25%); increasing diagnostic category probability from e.g. a prior baseline risk of 1% to genetically informed risk of 3% (or of 95% to 97%) is not clinically useful. Patients at ultra-high risk (UHR)^35,36^ for psychosis (as occurring in SCZ), who are already systematically identified in Mental Health Care^37^ and at increased risk not only for psychosis (long-term risk of 38%)^38^ but also for other disorders^36^, could be a suitable group for implementing DDx-PRS in clinical care. Third, other examples of clinical utility are (i) to distinguish between schizophrenia, bipolar disorder, and depression with psychotic features in patients who present with a psychosis^39^ by specifying a lower prior control probability, or (ii) to distinguish between bipolar disorder and depression in patients who present with a depressive episode by specifying lower SCZ and control prior probabilities. Fourth, ethical aspects should carefully be considered^40–42^. Specifically, not all individuals may be interested to know their risk for SCZ/BIP/MDD: informed consent should be thoroughly provided by trained professionals prior to testing, and opt-out of genetic testing should be made very easy (if not the default). Fifth, future clinical intervention studies will be needed to ultimately determine the clinical utility. Sixth, we note that, although we have emphasized psychiatric disorders in this study, DDx-PRS has potential for clinical utility in other settings; for example, previous studies have emphasized the importance of distinguishing inflammatory arthritis-causing diseases^10^, latent autoimmune diabetes vs. type 2 diabetes^43^, and asthma vs. COPD vs. asthma-COPD overlap syndrome^44^.

We note several limitations of this study. First, DDx-PRS has been developed for within-ancestry prediction, and extending DDx-PRS to cross-ancestry prediction^45–47^ is an important direction for future research to ensure equitable application of genetic prediction. Important progress to improve cross-ancestry prediction accuracy has already been made^29,29, 48–52;^ we additionally highlight the importance of improving the calibration of cross-ancestry prediction^53^. Second, DDx-PRS provides predictions based on polygenic effects of common variants with small effects^5^; extending DDx-PRS to also include rare variants with large effects (such as 22qdel in SCZ^54,55^) is a direction for future research. Even though rare variants may contribute little to prediction accuracy at the group level, rare variants with large effects have high clinical value for individuals who are carriers. Third, in the methods that require tuning data, we did not consider complex multiclass predictors such as XGBoost, which have proven useful in applications involving many predictive features^56^. However, given that we consider only three predictive features (the case-control PRS for SCZ, BIP and MDD) for four diagnostic categories, we anticipate that such predictors would provide little benefit. Fourth, the predicted posterior disorder probabilities computed by DDx-PRS depend strongly on the prior probabilities, which may be misspecified. However, the relative change between the prior and posterior diagnostic category probability is robust to misspecification of the prior, and thus still informative. Nevertheless, further developing strategies to specify accurate prior disorder probabilities (such as screening for UHR^37^) is an important direction for future research. Fifth, integrating risk factors that are strongly correlated with the case-control PRS (e.g. family history) requires further investigation, and model adjustments may be required^57–59^. Despite these limitations, DDx-PRS offers a novel and useful method for leveraging genomic information to inform differential diagnosis.

## Methods

### DDx-PRS method

DDx-PRS consists of 4 steps: (1) estimate prior probabilities of each possible configuration of liabilities; (2) compute case-control PRS; (3) estimate analytical variances and covariances across disorders of liabilities and case-control PRS (overall and for each configuration of liabilities); and (4) estimate posterior probabilities for each test sample. We describe each step in detail below.

*Step 1. Estimate prior probabilities of each possible configuration of liabilities*.

DDx-PRS relies on the fact that, for *n* disorders, there exist 2*^n^* possible configurations, *c*, of liabilities (above or below the liability threshold for each disorder^13^), e.g. 8 possible configurations for 3 disorders; the mapping from configurations of liabilities to diagnostic categories is informed by the PGC classifications used in ref. ^8,14–16^ (Table 1). Given prior clinical probabilities of each diagnostic category, DDx-PRS estimates prior probabilities of each configuration of liabilities by assuming random sampling of liabilities within each diagnostic category. We note that the prior clinical probability will typically be much larger than the population prevalence for help-seeking individuals. For simplicity, we assume a clinical prior probability of 25% for each of SCZ, BIP, MDD and control for the main analyses presented in this paper; however, in application of DDx-PRS, the prior can be specified flexibly by the physician based on e.g. psychiatric examination, questionnaires and/or disorder prevalence in a specific clinical setting.

Specifically, the pairwise genetic correlations SCZ-BIP, SCZ-MDD and BIP-MDD are estimated with cross-trait LDSC^19^. (We note that cross-trait LDSC is robust to sample overlap in controls in training data.^19^) We assume that the correlation of environmental effects equals the genetic correlation^60,61^; thus, the variance-covariance matrices of environmental and genetic effects are specified in the full population, which are added to obtain the variance covariance matrix of liabilities in the full population. Based on the liability thresholds corresponding to the respective disease prevalences (*K*_*SCZ*_ = 0.01, *K*_*BIP*_ = 0.02, *K*_*MDD*_ = 0.16)^8,14,62^, normal theory gives the expected prevalence of the eight configurations in Table 1 in the full population (e.g. 0.0024 for configuration *c*_1_, 0.0017 for configuration *c*_2_, 0.0022 for configuration *c*_3_ and 0.0037 for configuration 4, comprising the diagnostic category SCZ in Table 1). These are now proportionally scaled to add up to the specified prior probability (i.e. 25% for the main analyses in this paper) of diagnostic category (e.g. *P*(*c*_1_) = 0.0591 for configuration *c*_1_, *P*(*c*_2_) = 0.04215 for *c*_2_, *P*(*c*_3_) = 0.0554 for *c*_3_ and *P*(*c*_4_) = 0.0934 for *c*_4_, add up to 0.25 for the diagnostic category SCZ in Table 1). To aid interpretation of the results, we specified the prior probabilities of diagnostic categories throughout this paper at 25% for each of SCZ, BIP, MDD and controls. However, we note that the prior probabilities can be specified flexibly, based on e.g. questionnaires, incidence of psychiatric diagnoses in a specific clinical setting, and/or psychiatric examination by a psychiatrist.

Step 1 of DDx-PRS is computationally fast, with a running time of <10 seconds (excluding the running time of cross-trait LDSC).

*Step 2. Compute case-control PRS*.

DDx-PRS computes case-control PRSs by applying PRS-CS^5^ in two datasets: 1000G^18^ and the PGC test data (see below). The case-control PRSs are scaled to their respective liability scales^13^ by (i) setting *N*_*eff*_ as input in PRS-CS, and (ii) multiplying the posterior mean betas outputted by PRS-CS by {*K*^2^_D_(1 − *K*_*D*_)^2^}⁄{*Z*^2^_D_ ∗ 0.5(1 − 0.5)} where *K*_*D*_ is the prevalence of the respective disorder (i.e. SCZ, BIP or MDD) and *Z*_*D*_ the height of the normal distribution at the liability threshold corresponding to *K*_*D*_ ^23^ (more details on applying PRS-CS are provided below; all analyses in this paper are restricted to European ancestry).

In brief, the intuition of this transformation is that by specifying *N*_eff_ as input in PRS-CS, the posterior mean beta’s will be on the standardized observed scale with 50-50 case-control ascertainment^20^, which are then transformed to the liability scale with Eq 23 of ref.^23^. We note that case-control PRS on the liability scale have the property that the slope of regressing the liability on this PRS has an expected value of 1. A detailed description and evaluation of transforming case-control PRS to the liability scale is provided in ref.^17^. We note that the posterior mean betas could have also been computed with SBayesR^6^ (or any other Bayesian polygenic scoring method that has been shown to be well-calibrated^17^).

We use the individual-level 1000G data to compute the variances/covariances of the case-control PRS (see step 3). However, we note these variances/covariances could have also been estimated using summary-level LD data from 1000G in combination with the posterior mean betas, which would make DDx-PRS a method fully based on summary statistics. Nevertheless, in our analyses, we used the individual-level 1000G data, because 1000G is publicly available and there is no significant computational advantage of used summary-level LD data from 1000G instead of individual-level 1000G data. Instead of 1000G, any other population reference sample could have been used as well for this purpose. However, we specifically did not use the test data to estimate the variances/covariances of the case-control PRS, to ensure full independency of the test data from the data used to train DDx-PRS.

The computational cost of step 2 of DDx-PRS depends on the PRS method being applied, e.g. PRS-CS^5^.

*Step 3. Estimate analytical variances and covariances across disorders of liabilities and case-control PRS (overall and for each configuration of liabilities)*.

For all configurations *c* of liabilities (i.e. the eight rows in Table 1), we compute the means and covariances of case-control PRSs and liabilities as follows.

First, we consider the general population (i.e. not conditioned on case/control status) represented in the 1000G data. In the 1000G data, we estimate the variances and covariances of the case-control PRSs. We note that the variances of the case-control PRSs in the full population equal the variances explained by the case-control PRSs on their respective liability scales, i.e. *var*(*PRS*) = *r*^2^, because the case-control PRSs are well-calibrated on their respective liability scales and because the variance of the liabilities is modelled to be 1 (more details are described in ref.^17^). (We note that estimating and accounting for correlations between case-control PRS across disorders accounts for sample overlap in controls in training data.) We define the following variance/covariance matrix Σ:

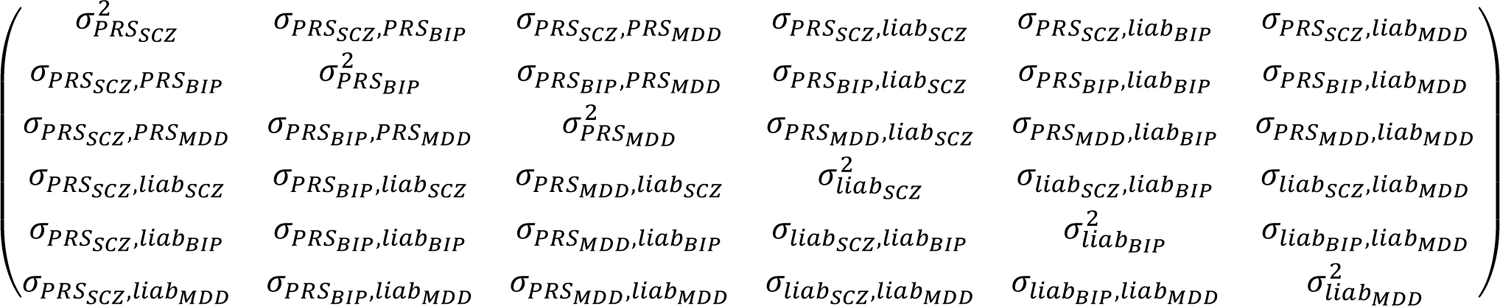

The upper-left 3×3 submatrix follows directly from the described estimates in the 1000G data. The covariances σ_*PRSA*_, *liab*_B_ are computed by 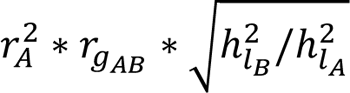, where *r*^2^ = σ^2^_PRSA_ represents the variance explained in *liab*_*A*_ by case-control *PRS*_*A*_; ℎ^2^ (resp. ℎ^2^) the liability scale SNP-heritability of A (resp. B) estimated with LD score regression (based on the baseline LD v2.0 model^63–65);^ and *r*_gAB_ the genetic correlation between disorder A and B estimated with cross-trait LD score regression^19^ (we note that cross-trait LDSC is robust to sample overlap in controls in training data.). The variances σ^2^ equal 1 per model assumptions of the liability scale, and the covariances σ_*liab_A_, liab_B_*_ are estimated by assuming the correlation of environmental effects are equal to the genetic correlation (see also step 1). The means equal (0,0,0,0,0,0) in the general population; this fully defines this six-dimensional normal distribution in the general population. In our application, the (cross-trait) LD score regression is based on the training data (i.e. summary statistics) of the PGC data (see below). The values of Σ inferred in this study are reported in Table S23.

The variances/covariances and means in all 8 configurations *c* of liabilities (i.e. the eight rows in Table 1) follow by iteratively applying theory of the truncated normal distribution, as described previously^66,67^ where truncation is based on the respective disease prevalences (*K*_*SCZ*_ = 0.01, *K*_*BIP*_ = 0.02, *K*_*MDD*_ = 0.16) and their corresponding liability thresholds (*thresh*_*SCZ*_ = 2.326, *thresh*_*BIP*_ = 2.054, *thresh*_*MDD*_ = 0.994). The variances/covariances and means in all 8 configurations *c* of liabilities are reported in Table S23.

For every configuration *c*_*j*_, this provides the means and variances/covariances of the distribution of the three case-control PRS for SCZ, BIP and MDD, which are approximately normally distributed (even though the liabilities are no longer normally distributed after truncating, the case-control PRS are still approximately normally distributed due to the limited proportion (<<1) of variance explained in liabilities by the PRS). Thus, for every value of 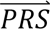_*i*_ = (*PRS*_*SCZ,i*_, *PRS*_*BIP,i*_, *PRS*_*MDD,i*_), the density *P*^(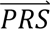^*S* _*i*_|*c*_*j*_) in configuration *c*_*j*_ is defined by the properties of the multivariate normal distribution.

The running time of step 3 of DDx-PRS is <10 seconds.

*Step 4. Estimate posterior probabilities for each test sample*.

In step 4, DDx-PRS applies Bayes’ Theorem to estimate posterior probabilities of each configuration of liabilities for each test sample, conditional on their case-control PRS. Letting *c*_*j*_ denote configuration *j* and 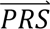_*i*_ denote the case-control PRS for each disorder of individual *i*, these posterior probabilities can be expressed as

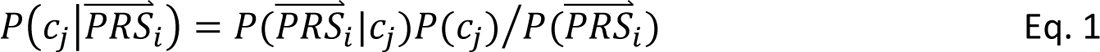

where *P*(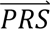|*c*_*j*_) denotes the probability density of 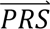_*i*_ given configuration *c*_*j*_ (computed from the output of step 3); *P*(*c*_*j*_) denotes the prior probability of configuration *c*_*j*_ (computed in step 1); and *P*^(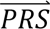^*S* _*i*_) denotes the probability density of 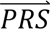_*i*_ summed across all eight configurations weighted by the prior probabilities, i.e. *P*^(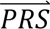^*S* _*i*_) = ∑_*j*′_ *P*^(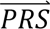^*S* _*i*_|*c*_*j*′_)*P*(*c*_*j*′_). The posterior probabilities of each configuration of liabilities are subsequently combined into posterior probabilities of each diagnostic category using the information in Table 1 (e.g. ∑^4^*P*(*c*_*j*′_^|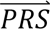^*S* _*i*_) provides the posterior probability of the diagnostic category SCZ).

Step 4 of DDx-PRS is computationally fast, with a running time < 1 minute in analysis of the concatenated PGC data (N=11,460; see below).

### Marginal-PRS method

We compare the results of DDx-PRS to the results of a simpler method, Marginal-PRS (Table 2). Marginal-PRS works in three steps: (1) estimate marginal disorder probabilities; (2) estimate probability to be a control; (3) rescale probabilities to add up to 1.

*Step 1. Estimate marginal disorder probabilities*.

Marginal-PRS computes the marginal disorder probabilities, *P*(*SCZ*|*PRS*_*SCZ,i*_), *P*(*BIP*|*PRS*_*BIP,i*_) and *P*(*MDD*|*PRS*_*MDD,i*_) one disorder at the time using the Bayesian polygenic score Probability Conversion (BPC) approach^17^. Briefly, for every disorder *A*, the BPC approach transforms the case-control PRS to its liability scale and models the mean and variance of the PRS based on 1000G data (analogue to step 2 of DDx-PRS). Subsequently, Bayes’ Theorem is applied to estimate the posterior probability of being a case for disorder *A* as *P*(*D*_*A*_ = 1|*PRS*_*A*_) = *P*(*PRS*_*A*_|*D*_*A*_ = 1)*P*(*D*_*A*_ = 1)⁄*P*(*PRS*_*A*_). The prior probability *P*(*D*_*A*_ = 1) directly follows from the prior probability of the diagnostic categories in Table 1. Marginal-PRS assumes that those with *D*_*A*_ = 0 are random controls (which is not correct as these contain an over representation of the two other diagnostic categories).

*Step 2. Estimate probability to be a control*.

Marginal-PRS computes the probability to be a control as *P*(*control*) = (1 − *P*(*SCZ*))(1 − *P*(*BIP*))(1 − *P*(*MDD*)).

*Step 3. Rescale probabilities to add up to 1*.

Marginal-PRS rescales these four probabilities to add up to 1. We note two important limitations of Marginal-PRS compared to DDx-PRS. First, Marginal-PRS does not model the covariance structure between the respective case-control PRS. Second, when computing marginal disorder probabilities, Marginal-PRS does not account for the other disorders (e.g., when computing *P*(*SCZ*), Marginal-PRS incorrectly assumes that non-SCZ individuals are all controls).

### Methods that require tuning data: DDx-PRS-tuned & Direct-tuned

DDx-PRS and Marginal-PRS do not require additional tuning data to train parameters connecting case-control PRS to probabilities of each diagnostic category. We also considered two methods that do require tuning data: DDx-PRS-tuned and Direct-tuned (Table 2). The tuning data consists of individuals from the four diagnostic categories (SCZ, BIP, MDD and controls) with the case-control PRS for SCZ, BIP and MDD. The tuning data is completely independent from both the training data and the test data.

### DDx-PRS-tuned

DDx-PRS consists of the same 4 steps as DDx-PRS: (1) estimate prior probabilities of each possible configuration of liabilities; (2) compute case-control PRS; (3) estimate analytical variances and covariances across disorders of liabilities and case-control PRS (overall and for each configuration of liabilities); and (4) estimate posterior probabilities for each test sample. We describe the difference between DDx-PRS-tuned and DDx-PRS in detail below.

*Step 1. Estimate prior probabilities of each possible configuration of liabilities*.

Step 1 of DDx-PRS-tuned is the same as step 1 of DDx-PRS (see above).

*Step 2. Compute case-control PRS*.

DDx-PRS-tuned computes case-control PRSs by applying PRS-CS^5^ in two datasets: the PGC tuning data and the PGC test data (see below). The tuning data is used to estimate the variance explained by *PRS*_*A*_ (*r*^2^) based on case vs. control analyses for disorder *A* as is commonly done, i.e. by regressing case/control status on the PRS with subsequently transforming the variance explained to the liability scale^24^ (note that DDx-PRS approximates *r*^2^based on 1000G data; see steps 2 and 3 of DDx-PRS). Subsequently, DDx-PRS-tuned uses the tuning data to scale the *PRS*_*A*_ such that *var*(*PRS*_*A*_) = *r*^2^ based on data of controls (as an approximation of data of the general population; this replaces step 2 in DDx-PRS). The scaling parameters of *PRS*_*A*_ are based on the tuning data and then also applied on every test sample.

*Step 3. Estimate analytical variances and covariances across disorders of liabilities and case-control PRS (overall and for each configuration of liabilities)*.

Step 3 of DDx-PRS-tuned is almost the same as step 3 of DDx-PRS (see above). The only difference is that in DDx-PRS-tuned, the variance/covariance of the case-control PRS (i.e. the upper-left quadrant of Σ above) is based on the case-control PRS in the PGC tuning data (see step 2 of DDx-PRS-tuned).

*Step 4. Estimate posterior probabilities for each test sample*.

Step 4 of DDx-PRS-tuned is the same as step 4 of DDx-PRS (see above).

Other than these two adjustments in steps 2 and 3 of DDx-PRS, DDx-PRS-tuned follows the rest of step 3 and the full steps 1 and 4 of DDx-PRS precisely (specifically, also still modelling the eight configurations *c*_*j*_). Thus, compared to DDx-PRS, DDx-PRS-tuned takes away the uncertainty that comes from the assumptions made to assess *r*^2^based on 1000G data^17^.

### Direct -tuned

Direct-tuned uses a simpler and more direct approach than DDx-PRS-tuned. Direct-tuned consists of 4 steps: (1) specify prior probabilities of each diagnostic category; (2) compute case-control PRS; (3) estimate variances and covariances across case-control PRS (for the diagnostic categories); and (4) estimate posterior probabilities for each test sample. We describe each step in detail below.

*Step 1. Specify prior probabilities of each diagnostic category*.

The prior probabilities of each diagnostic category need to be specified as input for Direct-tuned. (Contrary to DDx-PRS, Direct-tuned does not estimate the prior probabilities of each possible configuration of liabilities; Table 1)

*Step 2. Compute case-control PRS*.

DDx-PRS-tuned computes case-control PRSs by applying PRS-CS^5^ in two datasets: the PGC tuning data and the PGC test data (see below).

*Step 3. Estimate variances and covariances case-control PRS (for the diagnostic categories)*.

For each diagnostic category *dc*_*j*_ (i.e., SCZ/BIP/MDD/control), Direct-tuned estimated the variance/covariances of the case-control PRS in the PGC tuning data (see below). Thus, for every value of 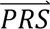_*i*_ = (*PRS*_*SCZ,i*_, *PRS*_*BIP,i*_, *PRS*_*MDD,i*_), the density *P*^(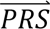^*S* _*i*_ |*dc*_*j*_) in diagnostic category *do*_*j*_ is estimated directly from the tuning data.

*Step 4. Estimate posterior probabilities for each test sample*.

For each test sample, the posterior probability of diagnostic category *do*_*j*_ follows from Bayes’ Theorem

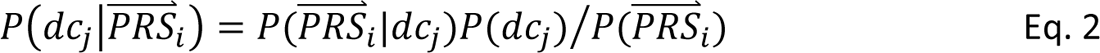

where *P*^(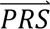^*S* _*i*_|*dc*_*j*_) denotes the probability density of 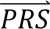_*i*_ given diagnostic category *do*_*j*_ (computed directly from the tuning data); *P*(*dc*_*j*_) denotes the prior probability of diagnostic category *do*_*j*_; and *P*^(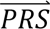^*S* _*i*_) denotes the probability density of 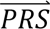_*i*_ summed across all four diagnostic categories weighted by the prior probabilities, i.e. *P*^(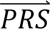^*S* _*i*_) = ∑_*j*′_ *P*^(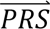^*S* _*i*_|*dc*_*j*′_)*P*(*dc*_*j*′_).

### Main simulations with individual-level SNP data

We simulated data based on the genetic architectures of SCZ^8^, BIP^14^ and MDD^15,16^, with liability-scale SNP-heritabilities^23^ ℎ^2^ of 0.24, 0.19 and 0.09, population prevalences of 1%, 2% and 16%, and liability-scale case-control PRS variance explained^24^ *r*^2^ of 0.10, 0.09 and 0.04, respectively, and genetic correlations of 0.70 for SCZ-BIP, 0.35 for SCZ-MDD and 0.45 for BIP-MDD (in line with our empirical findings; see below).

We simulated individual-level data for *M* = 1,000 SNPs, of which 50% were causal for all three disorders (i.e. number of causal SNPs *M*_*c*_ = 500) and 50% were non-causal for all three disorders. In line with previous work^67,68^, liability-scale SNP effect sizes (β_*i,hSCZ*_, β_*i,hBIP*_ and β_*i,hMDD*_) were drawn from a three dimensional normal distribution with variances ℎ^2^ /*M*_*c*_, ℎ^2^ /*M*_*c*_ and ℎ^2^ /*M*_*c*_; and covariances 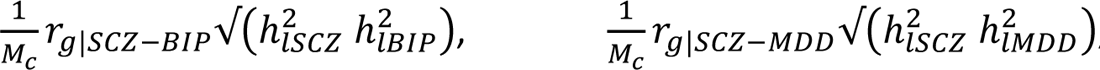 and *1/M_c_r_g_|BIP−MDD√(h^2^_lBIP_ h^2^_lMDD_)*. Effective allele frequencies (*EAF*) of *M* = 1,000 SNPs were drawn from a uniform distribution [0.01,0.5]. Individuals were simulated one-by-one by

1. Randomly assigning *M* = 1,000 genotypes *G*_*i*_ (i.e. 0, 1 or 2 effective alleles) with the probabilities given by the *EAF*s while assuming Hardy-Weinberg equilibrium
2. Defining genetic liabilities as *g*_*lD*_ = ∑ β_*i,hD*_ (*G*_*i*_ − 2*EAF*_*i*_)/√2*EAF*_*i*_(1 − *EAF*_*i*_), for *D* in SCZ, BIP and MDD
3. Defining liabilities as *l*_*D*_ = *g*_*lD*_ + *e*_*lD*_, for *D* in SCZ, BIP and MDD. The environmental effects *e*_*hD*_ are drawn from a three-dimensional normal distribution of *e*_*hSCZ*_, *e*_*hBIP*_ and *e*_*hMDD*_, with variances 1 − ℎ^2^, 1 − ℎ^2^ and 1 − ℎ^2^; and covariances (i.e. assuming the correlation of environmental effects is equal to the correlation of genetic effects).
4. Defining disorder status as *D* = 1 when *h*_*D*_ > *thresh*_*D*_ with *thresh*_*D*_ (resp. *T*_*B*_) corresponding to a population prevalence of *K*_*D*_, for *D* in SCZ, BIP and MDD
5. Subsequently, every individual was assigning to one of the diagnostic categories (SCZ, BIP, MDD or control) based on the link between the configurations of liabilities and the PGC classification in Table 1.

Individuals were simulated until the required number of nonoverlapping cases and controls were obtained: in training samples whose sample size was selected to attain the specified case-control PRS *r*^2^ (690/690 cases/controls for SCZ, 1,411/1,411 for BIP, and 4,542/4,542 for MDD)^69^ and 1,000 test samples for each diagnostic category (SCZ, BIP, MDD, controls). We also simulated 500 random individuals (i.e. not conditioned on case/control status) to serve as analogue of the population reference sample in applying DDx-PRS (see step 2 and 3 above).

We introduced sample overlap of controls in SCZ, BIP and MDD training data to mimic empirical data, which includes substantial overlap of controls^20^. Specifically, the correlation of error-terms was set at 0.2 for SCZ-BIP, 0.1 for SCZ-MDD and 0.1 for BIP-MDD roughly following the empirical findings of the intercept of cross-trait LDSC^19,20^. From these correlation of error terms, the sample overlap is given by the following formula^20^

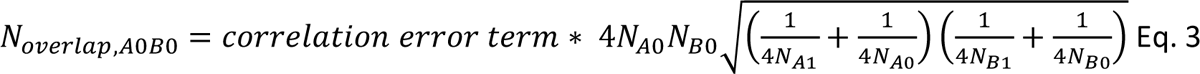

resulting in a sample overlap of controls of 395 for SCZ-BIP, 354 for SCZ-MDD and 506 for BIP-MDD.

After running the three case-control GWAS on the training samples, case-control PRS were computed using Bpred, which analytically computes posterior mean causal effects sizes under a point-normal prior in the special case of no LD^3^. We simulated individual-level SNP data (instead of directly simulating case-control PRS values in test samples) to also assess the performance of the complexities of transforming case-control PRS values to the liability scale (see Methods). We analyzed 50 simulation replicates for each simulation.

We simulated a small number of SNPs (*M*) and training samples (*N*) to limit computational cost, while noting that PRS *r*^2^ primarily depends on *M*/*N*, so that simulations at reduced values of both *M* and *N* are appropriate^3^. Because linkage disequilibrium (LD) does not impact cross-disorder architectures conditional on the above parameter values, we simulated genotypes without LD.

In the results displayed in Figure 1, we note that the concatenated AUC (averaged across the four comparisons) were slightly smaller than the mean AUC (0.645 vs. 0.650 for DDx-PRS and 0.635 vs. 0.639 for Marginal-PRS), which can be explained as follows. The 50 simulation replicates are fully independent, resulting in different causal SNP effect sizes and slightly different heritabilities and genetic correlations across simulation replicates. Thus, all simulation replicates are essentially from different populations that are very similar but not 100% identical, and concatenating data will introduce some very slight heterogeneity to the data causing the decrease of the AUC (we note estimates of the AUC are in principle sample-size independent). The slight discrepancy across simulation replicates is expected to also impact the ICI, but is obscured for the ICI because the ICI is not sample-size independent.

### Psychiatric Genomics Consortium data

The empirical analyses were conducted using data from the Psychiatric Genomics Consortium (PGC)^70^. The full PGC data that we analyzed consisted of European-ancestry case-control GWAS data for SCZ^8^ (76 PGC case-control cohorts; *N*=53,386 cases and 77,258 controls), BIP^14^ (36 PGC case-control cohorts; *N*=41,917 cases and 371,549 controls), and MDD^15,16^ (34 PGC case-control cohorts; *N*=173,140 cases and 331,433 controls, consisting of the data from ref.^16^ excluding 23andMe) (Table S1). We constructed 4 test cohorts by merging selected PGC case-control cohorts; constructed nonoverlapping training data sets for each disorder-specific PGC case-control cohort that was included in one of the 4 test cohorts; applied QC; and computed case-control PRS using PRS-CS and Plink 1.9. We ran S-LDSC and cross-trait LDSC using the full PGC data. Each of these steps is detailed below.

### Construction of 4 test cohorts

The 4 test cohorts were constructed by merging selected PGC case-control cohorts of SCZ^8^, BIP^14^ and MDD^15^. Control test samples were selected from the set of MDD controls, which have more stringent criteria for being labeled as a control (Table 1)^15^. SCZ cases were matched to BIP cases by merging PGC case-control cohorts that were from the same country and had at least 200,000 overlapping SNPs on their genotyping chip; BIP cases were matched to MDD cases with the same criteria. Combining the SCZ-BIP match with the BIP-MDD match also assured an accurate SCZ-MDD match (as confirmed with sensitivity analyses; see below). This resulted in 4 merged test cohorts: GER, UK1, UK2 and USA. From these test cohorts, subjects were randomly selected to obtain sample sizes of exactly 25% of each of SCZ, BIP, MDD and control (the main prior used throughout this study). The resulting test data consisted of 11,460 individuals (2,865 of each of SCZ, BIP, MDD, control): 3,368 in the GER test cohort, 1,136 in UK1, 6,080 in UK2, and 876 in USA. An overview of the PGC case-control cohorts included in the four test cohorts is provided in Table S2.

When applying methods that require tuning data (DDx-PRS-tuned & Direct-tuned), we primarily focused on cross-cohort tuning, designating one test cohort as the tuning cohort and another test cohort as the test cohort. This resulted in 4×3=12 tuning/test cohort pairs used for analyses. We restricted tuning data to 200 samples from each of the four diagnostic categories (total *N*_tuning_=800) and evaluated the performance of each method in test data. In addition to cross-cohort tuning, we also conducted analyses based on within-cohort tuning in the cohorts GER and UK2 (for UK1 and USA, the sample sizes were too small for assessing model performance after excluding 800 samples for tuning).

### Construction of training data sets for each disorder-specific PGC case-control test cohort

For each disorder-specific PGC case-control cohort that was included in one of the 4 test cohorts, we constructed corresponding training data sets (used to compute case-control PRS for each disorder) that excluded that PGC case-control cohort. (We note that the 4 test cohorts included only case samples from SCZ and BIP PGC case-control cohorts, and both case and control samples from MDD PGC case-control cohorts; see above). For PGC SCZ case-control cohort X, the SCZ training data consisted of the full SCZ data excluding PGC SCZ case-control cohort X, the BIP training data consisted of the full BIP data, and the MDD training data consisted of the full MDD data (and analogously for PGC BIP case-control cohort X and PGC MDD case-control cohort X). We tested for sample overlap of controls from each PGC MDD case-control cohort X (used as controls in the 4 test cohorts) and PGC SCZ (resp. BIP) case-control cohorts; when such overlap was identified, the overlapping PGC SCZ (resp. BIP) case-control cohort was excluded from the SCZ (resp. BIP) training data for PGC MDD case-control cohort X. With this strategy, we assured independence between training and test data, while also retaining maximum sample size of the training data. We note that the training data was slightly different across PGC case-control cohorts as different PGC training case-control cohorts were held out, but given the held-out samples are <<10% of the full training samples this had a negligible impact on our results (as confirmed with sensitivity analyses; see below). The precise training data thus used to compute the case-control PRS for every PGC case-control cohort is reported in Table S1.

We performed two sets of sensitivity analyses. First, we verified in the 1000G data that the case-control PRS obtained based on these slightly different training data sets had very high correlations (>0.94 for SCZ, >0.93 for BIP, >0.95 for MDD; see Table S24). We note that despite these very large correlations among case-control PRS, it is still crucial to exclude the test data from the training data, as the inflation in *r*^2^can be profound even when the test data is only a small fraction of the training data^71^. We also note that the slight heterogeneity thus introduced in the test data could result in slight conservative bias of our findings. Second, we computed the *r*^2^ contrasting the SCZ cases (resp. BIP cases) to their matched SCZ controls (resp. BIP controls) based on the original PGC case-control cohorts, and verified that this *r*^2^was highly similar when contrasting the SCZ cases (resp. BIP cases) to the matched MDD controls in the merged test cohorts used in this study (see Table S25).

When applying methods that require tuning data (DDx-PRS-tuned and Direct-tuned), instead of just test data and training data we have 4 sample sets: (i) test data; (ii) tuning data; (iii) training data used to compute PRS in tuning samples (training_for_tuning); and (iv) training data used to compute PRS in test data (training_for_test). As described above, of the 4 available test cohorts, we designate one as the test cohort and one as the tuning cohort, for a total of 4×3=12 tuning/test cohort pairs. The training_for_test and training_for_tuning data are specified as described above (see Table S1). We note that this has the consequence that there is slight overlap between the training_for_tuning data and the test data. We consider this overlap to be of minor importance, for two reasons. First, there is still no overlap between the training_for_test data and the test data. Second, even if the overlap between the training_for_tuning data and the test data improved prediction in test samples, it would do so only for DDx-PRS-tuned and Direct-tuned (and not DDx-PRS, which does not use tuning data). Given that DDx-PRS achieves comparable (or slightly better) performance as DDx-PRS-tuned and Direct-tuned (see Figure 5), any such impact must be minor, and would lead to conservative reporting of relative results for DDx-PRS (our recommended method) vs. DDx-PRS-tuned and Direct-tuned.

### Quality control

Quality control of SNPs was based on previous work^72^. We selected SNPs with imputation INFO>0.9 and MAF>0.10 in both the training and testing data, and we took the intersection of these SNPs with the HapMap3 SNPs included in the PRS-CS LD reference panel (see below). As a secondary analyses to this strict SNP-QC, we also performed analyses retaining SNPS with INFO>0.3 and MAF>0.01 (results in Figure S10 and Table S14).

For quality control of subjects, we tested for sample overlap of controls between the PGC MDD case-control cohorts (used as diagnostic category ‘control’ in this study) and PGC SCZ (resp. BIP) case-control cohorts; when such overlap was found, the respective PGC SCZ (resp. BIP) case-control cohort was excluded from the SCZ (resp. BIP) training data to compute the SCZ (resp. BIP) case-control PRS in the respective PGC MDD case-control cohort. Furthermore, in secondary analyses, we applied stricter subject QC to also exclude individuals from the analyses who were outliers with respect to principal component analyses of the merged cohorts (based on visual screening), SNP missingness >5% pre-imputation, and discrepancy between reported and genotypic sex (results in Figure S10 and Table S14).

### Applying PRS-CS and Plink 1.9 to compute case-control PRS

The case-control PRS in PGC and in 1000G were computed using PRS-CS (version Jun 4, 2021q)^5^. In short, sample size in the input GWAS results for PRS-CS was set at Neff^73^ resulting in posterior mean betas on the standardized observed scale with 50/50 case/control ascertainment^20^, which were subsequently transformed to the liability scale^23^. (Details of thus transforming the PRS to the liability scale are described in ref.^17^; the default PRS-CS command for PRS-CS-auto was used, i.e. “--a=1 --b=0.5--phi=None --beta_std=False --n_iter=1000 --thin=5 --n_burnin=500 ---seed=None”). We note that the posterior mean betas could have also been computed with SBayesR^6^ or any other Bayesian polygenic scoring method that has been shown to be well-calibrated (see ref.^17^ for details).

The lability-scale posterior mean betas were used to compute case-control PRS in PGC and in 1000G with Plink 1.9 (v1.90b7; command “--score header sum center”)^74^. The allele frequencies for the “--score” command were read from 1000G with the “--read-freq” command (the default of Plink is to estimate the allele frequencies from the test data), thereby assuring (i) full independency of the test data and (ii) that DDx-PRS can also be applied on a single individual only.

### S-LDSC and cross-trait LDSC analyses

Based on the full PGC data described above, S-LDSC with the baseline-LD (v2.0) model^63–65^ estimated the liability-scale SNP-heritabilities as 0.23 for SCZ, 0.19 for BIP and 0.09 for MDD, assuming population prevalences of 0.01, 0.02 and 0.16 respectively. Cross-trait LDSC^19^ estimated genetic correlations of 0.71 for SCZ-BIP, 0.34 for SCZ-MDD and 0.45 for BIP-MDD, and cross-trait LDSC intercept (equal to the correlations of error terms^20^) of 0.24 for SCZ-BIP, 0.07 for SCZ-MDD and 0.09 for BIP-MDD. These liability scale SNP heritabilities and genetic correlation estimates are used as input in steps 1 and 3 of DDx-PRS and steps 1 and 3 of DDx-PRS-tuned (see above), and as input parameters for simulations (see above).

### Simulations of case-control PRS directly for projections of clinical utility at larger training sample sizes

We computed projections to larger training sample sizes by directly simulating case-control PRS (instead of simulating individual-level SNP data; see Main simulations above) at different values of % of SNP-heritability captured by case-control PRS, ranging from 40% (similar to PGC data; 42-47%) to 100%. Again, data were simulated with liability-scale SNP-heritabilities^23^ ℎ^2^ of 0.24, 0.19 and 0.09, population prevalences of 1%, 2% and 16%, and genetic correlations of 0.70 for SCZ-BIP, 0.35 for SCZ-MDD and 0.45 for BIP-MDD (in line with our empirical findings; see above).

In this simulation, we first simulated the liability scale genetic component as a three-dimensional normal distribution with variances ℎ^2^, ℎ^2^ and ℎ^2^; covariances and means 0,0,0. We assumed to environmental correlation equal to the genetic correlation, and simulated environmental effects as a three-dimensional normal distribution with variances 1 − ℎ^2^, 1 − ℎ^2^ and 1 − ℎ^2^; covariances and means 0,0,0. Liabilities were computed as *h*_*D*_ = *g*_*hD*_ + *e*_*hD*_, and disease status as *D* = 1 when *h*_*D*_ > *thresh*_*D*_, for *D* in SCZ, BIP and MDD. Every individual was assigning to one of the diagnostic categories (SCZ, BIP, MDD or control) based on the link between the configurations of liabilities and the diagnostic categories in Table 1.

We then added an error term to the genetic components such that (i) the resulting case-control PRS explained the specified amount of variance *r*^2^ in liability (e.g. for the parametrization of case-control PRS explaining 60% in SCZ, *r*^2^ was specified at 0.6 ∗ 0.24 = 0.144), and (ii) the correlation of the case-control PRS was equal to the empirically observed correlation of the case-control PRS in 1000G (0.5 for SCZ-BIP, 0.3 for SCZ-MDD and 0.3 for BIP-MDD; for larger of % of SNP-heritability captured by case-control PRS, the correlation between PRS was set to converged to the *r*_*g*_). Subsequently, the case-control PRS were scaled such that *var*(*PRS*_*D*_) = *r*^2^, for *D* in SCZ, BIP and MDD, thereby assuring the PRS were well-calibrated on the liability scale.

DDx-PRS was applied to this simulated data. We verified that our empirical findings were similar to simulated results at the exact empirical parameters for both simulating PRS directly (as described here) and simulating individual-level SNP data (described above; see Figure S15 and Table S22).

## Code availability

DDx-PRS software: https://github.com/wouterpeyrot/DDxPRS PRS-CS software: https://github.com/getian107/PRScs Plink1.9 software: https://www.cog-genomics.org/plink/

## Data Availability

The full case-control GWAS results for Schizophrenia, Bipolar Disorder and Major Depressive Disorder can be downloaded from the website from the Psychiatric Genomics Consortium (PGC): https://pgc.unc.edu/for-researchers/download-results/. To analyze individual level data of the PGC and GWAS results from subset of the full case-control data, a secondary analysis proposal is required to apply for collaboration.

## Acknowledgements

This research was supported by NIH grants R01 HG006399 and R37 MH107649. We thank the participants who donated their time, life experiences and DNA to this research and the clinical and scientific teams that worked with them. We are deeply indebted to the investigators who comprise the PGC. The PGC has received major funding from the US National Institute of Mental Health (PGC4: R01MH124839, PGC3: U01 MH109528; PGC2: U01 MH094421; PGC1: U01 MH085520). Statistical analyses were carried out on the NL Genetic Cluster Computer (http://www.geneticcluster.org) hosted by SURFsara. The content is solely the responsibility of the authors and does not necessarily represent the official views of the National Institutes of Health.

## Author contributions

WJP and ALP conceived and designed the study and wrote the manuscript. WJP performed the analyses and wrote the software. GP and LMOL merged the PGC case-control cohorts into the test cohorts analyzed. All other authors provided substantial contributions during various phases of the research in shaping the research and software. All authors contributed to writing the manuscript.

