## Supplementary Information for "Distinguishing different psychiatric disorders using DDx-PRS"

1. Department of Psychiatry, Amsterdam UMC, Vrije Universiteit, Amsterdam, The Netherlands; 2. Department of Complex Trait Genetics, Center for Neurogenomics and Cognitive Research, Amsterdam Neuroscience, Vrije Universiteit Amsterdam, Netherlands; 3. Amsterdam Public Health, Amsterdam UMC, Vrije Universiteit, Amsterdam, The Netherlands; 4. Dept. of Psychiatry and Psychotherapy, Charité - Universitätsmedizin, Berlin 10117, Germany; 5. Center for Neurobehavioral Genetics, University of California Los Angeles, Los Angeles, CA, USA; 6. Department of Genetics and Genomics, University of California Los Angeles, Los Angeles, CA, USA; 7. Department of Computational Medicine, University of California Los Angeles, Los Angeles, CA, USA; 8. Division of Psychiatry, University of Edinburgh; 9. Psychiatric and Neurodevelopmental Genetics Unit, Center for Genomic Medicine, Massachusetts General Hospital, Boston, MA, USA; 10. Center for Precision Psychiatry, Department of Psychiatry, Massachusetts General Hospital, Boston, MA, USA; 11. Mental Health and Neuroscience, QIMR Berghofer Medical Research Institute, Brisbane, QLD, Australia; 12. Department of Psychiatry, Icahn School of Medicine at Mount Sinai, New York, NY, US; 13. Charles Bronfman Institute for Personalized Medicine, Icahn School of Medicine at Mount Sinai, New York, NY, US; 14. Department of Genetics and Genomic Sciences, Icahn School of Medicine at Mount Sinai, New York, NY, US; 15. Division of Mental Health and Addiction, Oslo University Hospital, Oslo, NO; 16. NORMENT, University of Oslo, Oslo, NO; 17. Department of Complex Trait Genetics, Center for Neurogenomics and Cognitive Research, Amsterdam Neuroscience, Vrije Universiteit Amsterdam and Vrije Universiteit Amsterdam Medical Center; 18. Stanley Center for Psychiatric Research, Broad Institute of Harvard and MIT, Cambridge MA 02142, USA; 19. German Center for Mental Health (DZPG), site Berlin-Potsdam; 20. Center for Digital Genomic Medicine, Department of Medicine, Vanderbilt University Medical Center, Nashville, TN, US; 21. Departments of Biomedical Informatics and Psychiatry and Behavioral Sciences, Vanderbilt University Medical Center, Nashville, TN, US; 22. Division of Genetic Medicine, Department of Medicine, Vanderbilt Genetics Institute, Vanderbilt University Medical Center, Nashville, TN, US; 23. National Centre for Register-based Research, Aarhus University, Aarhus, Denmark; 24. Bioinformatics Research Centre, Aarhus University, Aarhus, Denmark; 25. Novo Nordisk Foundation Center for Genomics Mechanisms of Disease, Broad Institute of MIT and Harvard, Cambridge, MA, USA; 26. Department of Epidemiology, Harvard T.H. Chan School of Public Health, Boston, MA, USA; 27. Department of Biostatistics, Harvard T.H. Chan School of Public Health, Boston, MA, USA; 28. Program in Medical and Population Genetics, Broad Institute of MIT and Harvard, Cambridge, MA, USA

*

### List of PGC SCZ collaborators

#### PGC BIP Collaborators

Vassily Trubetskoy 1, Antonio F Pardiñas 2, Georgia Panagiotaropoulou 1, Swapnil Awasthi 1, Tim B Bigdeli 3, 267, 397, Charlotte A Dennison 2, Lynsey S Hall 2, Max Lam 4, 268, 398, Oleksandr Frei 5, 269, 399, Alexander L Richards 2, Jakob Grove 6, 270, 400, Zhiqiang Li 7, 271, Mark Adams 8, Ingrid Agartz 5, 272, 401, Elizabeth G Atkinson 9, 273, Esben Agerbo 10, 6, Mariam Al Eissa 11, Margot Albus 12, Madeline Alexander 13, Behrooz Z Alizadeha 14, 274, Köksal Alptekin 15, 275, Thomas D Als 6, 270, 400, Farooq Amin 16, Volker Arolt 17, Manuel Arrojo 18, Lavinia Athanasiu 5, 269, Maria Helena Azevedo 19, Silviu A Bacanu 20, Nicholas J Bass 11, Martin Begemann 21, Richard A Belliveau 22, Judit Bene 23, Beben Benyamin 24, 276, 402, Sarah E Bergen 25, Giuseppe Blasi 26, Julio Bobes 27, 277, 403, Stefano Bonassi 28, Alice Braun 1, Rodrigo Affonseca Bressan 29, 278, Evelyn J Bromet 30, Richard Bruggeman 14, 279, Peter F Buckley 31, Randy L Buckner 32, Jonas Bybjerg-Grauholm 33, 280, Wiepke Cahn 34, 281, Murray J Cairns 35, 282, 404, Monica E Calkins 36, Vaughan J Carr 37, 283, 405, David Castle 38, 284, Stanley V Catts 39, 285, Kimberley D Chambert 40, Raymond CK Chan 41, 286, Boris Chaumette 42, 287, Wei Cheng 43, Eric FC Cheung 44, Siow Ann Chong 45, 288, David Cohen 46, 289, 406, Angèle Consoli 46, 290, Quirino Cordeiro 47, Javier Costas 48, Charles Curtis 49, 291, Michael Davidson 50, Kenneth L Davis 51, Lieuwe de Haan 52, 292, Franziska Degenhardt 53, Lynn E DeLisi 54, 293, Ditte Demontis 6, 270, 400, Faith Dickerson 55, Dimitris Dikeos 56, Timothy Dinan 57, 294, Srdjan Djurovic 58, 295, Jubao Duan 59, 296, Giuseppe Ducci 60, Johan G Eriksson 61, 297, 407, Lourdes Fañanás 62, 298, Stephen V Faraone 63, Alessia Fiorentino 11, Andreas Forstner 53, 299, Josef Frank 64, Nelson B Freimer 65, 300, Menachem Fromer 66, Alessandra Frustaci 67, Ary Gadelha 29, 278, Giulio Genovese 22, Elliot S Gershon 68, Marianna Giannitelli 69, 289, Ina Giegling 70, Paola Giusti-Rodríguez 71, Stephanie Godard 72, Jacqueline I Goldstein 73, Javier González Peñas 74, 301, Ana González-Pinto 75, 301, Srihari Gopal 76, Jacob Gratten 77, 302, Michael F Green 78, 303, Tiffany A Greenwood 79, Olivier Guillin 80, 304, 408, Sinan Gülöksüz 81, 305, Raquel E Gur 36, Ruben C Gur 36, Blanca Gutiérrez 82, Eric Hahn 83, Hakon Hakonarson 84, Vahram Haroutunian 51, 306, 409, Annette M Hartmann 70, Carol Harvey 38, 307, Caroline Hayward 85, Frans A Henskens 86, Stefan Herms 87, Per Hoffmann 87, Daniel P Howrigan 73, 308, Masashi Ikeda 88, Conrad Iyegbe 89, Inge Joa 90, Antonio Julià 91, Anna K Kähler 25, Tony Kam-Thong 92, Yoichiro Kamatani 93, 309, Sena Karachanak-Yankova 94, 310, Oussama Kebir 42, Matthew C Keller 95, Brian J Kelly 86, Andrey Khrunin 96, Sung-Wan Kim 97, Janis Klovins 98, Nikolay Kondratiev 99, Bettina Konte 70, Julia Kraft 1, 311, Michiaki Kubo 100, Vaidutis Kučinskas 101, Zita Ausrele Kučinskiene 101, Agung Kusumawardhani 102, Hana Kuzelova-Ptackova 103, Stefano Landi 104, Laura C Lazzeroni 105, 312, Phil H Lee 106, 22, Sophie E Legge 2, Douglas S Lehrer 107, Rebecca Lencer 17, Bernard Lerer 108, Miaoxin Li 109, Jeffrey Lieberman 110, Gregory A Light 111, 79, Svetlana Limborska 96, Chih-Min Liu 112, 212, Jouko Lönnqvist 113, 313, Carmel M Loughland 114, Jan Lubinski 115, Jurjen J Luykx 116, 314, 410, Amy Lynham 2, Milan Macek Jr 117, Andrew Mackinnon 118, 315, Patrik KE Magnusson 25, Brion S Maher 119, Wolfgang Maier 120, Dolores Malaspina 51, 316, Jacques Mallet 121, Stephen R Marder 122, Sara Marsal 91, Alicia R Martin 73, 317, 411, Lourdes Martorell 123, Manuel Mattheisen 124, 318, 412, Robert W McCarley 125, 319, Colm McDonald 126, John J McGrath 10, 320, 413, Helena Medeiros 127, 321, Sandra Meier 128, 322, Bela Melegh 129, Ingrid Melle 5, 269, Raquelle I Mesholam-Gately 130, 323, Andres Metspalu 131, Patricia T Michie 132, Lili Milani 133, Vihra Milanova 134, Marina Mitjans 21, Espen Molden 135, 324, Esther Molina 136, María Dolores Molto 137, 301, 414, Valeria Mondelli 138, 245, Carmen Moreno 74, 301, Christopher P Morley 139, Gerard Muntané 123, 325, Kieran C Murphy 140, Inez Myin-Germeys 141, Igor Nenadić 142, 326, Gerald Nestadt 143, Liene Nikitina-Zake 98, Cristiano Noto 29, 278, Keith H Nuechterlein 78, Niamh Louise O'Brien 11, F Anthony O'Neill 144, Sang-Yun Oh 145, 327, Ann Olincy 146, Vanessa Kiyomi Ota 147, 278, Christos Pantelis 148, 328, 307, George N Papadimitriou 56, Mara Parellada 74, 301, Tiina Paunio 149, 329, Renata Pellegrino 84, Sathish Periyasamy 150, 330, Diana O Perkins 151, Bruno Pfuhlmann 152, Olli Pietiläinen 153, 331, 415, Jonathan Pimm 154, David Porteous 155, John Powell 156, Diego Quattrone 49, 291, 416, Digby Quested 157, 332, Allen D Radant 158, 333, Antonio Rampino 26, Mark H Rapaport 159, Anna

Rautanen 92, Abraham Reichenberg 51, Cheryl Roe 160, Joshua L Roffman 161, Julian Roth 162, Matthias Rothermundt 17, Bart PF Rutten 163, Safaa Saker-Delye 164, Veikko Salomaa 165, Julio Sanjuan 166, 301, 414, Marcos Leite Santoro 147, 278, Adam Savitz 76, Ulrich Schall 167, 334, Rodney J Scott 35, 335, 417, Larry J Seidman 168, 336, Sally Isabel Sharp 11, Jianxin Shi 169, Larry J Siever 51, 337, Kang Sim 170, 338, 418, Nora Skarabis 1, Petr Slominsky 96, Hon-Cheong So 171, 339, Janet L Sobell 127, Erik Söderman 172, Helen J Stain 173, 340, Nils Eiel Steen 174, 269, Agnes A. Steixner-Kumar 21, Elisabeth Stögmann 175, William S Stone 176, 341, Richard E Straub 177, Fabian Streit 178, Eric Strengman 179, T Scott Stroup 110, Mythily Subramaniam 45, 342, Catherine A Sugar 180, 78, Jaana Suvisaari 165, Dragan M Svrakic 181, Neal R Swerdlow 79, Jin P Szatkiewicz 71, Thi Minh Tam Ta 182, 343, Atsushi Takahashi 163, 183, Chikashi Terao 183, Florence Thibaut 184, 344, Draga Toncheva 94, 345, Paul A Tooney 35, 282, 404, Silvia Torretta 26, Sarah Tosato 185, Gian Battista Tura 186, Bruce I Turetsky 36, Alp Üçok 187, Arne Vaaler 188, 346, Therese van Amelsvoort 163, 245, Ruud van Winkel 189, 163, Juha Veijola 190, 347, John Waddington 191, Henrik Walter 192, Anna Waterreus 193, 348, Bradley T Webb 20, Mark Weiser 194, Nigel M Williams 2, Stephanie H Witt 64, Brandon K Wormley 20, Jing Qin Wu 195, Zhida Xu 196, Robert Yolken 197, Clement C Zai 198, 349, Wei Zhou 199, Feng Zhu 200, 350, Fritz Zimprich 201, Eşref Cem Atbaşoğlu 202, 316, Muhammad Ayub 203, Alessandro Bertolino 26, Donald W Black 204, Nicholas J Bray 2, Gerome Breen 49, Nancy G Buccola 205, William F Byerley 206, Wei J Chen 207, 351, C Robert Cloninger 181, Benedicto Crespo-Facorro 208, 352, Gary Donohoe 209, Robert Freedman 146, Cherrie Galletly 210, 353, 419, Massimo Gennarelli 211, 354, David M Hougaard 33, 280, Hai-Gwo Hwu 212, 355, Assen V Jablensky 213, Steven A McCarroll 22, Jennifer L Moran 40, 356, Ole Mors 6, 357, Preben B Mortensen 10, 6, Bertram Müller-Myhsok 214, 358, 420, Amanda L Neil 215, Merete Nordentoft 33, 359, Michele T Pato 216, 360, Tracey L Petryshen 106, Ann E Pulver 143, Thomas G Schulze 217, 361, 421, Jeremy M Silverman 51, 337, Jordan W Smoller 106, 362, Eli A Stahl 66, 363, 422, Debby W Tsuang 218, 364, Elisabet Vilella 123, Shi-Heng Wang 219, Shuhua Xu 220, 365, 423, Rolf Adolfsson 221, Celso Arango 74, 301, Bernhard T Baune 17, 366, 328, Sintia Iole Belangero 147, 278, Anders D Børglum 6, 270, 400, David Braff 79, 367, Elvira Bramon 222, Joseph D Buxbaum 51, Dominique Campion 80, 304, Jorge A Cervilla 223, Sven Cichon 224, 368, 424, David A Collier 225, Aiden Corvin 226, Marta Di Forti 49, 245, 416, Enrico Domenici 227, Hannelore Ehrenreich 21, Valentina Escott-Price 228, 369, Tõnu Esko 229, 370, Ayman H Fanous 230, 371, 425, Anna Gareeva 231, 372, Micha Gawlik 232, Pablo V Gejman 59, 296, Michael Gill 226, Stephen J Glatt 233, Vera Golimbet 99, Kyung Sue Hong 234, Christina M Hultman 25, Steven E Hyman 40, 373, Nakao Iwata 88, Erik G Jönsson 172, 374, René S Kahn 235, 34, James L Kennedy 198, 349, Elza Khusnutdinova 236, 375, George Kirov 2, James A Knowles 237, 376, Marie-Odile Krebs 42, Claudine Laurent-Levinson 46, 377, Jimmy Lee 238, 378, Todd Lencz 239, 379, 426, Douglas F Levinson 240, Qingqin S Li 76, Jianjun Liu 241, 380, Anil K Malhotra 239, 379, 426, Dheeraj Malhotra 242, Andrew McIntosh 8, Andrew McQuillin 154, Paulo R Menezes 243, Vera A Morgan 244, 381, Derek W Morris 126, Bryan J Mowry 150, 330, Robin M Murray 245, 382, Vishwajit Nimgaonkar 246, Markus M Nöthen 53, Roel A Ophoff 247, 65, 427, Sara A Paciga 248, Aarno Palotie 249, 383, 428, Carlos N Pato 216, 360, Shengying Qin 199, 384, Marcella Rietschel 64, Brien P Riley 20, Margarita Rivera 250, 385, Dan Rujescu 70, Meram C Saka 251, Alan R Sanders 59, 386, Sibylle G Schwab 252, 387, Alessandro Serretti 253, Pak C Sham 254, 388, 429, Yongyong Shi 255, 389, David St Clair 256, Ming T Tsuang 257, 390, Jim van Os 258, 391, Marquis P Vawter 259, Daniel R Weinberger 177, Thomas Werge 260, 392, 430, Dieter B Wildenauer 261, Xin Yu 262, 393, Weihua Yue 262, 393, 431, Peter A Holmans 2, Panos Roussos 263, 394, Evangelos Vassos 49, 245, 432, Danielle Posthuma 264, Ole A Andreassen 5, 269, Kenneth S Kendler 20, Michael J Owen 2, Naomi R Wray 265, 150, Mark J Daly 73, 395, 433, Hailiang Huang 73, 317, 411, Benjamin M Neale 73, 317, Patrick F Sullivan 266, 71, 434, Stephan Ripke 1, 396, 435, James TR Walters 2, Michael C O'Donovan 2

**PGC SCZ Affiliations**

1: Dept. of Psychiatry and Psychotherapy, Charité - Universitätsmedizin, Berlin 10117, Germany, 2: MRC Centre for Neuropsychiatric Genetics and Genomics, Division of Psychiatry and Clinical Neurosciences, Cardiff University, Hadyn Ellis Building, Maindy Road, Cardiff, CF24 4HQ, 3: Department of Psychiatry and the Behavioral Sciences, State University of New York, Downstate Medical Center, 450 Clarkson Ave, Brooklyn, NY, 11203, 4: Stanley Center for Psychiatric Research, Broad Institute of MIT and Harvard, Cambridge, MA 02142, 5: NORMENT Centre, Division of Mental Health and Addiction, University of Oslo, 0424 Oslo, Norway, 6: The Lundbeck Foundation Initiative for Integrative Psychiatric Research (iPSYCH), Aarhus, Denmark, 7: Affiliated Hospital of Qingdao University & Biomedical Sciences Institute of Qingdao University, Qingdao University, 8: Division of Psychiatry, Centre for Clinical Brain Sciences, University of Edinburgh, Royal Edinburgh Hospital, Edinburgh, EH10 5HF, UK, 9: Analytic and Translational Genetics Unit, Massachusetts General Hospital, Boston, MA, 10: National Centre for Register-based Research, Aarhus University, DK-8210 Aarhus, Denmark, 11: Molecular Psychiatry Laboratory, Division of Psychiatry, University College London, London, WC1E 6BT, 12: Comedicum Lindwurmhof, Lindwurmstr. 88, 80337, Munich, Germany, 13: Center for Depression, Anxiety and Stress Research, McLean Hospital, Belmont, MA, USA, 14: University Medical Center Groningen, University Center for Psychiatry, Rob Giel Research Center, Unviersity of Groningen, Groningen, The Netherlands, 15: Department of Psychiatry, Dokuz Eylül University School of Medicine, Izmir, Turkey, 16: Department of Psychiatry and Behavioral Sciences, Emory University, Atlanta, Georgia 30322, USA, 17: Department of Psychiatry, University of Münster, Münster, Germany, 18: Servizo de Psiquiatría, Complexo Hospitalario Universitario de Santiago de Compostela, Servizo Galego de Saúde (SERGAS), Santiago de Compostela, Galicia, Spain, 19: Institute of Medical Psychology, Faculty of Medicine, University of Coimbra, Coimbra, PT, Portugal., 20: Virginia Institute for Psychiatric and Behavioral Genetics, Department of Psychiatry, Virginia Commonwealth University, Richmond, Virginia 23298, USA, 21: Clinical Neuroscience, Max Planck Institute of Experimental Medicine, Göttingen 37075, Germany, 22: Stanley Center for Psychiatric Research, Broad Institute of MIT and Harvard, Cambridge, Massachusetts 02142, USA, 23: Department of Medical Genetics, Medical School, University of Pécs, Pécs, Hungary, 24: Australian Centre for Precision Health, University of South Australia Cancer Research Institute, University of South Australia, Adelaide, Australia, 25: Department of Medical Epidemiology and Biostatistics, Karolinska Institutet, Stockholm SE-17177, Sweden, 26: Department of Basic Medical Science, Neuroscience and Sense Organs, University of Bari 'Aldo Moro', Bari, Italy, 27: Área de Psiquiatría-Universidad de Oviedo, Hospital Universitario Central de Asturias (HUCA), Asturias, Spain, 28: Unit of Clinical and Molecular Epidemiology, IRCCS San Raffaele Pisana, and San Raffaele University, Rome, Italy, 29: Department of Psychiatry, Universidade Federal de Sao Paulo, Rua Major Maragliano, 241- CEP 04017-030, Sao Paulo, SP, Brazil, 30: Department of Psychiatry and Behavioural Health, Stony Brook University, HSC, Level T-10, Stony Brook, NY, 31: School of Medicine, Virginia Commonwealth University,, Richmond VA, USA, 32: Department of Psychology, Harvard University, Cambridge, Massachusetts 02138, USA, 33: The Lundbeck Foundation Initiative for Integrative Psychiatric Research (iPSYCH), Denmark, 34: University Medical Center Utrecht, Department of Psychiatry, Rudolf Magnus Institute of Neuroscience, 3584 Utrecht, The Netherlands, 35: School of Biomedical Sciences and Pharmacy, University of Newcastle, Callaghan NSW 2308, Australia, 36: Department of Psychiatry, University of Pennsylvania, 3400 Spruce Street, Philadelphia, PA 19104, 37: School of Psychiatry, University of New South Wales, Sydney, Australia, 38: Department of Psychiatry, The University of Melbourne, Parkville, Victoria 3010, Australia, 39: Brain and Mind Centre, The University of Sydney, Sydney, NSW, Australia, 40: Stanley Center for Psychitric Research, Broad Institute of MIT and Harvard, Cambridge, MA 02142, 41: Institute of Psychology, Chinese Academy of Science, Beijing 100101, China, 42: INSERM U1266, Institute of Psychiatry and Neuroscience of Paris, Université de Paris, GHU Paris Psychiatrie & Neurosciences, 108 rue de la Santé, 75014 Paris, France, 43: Department of Computer Science, University of North Carolina, Chapel Hill, North Carolina 27514, USA, 44: Castle Peak Hospital, Hong Kong, China, 45: Research Division, Institute of Mental Health, Singapore, 46: Faculté de Médecine

Sorbonne Université, Groupe de Recherche Clinique n°15 - Troubles Psychiatriques et Développement (PSYDEV), Department of Child and Adolescent Psychiatry, Hôpital Universitaire de la Pitié-Salpêtrière, 47-83 Boulevard de l'Hôpital, 75651 Paris Cedex 13, France., 47: Department of Psychiatry, Irmandade da Santa Casa de Misericórdia de São Paulo, Rua Dona Veridiana, 55 - São Paulo - SP - CEP 01238-010, São Paulo, SP, Brazil, 48: Instituto de Investigación Sanitaria (IDIS) de Santiago de Compostela, Complexo Hospitalario Universitario de Santiago de Compostela (CHUS), Servizo Galego de Saúde (SERGAS), Santiago de Compostela, Galicia, Spain., 49: Social, Genetic and Developmental Psychiatry Centre, Institute of Psychiatry, Psychology and Neuroscience, King's College London, London SE5 8AF, UK, 50: University of Nicosia Medical School, Nicosia, Cyprus, 51: Department of Psychiatry, Icahn School of Medicine at Mount Sinai, New York, New York 10029, USA, 52: Department of Psychiatry, Academic Medical Centre, University of Amsterdam, Amsterdam, The Netherlands, 53: Institute of Human Genetics, University of Bonn, D-53127 Bonn, Germany, 54: Cambridge Health Alliance, Cambridge, MA 02139, USA, 55: Sheppard Pratt Health System, Baltimore, MD, USA, 56: First Department of Psychiatry, Medical School, National and Kapodistrian University of Athens, Eginition Hospital, Athens 11528, Greece, 57: Department of Psychiatry and Neurobehavioural Sciences, University College Cork, Cork, Ireland, 58: NORMENT Centre, Department of Clinical Science, University of Bergen, Bergen, Norway, 59: Center for Psychiatric Genetics, NorthShore University HealthSystem, Evanston, IL 60201, USA., 60: Department of Mental Health, ASL Rome 1, 00135 Rome, Italy, 61: Department of General Practice and Primary Health Care, University of Helsinki and Helsinki University Hospital, Helsinki, Finland, 62: Department of Evolutionary Biology, Ecology and Environmental Sciences, Faculty of Biology, University of Barcelona, Spain, 63: Departments of Psychiatry and Neuroscience and Physiology, SUNY Upstate Medical University, Syracuse, New York, USA, 64: Department of Genetic Epidemiology in Psychiatry, Central Institute of Mental Health, Medical Faculty Mannheim, University of Heidelberg, Heidelberg, D-68159 Mannheim, Germany, 65: Department of Human Genetics, David Geffen School of Medicine, University of California, Los Angeles, California 90095, USA, 66: Division of Psychiatric Genomics, Department of Psychiatry, Icahn School of Medicine at Mount Sinai, New York, New York 10029, USA, 67: Barnet, Enfield and Haringey Mental Health NHS Trust, St.Ann's Hospital, St.Ann's Road, London, N15 3 TH, UK, 68: Departments of Psychiatry and Human Genetics, University of Chicago, Chicago, Illinois 60637, USA, 69: Faculté de Médecine Sorbonne Université, Groupe de Recherche Clinique n°15 - Troubles Psychiatriques et Développement (PSYDEV), Department of Child and Adolescent Psychiatry,Hôpital Universitaire de la Pitié-Salpêtrière, 47-83 Boulevard de l'Hôpital, 75651 Paris Cedex 13, France., 70: Department of Psychiatry and Psychotherapy, Medical University of Vienna, Vienna, Austria, 71: Department of Genetics, University of North Carolina, Chapel Hill, North Carolina 27599-7264, USA, 72: Departments of Psychiatry and Human and Molecular Genetics, INSERM, Institut de Myologie, Hôpital de la Pitiè-Salpêtrière, Paris, 75013, France, 73: Analytic and Translational Genetics Unit, Massachusetts General Hospital, Boston, Massachusetts 02114, USA, 74: Department of Child and Adolescent Psychiatry, Hospital General Universitario Gregorio Marañón, School of Medicine, Universidad Complutense, Investigación Sanitaria del Hospital Gregorio Marañón, Madrid, Spain, 75: BIOARABA Health Research Institute. OSI Araba. University Hospital, University of the Basque Country, Vitoria, Spain, 76: Neuroscience Therapeutic Area, Janssen Research and Development, Titusville, New Jersey 08560, USA, 77: Mater Research Institute, University of Queensland, Brisbane, Queensland 4102, Australia, 78: Department of Psychiatry and Biobehavioral Sciences, Geffen School of Medicine, University of California Los Angeles, 10833 Le Conte Ave, Los Angeles, CA 90095, 79: Department of Psychiatry, University of California San Diego, 9500 Gilman Drive, La Jolla, CA 92093-0804, 80: INSERM U1245, 76000 Rouen, Normandie, France, 81: Department of Psychiatry and Neuropsychology, School for Mental Health and Neuroscience, Maastricht University Medical Centre, Maastricht, the Netherlands, 82: Department of Psychiatry, Faculty of Medicine and Biomedical Research Centre (CIBM), University of Granada, Granada, Spain., 83: Department of Psychiatry, Charité - Universitätsmedizin, Berlin, Campus Benjamin Franklin, Germany, 84: Children's Hospital of Philadelphia, Leonard Madlyn Abramson Research Center,3615 Civic Center Boulevard, Suite 1216, Philadelphia, PA , USA 19104-

4318, 85: MRC Human Genetics Unit, University of Edinburgh, Institute of Genetics and Molecular Medicine, Western General Hospital, Edinburgh, UK, 86: School of Medicine and Public Health, University of Newcastle, Newcastle NSW 2308, Australia, 87: Division of Medical Genetics, Department of Biomedicine, University of Basel, Basel, CH-4058, Switzerland, 88: Department of Psychiatry, Fujita Health University School of Medicine, Toyoake, Aichi, 470-1192, Japan, 89: Department of Psychosis Studies, Institute of Psychiatry, Psychology and Neuroscience, King's College London, London SE5 8AF, UK, 90: Regional Centre for Clinical Research in Psychosis, Department of Psychiatry, Stavanger University Hospital, 4011 Stavanger, Norway, 91: Rheumatology Research Group, Vall d'Hebron Research Institute, Barcelona, 08035, Spain, 92: Roche Pharma Research and Early Development, Pharmaceutical Sciences, Roche Innovation Center Basel, F. Hoffman-La Roche Ltd, Grenzacherstrasse 124, 4070 Basel, Switzerland, 93: Laboratory of Complex Trait Genomics, Department of Computational Biology and Medical Sciences, Graduate School of Frontier Sciences, The University of Tokyo, Tokyo,108-8639, Japan., 94: Department of Medical Genetics, Medical University, Sofia 1431, Bulgaria, 95: Institute for Behavioural Genetics, University of Colorado Boulder, Boulder, Colorado 80309, USA, 96: Institute of Molecular Genetics of National Research Centre "Kurchatov Institute", Moscow, Russia, 97: Department of Psychiatry, Chonnam National University Medical School, Gwangju, Korea, 98: Latvian Biomedical Research and Study Centre, Riga, LV-1067, Latvia, 99: Mental Health Research Center, Moscow, Russian Federation, 100: RIKEN Center for Integrative Medical Sciences, Yokohama, Kanagawa, 230-0045, Japan, 101: Faculty of Medicine, Vilnius University, LT-01513 Vilnius, Lithuania, 102: Psychiatry Department, University of Indonesia - Cipto Mangunkusumo National General Hospital, Jakarta Pusat, DKI Jakarta, 10430, Indonesia, 103: Department of Psychiatry, 1st Faculty of Medicine and General University Hospital, Prague, Czech Republic, 104: Dipartimento di Biologia, Universita' di Pisa, Pisa, Italy, 105: Departments of Psychiatry and Behavioral Sciences, Stanford University, 450 Serra Mall, Stanford, CA 94305, 106: Psychiatric and Neurodevelopmental Genetics Unit, Department of Psychiatry and Center for Genomic Medicine, Massachusetts General Hospital, Harvard Medical School, Boston, MA, USA, 107: Department of Psychiatry, Wright State University, 3640 Colonel Gleen Hwy, Dayton, OH, 108: Department of Psychiatry, Hadassah-Hebrew University Medical Center, Jerusalem 91120, Israel, 109: Zhongshan School of Medicine and Key Laboratory of Tropical Diseases Control (SYSU), Sun Yat-sen University, Guangzhou 510080, China, 110: Department of Psychiatry, Columbia University, New York, New York 10032, USA, 111: VISN 22, Mental Illness Research, Education & Clinical Center (MIRECC), VA San Diego Healthcare System, 3350 La Jolla Village Drive, San Diego, CA 92161, 112: Department of Psychiatry, National Taiwan University Hospital, Taipei, Taiwan, 113: Mental Health Unit, Department of Public Health Solutions, National Institute for Health and Welfare, PO Box 30, FI-00271, Helsinki, Finland, 114: Hunter New England Health & University of Newcastle, Newcastle NSW 2308, Australia, 115: Department of Genetics and Pathology, International Hereditary Cancer Center, Pomeranian Medical University in Szczecin, 70-453 Szczecin, Poland, 116: Department of Psychiatry, UMC Utrecht Brain Center, University Medical Centre Utrecht, Utrecht University, Utrecht, The Netherlands, 117: Department of Biology and Medical Genetics, 2nd Faculty of Medicine and University Hospital Motol, 150 06 Prague, Czech Republic, 118: Black Dog Institute, University of new South Wales, NSW, Australia, 119: Department of Mental Health, Bloomberg School of Public Health, Johns Hopkins University, Baltimore, Maryland 21205, USA, 120: Department for Neurodegenerative Diseases and Geriatric Psychiatry, University Hospital Bonn, Bonn, Germany , 121: Asfalia Biologics, iPEPS-ICM, Hôpital Universitaire de la Pitié-Salpêtrière, AP-HP, Paris 75013, France, 122: Semel Institute for Neurosciene, University of California, Los Angeles, 11301 Wilshire Blvd, Los Angeles, CA, 123: Hospital Universitari Institut Pere Mata, IISPV, Universitat Rovira i Virgili, CIBERSAM, Reus, Spain, 124: Department of Psychiatry, Dalhousie University, Halifax NS, Canada, 125: VA Boston Health Care System, Brockton, Massachusetts 02301, USA, 126: Centre for Neuroimaging, Cognition and Genomics (NICOG), National University of Ireland Galway, Galway, Ireland., 127: Department of Psychiatry and the Behavioral Sciences, Keck School of Medicine, University of Southern California, Los Angeles, CA, USA, 128: Department of Psychiatry, Dalhousie University, 5850/5980 University Avenue | PO Box

9700, Halifax, Nova Scotia | B3K 6R8, 129: Department of Medical Genetics, University of Pécs, Pécs H-7624, Hungary, 130: Massachusetts Mental Health Center Public Psychiatry Division of the Beth Israel Deaconess Medical Center, Boston, Massachusetts 02114, USA, 131: Estonian Genome Center, Institute of Genomics, University of Tartu, Tartu 50090, Estonia, 132: School of Psychology, University of Newcastle, Newcastle NSW 2308, Australia, 133: Estonian Genome Center, Institute of Genomics, University of Tartu, Tartu 51010, Estonia, 134: First Psychiatric Clinic, Medical University, Sofia 1431, Bulgaria, 135: Department of Pharmacy, University of Oslo, Oslo, Norway, 136: Department of Nursing, Faculty of Health Sciences and Biomedical Research Centre (CIBM), University of Granada, Granada, Spain., 137: Department of Genetics, Faculty of Biological Sciences, Campus of Burjassot, Universidad de Valencia, Valencia, Spain, 138: Department of Psychological Medicine, Institute of Psychiatry, Psychology, and Neuroscience, King's College London, London, SE5 8AF, UK, 139: Departments of Public Health and Preventive Medicine, Family Medicine, and Psychiatry and Behavioral Sciences, State University of New York, Upstate Medical University, Syracuse, NY, 140: Department of Psychiatry, Royal College of Surgeons in Ireland, Dublin, Ireland, 141: Department for Neurosciences, Center for Contextual Psychiatry, KU Leuven, Leuven, Belgium, 142: Cognitive Neuropsychiatry Lab, Department of Psychiatry and Psychotherapy, Philipps Universität Marburg, Marburg, Germany, 143: Department of Psychiatry and Behavioral Sciences, Johns Hopkins University School of Medicine, Baltimore, Maryland 21205, USA, 144: Centre for Public Health, Institute of Clinical Sciences, Queen's University Belfast, Belfast BT12 6AB, UK, 145: Department of Statistics and Applied Probability, University of California at Santa Barbara, Santa Barbara, California, USA., 146: Department of Psychiatry, University of Colorado Denver, Aurora, Colorado 80045, USA, 147: Department of Morphology and Genetics, Universidade Federal de Sao Paulo, Rua Botucatu, 740, Laboratorio de Genetica, CEP 04023-900, Sao Paulo, SP, Brazil, 148: Melbourne Neuropsychiatry Centre, University of Melbourne & Melbourne Health, Melbourne VIC 3053, Australia, 149: Department of Public Health Solutions, Genomics and Biomarkers Unit, National Institute for Health and Welfare, PO Box 30, FI-00271, Helsinki, 150: Queensland Brain Institute, The University of Queensland, Brisbane, QLD 4072, Australia, 151: Department of Psychiatry, University of North Carolina, Chapel Hill, North Carolina 27599-7160, USA, 152: Clinic of Psychiatry and Psychotherapy, Weißer Hirsch, Dresden, Germany, 153: Department of Stem Cell and Regenerative Biology, Harvard University, Cambridge, MA 02138, USA , 154: Molecular Psychiatry Laboratory, Division of Psychiatry, University College London, London WC1E 6JJ, UK, 155: Centre for Genomic and Experimental Medicine, Institute of Genetics and Molecular Medicine, University of Edinburgh, Western General Hospital, Crewe Road, Edinburgh, EH4 2XU, UK, 156: Department of Basic and Clinical Neuroscience, Institute of Psychiatry, Psychology and Neuroscience, King's College London, London SE5 8AF, UK, 157: Oxford Health NHS Foundation Trust, Warneford Hospital, Oxford, UK, 158: Department of Psychiatry and Behavioral Sciences, University of Washington, 2815 Eastlake Ave E #200, Seattle, WA 98102, 159: Huntsman Mental Health Institute, Department of Psychiatry, University of Utah School of Medicine, Salt Lake City, UT, USA, 160: SUNY Upstate Medical University, Syracuse, New York, USA, 161: Department of Psychiatry, Massachusetts General Hospital, Boston, Massachusetts 02114, USA, 162: Department of Psychiatry, Psychosomatics and Psychotherapy, Julius-Maximilians-Universität Würzburg, Wuerzburg, Germany, 163: Maastricht University Medical Center, Department of Psychiatry and Neuropsychology, School for Mental Health and Neuroscience, Maastricht, The Netherlands, 164: Généthon, 1 bis, Rue de l'Internationale, 91000 Evry, France., 165: THL-Finnish Institute for Health and Welfare, P.O BOX 30, Mannerheimintie 166, FI-00271 Helsinki, Finland, 166: Department of Psychiatry, School of Medicine, University of Valencia, Hospital Clínico Universitario de Valencia, Spain, 167: Priority Centre for Brain & Mental Health Research, The University of Newcastle, Mater Hospital, McAuley Centre, Waratah, New South Wales 2298, Australia, 168: Department of Psychiatry, Harvard Medical School, 25 Shattuck St, Boston, MA 02115, 169: Division of Cancer Epidemiology and Genetics, National Cancer Institute, Bethesda, Maryland 20892, USA, 170: West Region, Institute of Mental Health, Singapore, 171: School of Biomedical Sciences, The Chinese University of Hong Kong, Hong Kong, China, 172: Centre for Psychiatry Research, Department of

Clinical Neuroscience, Karolinska Institutet & Stockholm Health Care Services, Stockholm Region, SE-171 77 Stockholm, Sweden, 173: School of Social and Health Sciences, Leeds Trinity University, Leeds, 174: NORMENT Centre, Institute of Clinical Medicine, University of Oslo, 0424 Oslo, Norway, 175: Department of Clinical Neurology, Medical University of Vienna, 1090 Wien, Austria, 176: Harvard Medical School Department of Psychiatry at Beth Israel Deaconess Medical Center, 330 Brookline Ave, Boston MA 02215, 177: Lieber Institute for Brain Development, Baltimore, Maryland 21205, USA, 178: Department of Genetic Epidemiology in Psychiatry, Central Institute of Mental Health, Medical Faculty Mannheim, University of Heidelberg, Heidelberg,D-68159 Mannheim, Germany, 179: Department of Medical Genetics, University Medical Centre Utrecht, Universiteitsweg 100, 3584 CG, Utrecht, The Netherlands, 180: Department of Biostatistics, Fielding School of Public Health, University of California Los Angeles, 650 Charles E. Young Dr. South, Los Angeles, CA 90095, 181: Department of Psychiatry, Washington University, St. Louis, Missouri 63110, USA, 182: Department of Psychiatry, Campus Benjamin Franklin, Charité – Universitätsmedizin Berlin, 183: Laboratory for Statistical and Translational Genetics, RIKEN Center for Integrative Medical Sciences, Yokohama, Kanagawa, 230-0045, Japan, 184: Université de Paris, Faculté de médecine, Hôpital Cochin-Tarnier, Paris 75006, France, 185: Department of Neuroscience, Biomedicine and Movement Sciences, Section of Psychiatry, University of Verona, 37134 Verona, Italy, 186: Psychiatry Unit, IRCCS Istituto Centro San Giovanni di Dio Fatebenefratelli, Brescia, Italy, 187: Department of Psychiatry, Faculty of Medicine, Istanbul University, Istanbul, Turkey, 188: Division of Mental Health, St. Olav's Hospital, Trondheim University Hospital, Trondheim, Norway, 189: KU Leuven, Department of Neurosciences, Center for Clinical Psychiatry, Leuven, Belgium, 190: Department of Psychiatry, Research Unit of Clinical Neuroscience, University of Oulu, Oulu, Finland, 191: Molecular and Cellular Therapeutics, Royal College of Surgeons in Ireland, Dublin 2, Ireland, 192: Department for Psychiatry and Psychotherapy, CCM Charité Universitätsmedizin Berlin, corporate member of Freie Universität Berlin, Humboldt-Universität zu Berlin, and Berlin Institute of Health, Berlin, Germany, 193: Neuropsychiatric Epidemiology Research Unit, School of Population and Global Health, University of Western Australia, Perth, Australia, 194: Sheba Medical Center, Tel Hashomer 52621, Israel, 195: School of Life and Environmental Sciences, University of Sydney, Sydney, NSW 2006, Australia, 196: Department of Psychiatry, GGz Centraal, The Netherlands, 197: Stanley Neurovirology Laboratory, Johns Hopkins University School of Medicine, Baltimore, MD, 21287, USA., 198: Campbell Family Mental Health Research Institute, Centre for Addiction and Mental Health, Toronto, Ontario, M5T 1R8, Canada, 199: Bio-X Institutes, Key Laboratory for the Genetics of Developmental and Neuropsychiatric Disorders, Ministry of Education, Shanghai Jiao Tong University, Shanghai 200030, PR China, 200: Department of Psychiatry, The First Affiliated Hospital of Xi'an Jiaotong University, 277 Yanta West Road, Xi'an 710061, China., 201: Department of Neurology, Medical University of Vienna, Austria, 202: Department of Psychiatry, Ankara University Faculty of Medicine, Ankara, Turkey, 203: Department of Psychiatry, Queens University Kingston, 191 Portsmouth Avenue, Kingston ON Canada K7M 8A6, 204: Department of Psychiatry, University of Iowa Carver College of Medicine, Iowa City, Iowa 52242, USA, 205: School of Nursing, Louisiana State University Health Sciences Center, New Orleans, Louisiana 70112, USA, 206: Department of Psychiatry, University of California San Francisco, San Francisco, California, 94143 USA, 207: Center for Neuropsychiatric Research, National Health Research Institutes, Zhunan Town, Miaoli County, Taiwan, 208: University of Sevilla, CIBERSAM, Sevilla, Spain, 209: Centre for Neuroimaging, Cognition and Genomics (NICOG), National University of Ireland Galway, Galway, Ireland, 210: Discipline of Psychiatry, Adelaide Medical School, University of Adelaide, Adelaide, SA, Australia., 211: Department of Molecular and Translational Medicine, University of Brescia, Brescia, Italy, 212: Neurobiology and Cognitive Science Center, National Taiwan University, Taipei, Taiwan, 213: Centre for Clinical Research in Neuropsychiatry, The University of Western Australia, Perth, WA, Australia, 214: Max Planck Institute of Psychiatry, 80804 Munich, Germany, 215: Menzies Institute for Medical Research, University of Tasmania, 216: Rutgers University, Robert Wood Johnson Medical School, New Brunswick NJ, USA, 217: Institute of Psychiatric Phenomics and Genomics (IPPG), University Hospital, LMU Munich, Munich, Germany, 218: VA Puget Sound Health Care System, 1660

S. Columbian Way, Seattle, WA 98108D WA 98102, 219: College of Public Health, China Medical University, Taichung, Taiwan, 220: State Key Laboratory of Genetic Engineering and Ministry of Education (MOE) Key Laboratory of Contemporary Anthropology, Collaborative Innovation Center of Genetics and Development, Human Phenome Institute, School of Life Sciences, Fudan University, Shanghai, China, 221: Department of Clinical Sciences, Psychiatry, Umeå University, SE-901 87 Umeå, Sweden, 222: Division of Psychiatry, Department of Mental Health Neuroscience, University College London, London WC1E 6BT, UK, 223: Department of Psychiatry, San Cecilio University Hospital, University of Granada, Granada, Spain., 224: Institute of Medical Genetics and Pathology, University Hospital Basel, Schönbeinstrasse 40, CH-4031 Basel, Switzerland, 225: Eli Lilly and Company, Erl Wood Manor, Sunninghill Road, Windlesham, Surrey, GU20 6PH, UK, 226: Neuropsychiatric Genetics Research Group, Department of Psychiatry, Trinity College Dublin, Dublin, Ireland, 227: Department of Cellular, Computational and Integrative Biology, University of Trento, Trento, Italy, 228: Dementia Research Institute, Cardiff University, Hadyn Ellis Building, Maindy Road, Cardiff, CF24 4HQ, 229: Program in Medical and Population Genetics, The Broad Institute of MIT and Harvard, Cambridge, MA, USA, 230: Department of Psychiatry, Phoenix VA Healthcare System, Phoenix AZ, USA, 231: Department of Human Molecular Genetics of the Institute of Biochemistry and Genetics of the Ufa Federal Research Center of the Russian Academy of Sciences. (IBG UFRC RAS) Ufa, 450054, Prospekt Octyabrya, 71, 232: Department of Psychiatry, Psychosomatics and Psychotherapy, University of Würzburg, Margarete-Höppel-Platz 1, 97080, Würzburg, Germany., 233: Psychiatric Genetic Epidemiology and Neurobiology Laboratory (PsychGENe lab), Department of Psychiatry and Behavioral Sciences, SUNY Upstate Medical University, Syracuse, NY, USA, 234: Department of Psychiatry, Sungkyunkwan University School of Medicine, Samsung Medical Center, Seoul, Korea, 235: Department of Psychiatry, Icahn School of Medicine at Mount Sinai, New York NY, 10029, USA, 236: Institute of Biochemistry and Genetics of the Ufa Federal Research Center of the Russian Academy of Sciences (IBG UFRC RAS), Ufa, Russia, 237: Department of Psychiatry and Zilkha Neurogenetics Institute, Keck School of Medicine at University of Southern California, Los Angeles, California 90089, USA, 238: Department of Psychosis, Institute of Mental Health, Singapore, 239: Division of Psychiatry Research, Zucker Hillside Hospital, 75-59 263rd Street, Glen Oaks, NY, 11004, USA, 240: Department of Psychiatry, Stanford University, 401 Quarry Rd., Stanford, CA 94305-5797, USA., 241: Human Genetics, Genome Institute of Singapore, A*STAR, Singapore 138672, Singapore, 242: Roche Pharma Research and Early Development, Roche Innovation Center Basel, F. Hoffman-La Roche Ltd, Basel, Switzerland, 243: Department of Preventative Medicine, Faculdade de Medicina FMUSP, University of Sao Paulo, Brazil;, 244: Neuropsychiatric Epidemiology Research Unit, School of Population and Global Health M431, University of Western Australia, Perth, Australia 6009, 245: National Institute for Health Research (NIHR) Mental Health Biomedical Research Centre at South London and Maudsley NHS Foundation Trust and King's College London, UK, 246: Department of Psychiatry, University of Pittsburgh, 3811 O'Hara St., Pittsburgh PA 15213, 247: Center for Neurobehavioral Genetics, Semel Institute for Neuroscience and Human Behavior, University of California, Los Angeles, California 90095, USA, 248: Early Clinical Development, Pfizer Worldwide Research and Development, Groton, Connecticut 06340, USA, 249: Institute for Molecular Medicine Finland (FIMM), University of Helsinki, Helsinki, Finland, 250: Department of Biochemistry and Molecular Biology II, Faculty of Pharmacy, University of Granada, Granada, Spain, 251: Department of Psychiatry, School of Medicine, Ankara University, Ankara, Turkey, 252: Faculty of Science, Medicine and Health, School of Chemistry and Molecular Bioscience, University of Wollongong, Wollongong NSW 2522, Australia, 253: Department of Biomedical and Neuromotor Sciences, University of Bologna, Italy., 254: Centre for PanorOmic Sciences, LKS Faculty of Medicine, The University of Hong Kong, 255: Bio-X Institutes, Key Laboratory for the Genetics of Developmental and Neuropsychiatric Disorders (Ministry of Education), Collaborative Innovation Center for Brain Science, Shanghai Jiao Tong University, Shanghai, China., 256: Institute of Medical Sciences, University of Aberdeen, Aberdeen, AB25 2ZD, UK, 257: Center for Behavioral Genomics; Department of Psychiatry; University of California, San Diego; La Jolla, CA 92039; U.S.A., 258: University Medical Center Utrecht, Department of Psychiatry, PO BOX 85500, 3584

GA Utrecht, The Netherlands, 259: Department of Psychiatry & Human Behavior, School of Medicine, University of California, Irvine CA 92697-4260, USA, 260: Institute of Biological Psychiatry, Mental Health Services, Copenhagen University Hospital, Copenhagen, Denmark, 261: School of Psychiatry and Clinical Neurosciences, The University of Western Australia, Perth WA 6009, Australia, 262: Peking University Sixth Hospital, Peking University Institute of Mental Health, Beijing, 100191, China;, 263: Department of Psychiatry, Pamela Sklar Division of Psychiatric Genomics, Friedman Brain Institute, Department of Genetics and Genomic Science and Institute for Data Science and Genomic Technology, Icahn School of Medicine at Mount Sinai, New York, NY, 10029, USA, 264: Department of Functional Genomics, Center for Neurogenomics and Cognitive Research, Neuroscience Campus Amsterdam, VU University, Amsterdam 1081, The Netherlands, 265: Institute for Molecular Bioscience, The University of Queensland, Brisbane, QLD 4067, Australia, 266: Department of Medical Epidemiology and Biostatistics, Karolinska Institutet, Stockholm, Sweden, 267: Institute for Genomic Health, SUNY Downstate Medical Center, Brooklyn, NY, USA, 268: Research Division, Institute of Mental Health, Singapore, Republic of Singapore, 269: Division of Mental Health and Addiction, Oslo University Hospital, 0424 Oslo, Norway, 270: Department of Biomedicine and Centre for Integrative Sequencing (iSEQ), Aarhus University, Aarhus, Denmark., 271: Bio-X Institutes, Key Laboratory for the Genetics of Developmental and Neuropsychiatric Disorders (Ministry of Education), Collaborative Innovation Centre for Brain Science, Shanghai Jiao Tong University, 272: Department of Psychiatric Research, Diakonhjemmet Hospital, Oslo, Norway., 273: Stanley Center for Psychiatric Research, Broad Institute of MIT and Harvard, Cambridge, MA, 274: Department of Epidemiology, University Medical Center Groningen, Unviersity of Groningen, Groningen, The Netherlands, 275: Department of Neuroscience, Dokuz Eylül Univ Graduate School of Health Sciences, 276: UniSA Allied Health & Human Performance, University of South Australia, Adelaide, Australia, 277: Instituto de Investigación Sanitaria del Principado de Asturias (ISPA), Asturias, Spain, 278: Laboratory of Integrative Neuroscience, Universidade Federal de Sao Paulo, Rua Pedro de Toledo, 669 - 3° andar fundos, CEP 04039-032, Sao Paulo, SP, Brazil, 279: University of Groningen, Department of Clinical and Developmental Neuropsychology, Groningen, The Netherlands, 280: Center for Neonatal Screening, Department for Congenital Disorders, Statens Serum Institut, Copenhagen, Denmark, 281: Altrecht, General Menthal Health Care, Utrecht, The Netherlands, 282: Hunter Medical Research Institute, Newcastle, New South Wales, Australia, 283: Department of Psychiatry, Monash University, Melbourne, Australia, 284: St Vincent's Hospital, 41 Victoria Parade, Fitzroy, Victoria3065, Australia., 285: School of Medicine, University of Queensland, Herston, QLD Australia, 286: Department of Psychology, University of Chinese Academy of Sciences, Beijing, China, 287: Department of Psychiatry, McGill University, Montreal, Canada, 288: Saw Swee Hock School of Public Health, University of Singapore, Singapore, 289: Centre de Référence des Maladies Rares à Expression Psychiatrique, Department of Child and Adolescent Psychiatry, AP-HP Sorbonne Université, Hôpital Universitaire de la Pitié-Salpêtrière, 47 - 83 Boulevard de l'Hôpital, 75651 Paris Cedex 13, France, 290: Centre de Référence des Maladies Rares à Expression Psychiatrique, Department of Child and Adolescent Psychiatry, AP-HP Sorbonne Université, Hôpital Universitaire de la Pitié-Salpêtrière, 47 - 83 Boulevard de l'Hôpital, 75651 Paris Cedex 13, France;, 291: National Institute for Health Research (NIHR) Maudsley Biomedical Research Centre at South London and Maudsley NHS Foundation Trust and King's College London, UK, 292: Arkin, Institute for Mental Health, Amsterdam, The Netherlands, 293: Department of Psychiatry, Harvard Medical School, Boston, Massachusetts 02115, USA, 294: APC Microbiome Ireland, University College Cork, Cork, Ireland, 295: Department of Medical Genetics, Oslo University Hospital, 0424 Oslo, Norway, 296: Department of Psychiatry and Behavioral Neurosciences, The University of Chicago, Chicago, IL 60637, USA., 297: Folkhälsan Research Center, Helsinki, Finland, 298: Centro de Investigación Biomédica en Red en Salud Mental (CIBERSAM), Madrid, Spain, 299: Centre for Human Genetics, University of Marburg, D-35033 Marburg, Germany, 300: Department of Psychiatry and Biobehavioral Sciences, University of California, Los Angeles, Los Angeles, CA, USA, 301: Centro de Investigación Biomédica en Red de Salud Mental, Spain (CIBERSAM), 302: Institute for Molecular Biology, University of Queendland, Brisbane, Queensland 4072, Australia, 303: VA Greater Los Angeles

Healthcare System, 11301 Wilshire Boulevard, Los Angeles, CA90073, 304: Centre Hospitalier du Rouvray, Rouen 76000 France, 305: Department of Psychiatry, Yale School of Medicine, New Haven, CT, USA, 306: Department of Neuroscience, Icahn School of Medicine at Mount Sinai, New York, New York 10029, USA, 307: NorthWestern Mental Health, Melbourne, Vic, Australia, 308: Broad Institute of MIT and Harvard, Cambridge, Massachusetts, USA, 309: Laboratory for Statistical Analysis, RIKEN Center for Integrative Medical Sciences, Yokohama, Kanagawa, 230-0045, Japan, 310: Department of Genetics, Faculty of Biology, Sofia University "St. Kliment Ohridski", Sofia 1164, Bulgaria, 311: Berlin School of Mind and Brain, Humboldt-Universität zu Berlin, Berlin, Germany, 312: Department of Biomedical Data Science, Stanford University, CA, 313: Department of Psychiatry, University of Helsinki, PO Box 22, FI-00014, Helsinki, Finland, 314: Department of Translational Neuroscience, UMC Utrecht Brain Center, University Medical Center Utrecht, Utrecht University, Utrecht, The Netherlands., 315: Melbourne School of Population and Global health, University of Melbourne, Victoria, Australia, 316: Department of Genetics & Genomics, Icahn School of Medicine at Mount Sinai, Mount Sinai, NY, USA, 317: Stanley Center for Psychiatric Research, the Broad Institute of MIT and Harvard, Cambridge, MA, USA, 318: Department of Community Health and Epidemiology, Dalhousie University, Halifax Nova Scotia, Canada, 319: Deceased, 320: Queensland Brain Institute, University of Queensland, St Lucia, Queensland, Australia, 321: College of Medicine, SUNY Downstate Health Sciences University, Brooklyn, NY, USA, 322: Department of Biomedicine, Aarhus University, DK-8000 Aarhus C, Denmark, 323: Department of Psychiatry, Harvard Medical School, Boston MA, USA, 324: Center for Psychopharmacology, Diakonhjemmet Hospital, Oslo, Norway, 325: Institut de Biologia Evolutiva (UPF-CSIC), Departament de Ciències Experimentals i de la Salut, Universitat Pompeu Fabra, PRBB, Barcelona, Spain, 326: Department of Psychiatry and Psychotherapy, Jena University Hospital, 07743 Jena, Germany, 327: Computational Research Division, Lawrence Berkeley National Laboratory, Berkeley, California, USA, 328: The Florey Institute of Neuroscience and Mental Health, The University of Melbourne, Parkville, VIC, Australia, 329: Department of Psychiatry and SleepWell Research Program, Faculty of medicine, University of Helsinki and Helsinki University Central Hospital, 330: Queensland Centre for Mental Health Research, The University of Queensland, Brisbane, QLD, Australia, 331: The Stanley Center for Psychiatric Research, The Broad Institute, Cambridge, MA 02142, USA, 332: Department of Psychiatry, University of Oxford, Oxford, OX3 7JX, UK, 333: VA Puget Sound Health Care System, 1660 S. Columbian Way, Seattle, WA 98108, 334: Hunter Medical Research Institute, New Lambton Heights, New South Wales 2305, Australia, 335: Division of Molecular Medicine, NSW Health Pathology North, Newcastle, NSW 2305, Australia, 336: Massachusetts Mental Health Center Public Psychiatry Division of the Beth Israel Deaconess Medical Center, 330 Brookline Ave, Boston, MA 02215, 337: James J. Peters VA Medical Center, 130 W Kingsbridge Road, Bronx, NY 10468, 338: Yoo Loo Lin School of Medicine, National University of Singapore, 339: Department of Psychiatry, The Chinese University of Hong Kong, Hong Kong, China, 340: TIPS - Network for Clinical Research in Psychosis; Stavanger University Hospital, Stavanger, Norway, 341: Massachusetts Mental Health Center, 75 Fenwood Road, MA 02115, 342: Saw Swee Hock School of Public Health, National University of Singapore, 343: Berlin Institute of Health (BIH), 10178 Berlin, Germany, 344: INSERM U1266, Institut de psychiatrie et de neurosciences, Paris, France, 345: Bulgarian Academy of Science, 346: Department of Mental Health, Norwegian University of Science and Technology, Trondheim, Norway, 347: Medical Research Center Oulu, Oulu University Hospital and University of Oulu, Oulu, Finland, 348: Centre for Clinical Research in Neuropsychiatry, Division of Psychiatry, Medical School, University of Western Austral, 349: Department of Psychiatry, University of Toronto, Toronto, ON, Canada, 350: Center for Translational Medicine, The First Affiliated Hospital of Xi'an Jiaotong University, 277 Yanta West Road, Xi’an 710061, China., 351: Institute of Epidemiology and Preventive Medicine, College of Public Health, National Taiwan University, Taipei, Taiwan, 352: Hospital Universitario Virgen del Rocio, Department of Psychiatry, Universidad del Sevilla, Sevilla, Spain, 353: Ramsay Health Care (SA) Mental Health, 354: Genetic Unit, IRCCS Istituto Centro San Giovanni di Dio Fatebenefratelli, Brescia, Italy, 355: Department of Psychiatry, College of Medicine and National Taiwan University Hospital, National Taiwan University, Taipei, Taiwan , 356: Department of

Psychiatry, Massachusetts General Hospital, Boston, MA 02114, USA, 357: Psychosis Research Unit, Aarhus University Hospital, Aarhus, Denmark, 358: Munich Cluster for Systems Neurology, Munich, Germany, 359: Mental Health Services in the Capital Region of Denmark, Mental Health Center Copenhagen, University of Copenhagen, Copenhagen, Denmark, 360: Rutgers University, New Jersey Medical School, Newark NJ, USA, 361: Department of Psychiatry and Behavioral Sciences, SUNY Upstate Medical University, Syracuse, NY, USA, 362: Stanley Center for Psychiatric Research, Broad Institute of MIT and Harvard, Cambridge, MA 02138, USA, 363: Medical and Population Genetics, The Broad Institute of MIT and Harvard, Cambridge, MA, USA, 364: Department of Psychiatry and Behavioral Sciences, University of Washington, Seattle,, 365: School of Life Science and Technology, ShanghaiTech University, Shanghai, China, 366: Department of Psychiatry, Melbourne Medical School, The University of Melbourne, Parkville, VIC, Australia, 367: VISN 22, Mental Illness Research, Education & Clinical Center (MIRECC) , VA San Diego Healthcare System, 3350 La Jolla Village Drive, San Diego, CA 92161, 368: Department of Biomedicine, University of Basel, Basel, Switzerland, 369: MRC Centre for Neuropsychiatric Genetics and Genomics, Cardiff University, Hadyn Ellis Building, Cardiff, CF24 4HQ, 370: Estonian Genome Center, Institute of Genomics, University of Tartu, Tartu, 51010, Estonia, 371: Banner-University Medical Center, Phoenix AZ, USA, 372: Federal State Educational Institution of Highest Education Bashkir State Medical University of Public Health Ministry of Russian Federation (BSMU). Lenina, 3. 450008 Ufa, Russia, 373: Department of Stem Cell and Regnerative Biology, Harvard University, 374: NORMENT, Institute of Clinical Medicine, University of Oslo, 0424 Oslo, Norway, 375: Federal State Educational Institution of Highest Education Bashkir State Medical University of Public Health Ministry of Russian Federation (BSMU), Ufa, Russia, 376: Department of Cell Biology, State University of New York, Downstate Medical Center, 450 Clarkson Ave, Brooklyn NY, 377: Centre de Référence des Maladies Rares à Expression Psychiatrique, Department of Child and Adolescent Psychiatry,AP-HP Sorbonne Université, Hôpital Universitaire de la Pitié-Salpêtrière, 47 - 83 Boulevard de l'Hôpital, 75651 Paris Cedex 13, France., 378: Neuroscience and Mental Health, Lee Kong Chian School of Medicine, Nanyang Technological University, Singapore, 379: Institute of Behavioral Science, Feinstein Institutes for Medical Research, 350 Community Drive, Manhasset, NY, 11030, USA, 380: Department of Medicine, Yong Loo Lin School of Medicine, National University of Singapore, Singapore, 381: Centre for Clinical Research in Neuropsychiatry, Division of Psychiatry, Medical School, University of Western Australia, Perth, Australia 6009, 382: Department of Psychosis Studies, Institute of Psychiatry, King's College London, De Crespigny Park, Denmark Hill, London SE5 8AF, UK, 383: Analytic and Translational Genetics Unit, Department of Medicine, Department of Neurology and Department of Psychiatry Massachusetts General Hospital, Boston, MA, USA., 384: Shanghai Key Laboratory of Psychotic Disorders, Shanghai Mental Health Center, Shanghai Jiao Tong University School of Medicine, Shanghai, China., 385: Institute of Neurosciences, Biomedical Research Center (CIBM), University of Granada, Granada, Spain, 386: Department of Psychiatry and Behavioral Neurosciences, The University of Chicago, Chicago IL, USA, 387: Illawarra Health and Medical Research Institute, Wollongong NSW 2522, Australia, 388: State Key Laboratory of Brain and Cognitive Sciences, LKS Faculty of Medicine, The University of Hong Kong, 389: Affiliated Hospital of Qingdao University and Biomedical Sciences Institute of Qingdao University (Qingdao Branch of SJTU Bio-X Institutes), Qingdao University, Qingdao, China., 390: Institute of Genomic Medicine, University of California, San Diego; La Jolla, CA 92039; U.S.A, 391: King's College London, King's Health Partners, Department of Psychosis Studies, Institute of Psychiatry, London, United Kingdom, 392: Department of Clinical Medicine, University of Copenhagen, Copenhagen, Denmark, 393: National Clinical Research Center for Mental Disorders & NHC Key Laboratory of Mental Health (Peking University) & Chinese Academy of Medical Sciences Research Unit (No.2018RU006), Beijing, 100191, China., 394: Mental Illness Research, Education, and Clinical Center (VISN 2 South), James J. Peters VA Medical Center, Bronx, NY, USA, 395: Broad Institute of MIT and Harvard, Cambridge, Massachusetts, USA., 396: Analytic and Translational Genetics Unit, Massachusetts General Hospital, Boston MA 02114, USA, 397: Department of Psychiatry, Veterans Affairs New York Harbor Healthcare System, Brooklyn, NY, USA, 398: Division of Psychiatry Research, Zucker Hillside Hospital, Glen Oaks NY, USA, 399: Center for

Bioinformatics, Department of Informatics, University of Oslo, PO box 1080, Blindern, 0316 Oslo, Norway, 400: Center for Genomics and Personalized Medicine, Aarhus, Denmark, 401: Centre for Psychiatry Research, Department of Clinical Neuroscience, Karolinska Institutet & Stockholm Health Care Services, Stockholm County Council, Stockholm, Sweden., 402: South Australian Health and Medical Research Institute, Adelaide, South Australia, Australia, 403: Centro de Investigación Biomédica en Red de Salud Mental, Oviedo, Asturias, Spain, 404: Centre for Brain & Mental Health Research, The University of Newcastle, Callaghan, NSW, Australia, 405: Neuroscience Research Australia, Sydney, Australia, 406: Institut des Systèmes Intelligents et de Robotique (ISIR), CNRS UMR7222, Sorbonne Université, Campus Pierre et Marie Curie, Faculté des Sciences et Ingénierie, Pyramide, Tour 55, Boîte courrier 173, 4 Place Jussieu, 75252 Paris Cedex 05, France, 407: Department of Obstetrics & Gynecology, Yong Loo Lin School of Medicine, National University of Singapore, 408: UFR santé, Université de Rouen Normandie, Rouen, France, 409: Mental Illness Research Clinical and Education Center (MIRECC), JJ Peters VA Medical Center, Bronx, NY 10468, USA, 410: Second opinion outpatient clinic, GGNet mental health, Warnsveld, The Netherlands, 411: Department of Medicine, Harvard Medical School, Boston, MA, USA, 412: Department of Biomedicine and Centre for Integrative Sequencing (iSEQ), Aarhus University, Aarhus, Denmark, 413: Queensland Centre for Mental Health Research, The Park Centre for Mental Health, Queensland, Australia, 414: Biomedical Research Institute INCLIVA, Valencia, Spain, 415: Institute for Molecular Medicine Finland, FIMM, University of Helsinki, P.O. BOX 20 FI-00014, Helsinki, Finland, 416: South London and Maudsley NHS Mental Health Foundation Trust, 417: Hunter Medical Research Institute, Newcastle NSW, Australia, 418: Lee Kong Chian School of Medicine, Nanyang Technological University, Singapore, 419: Northern Adelaide Local Health Network, 420: Department of Health Data Science, University of Liverpool, Liverpool, UK, 421: Department of Psychiatry and Psychotherapy, University Medical Center Göttingen, Göttingen, Germany, 422: Regeneron Genetics Center, 423: Center for Excellence in Animal Evolution and Genetics, Chinese Academy of Sciences, Kunming, China, 424: Institute of Neuroscience and Medicine (INM-1), Research Center Juelich, Juelich, Germany, 425: Department of Psychiatry, Veterans Affairs New York Harbor Healthcare System, Brooklyn, NY, USA., 426: Department of Psychiatry, Zucker School of Medicine at Hofstra/Northwell, 500 Hofstra University, Hempstead, NY, 11549, USA, 427: Department of Psychiatry, Erasmus University Medical Center, Rotterdam, The Netherlands, 428: The Stanley Center for Psychiatric Research and Program in Medical and Population Genetics, The Broad Institute of MIT and Harvard, Cambridge, MA, USA., 429: Department of Psychiatry, LKS Faculty of Medicine, The University of Hong Kong, 430: Center for GeoGenetics, GLOBE Institute, University of Copenhagen, Copenhagen, Denmark, 431: PKU-IDG/McGovern Institute for Brain Research, Peking University, Beijing, 100871, China, 432: Oxford Health NHS Foundation Trust, 433: Institute for Molecular Medicine Finland (FIMM), Helsinki, Finland, 434: Department of Psychiatry, University of North Carolina, Chapel Hill, North Carolina 27599-7264, USA, 435: Stanley Center for Psychiatric Research, Broad Institute of MIT and Harvard, Cambridge MA 02142, USA

### List of PGC BIP collaborators

#### PGC BIP Collaborators

Niamh Mullins[1,2,235†], Andreas J. Forstner[3,4,5,235], Kevin S. O'Connell[6,7], Brandon Coombes[8], Jonathan R. I. Coleman[9,10], Zhen Qiao[11], Thomas D. Als[12,13,14], Tim B. Bigdeli[15,16], Sigrid Børte[17,18,19], Julien Bryois[20], Alexander W. Charney[2], Ole Kristian Drange[21,22], Michael J. Gandal[23], Saskia P. Hagenaars[9,10], Masashi Ikeda[24], Nolan Kamitaki[25,26], Minsoo Kim[23], Kristi Krebs[27], Georgia Panagiotaropoulou[28], Brian M. Schilder[1,29,30,31], Laura G. Sloofman[1], Stacy Steinberg[32], Vassily Trubetskoy[28], Bendik S. Winsvold[19,33], Hong-Hee Won[34], Liliya Abramova[35], Kristina Adorjan[36,37], Esben Agerbo[14,38,39], Mariam Al Eissa[40], Diego Albani[41], Ney Alliey-Rodriguez[42,43], Adebayo Anjorin[44], Verneri Antilla[45], Anastasia Antoniou[46], Swapnil Awasthi[28], Ji Hyun Baek[47], Marie Bækvad-Hansen[14,48], Nicholas Bass[40], Michael Bauer[49], Eva C. Beins[3], Sarah E. Bergen[20], Armin Birner[50], Carsten Bøcker Pedersen[14,38,39], Erlend Bøen[51], Marco P. Boks[52], Rosa Bosch[53,54,55,56], Murielle Brum[57], Ben M. Brumpton[19], Nathalie Brunkhorst-Kanaan[57], Monika Budde[36], Jonas Bybjerg-Grauholm[14,48], William Byerley[58], Murray Cairns[59], Miquel Casas[53,54,55,56], Pablo Cervantes[60], Toni-Kim Clarke[61], Cristiana Cruceanu[60,62], Alfredo Cuellar-Barboza[63,64], Julie Cunningham[65], David Curtis[66,67], Piotr M. Czerski[68], Anders M. Dale[69], Nina Dalkner[50], Friederike S. David[3], Franziska Degenhardt[3,70], Srdjan Djurovic[71,72], Amanda L. Dobbyn[1,2], Athanassios Douzenis[46], Torbjørn Elvsåshagen[18,73,74], Valentina Escott-Price[75], I. Nicol Ferrier[76], Alessia Fiorentino[40], Tatiana M. Foroud[77], Liz Forty[75], Josef Frank[78], Oleksandr Frei[6,18], Nelson B. Freimer[23,79], Louise Frisén[80], Katrin Gade[36,81], Julie Garnham[82], Joel Gelernter[83,84,85], Marianne Giørtz Pedersen[14,38,39], Ian R. Gizer[86], Scott D. Gordon[87], Katherine Gordon-Smith[88], Tiffany A. Greenwood[89], Jakob Grove[12,13,14,90], José Guzman-Parra[91], Kyooseob Ha[92], Magnus Haraldsson[93], Martin Hautzinger[94], Urs Heilbronner[36], Dennis Hellgren[20], Stefan Herms[3,95,96], Per Hoffmann[3,95,96], Peter A. Holmans[75], Laura Huckins[1,2], Stéphane Jamain[97,98], Jessica S. Johnson[1,2], Janos L. Kalman[36,37,99], Yoichiro Kamatani[100,101], James L. Kennedy[102,103,104,105], Sarah Kittel-Schneider[57,106], James A. Knowles[107,108], Manolis Kogevinas[109], Maria Koromina[110], Thorsten M. Kranz[57], Henry R. Kranzler[111,112], Michiaki Kubo[113], Ralph Kupka[114,115,116], Steven A. Kushner[117], Catharina Lavebratt[118,119], Jacob Lawrence[120], Markus Leber[121], Heon-Jeong Lee[122], Phil H. Lee[123], Shawn E. Levy[124], Catrin Lewis[75], Calwing Liao[125,126], Susanne Lucae[62], Martin Lundberg[118,119], Donald J. MacIntyre[127], Sigurdur H. Magnusson[32], Wolfgang Maier[128], Adam Maihofer[89], Dolores Malaspina[1,2], Eirini Maratou[129], Lina Martinsson[80], Manuel Mattheisen[12,13,14,106,130], Steven A. McCarroll[25,26], Nathaniel W. McGregor[131], Peter McGuffin[9], James D. McKay[132], Helena Medeiros[108], Sarah E. Medland[87], Vincent Millischer[118,119], Grant W. Montgomery[11], Jennifer L. Moran[25,133], Derek W. Morris[134], Thomas W. Mühleisen[4,95], Niamh O'Brien[40], Claire O'Donovan[82], Loes M. Olde Loohuis[23,79], Lilijana Oruc[135], Sergi Papiol[36,37], Antonio F. Pardiñas[75], Amy Perry[88], Andrea Pfennig[49], Evgenia Porichi[46], James B. Potash[136], Digby Quested[137,138], Towfique Raj[1,29,30,31], Mark H. Rapaport[139], J. Raymond DePaulo[136], Eline J. Regeer[140], John P. Rice[141], Fabio Rivas[91], Margarita Rivera[142,143], Julian Roth[106], Panos Roussos[1,2,29], Douglas M. Ruderfer[144], Cristina Sánchez-Mora[53,54,56,145], Eva C. Schulte[36,37], Fanny Senner[36,37], Sally Sharp[40], Paul D. Shilling[89], Engilbert Sigurdsson[93,146], Lea Sirignano[78], Claire Slaney[82], Olav B. Smeland[6,7], Daniel J. Smith[147], Janet L. Sobell[148], Christine Søholm Hansen[14,48], Maria Soler Artigas[53,54,56,145], Anne T. Spijker[149], Dan J. Stein[150], John S. Strauss[102], Beata Świątkowska[151], Chikashi Terao[101], Thorgeir E. Thorgeirsson[32], Claudio Toma[152,153,154], Paul Tooney[59], Evangelia-Eirini Tsermpini[110], Marquis P. Vawter[155], Helmut Vedder[156], James T. R. Walters[75], Stephanie H. Witt[78], Simon Xi[157], Wei Xu[158], Jessica Mei Kay Yang[75], Allan H. Young[159,160], Hannah Young[1], Peter P. Zandi[136], Hang Zhou[83,84], Lea Zillich[78], HUNT All-In Psychiatry[*], Rolf Adolfsson[161], Ingrid Agartz[51,130,162], Martin Alda[82,163], Lars Alfredsson[164], Gulja Babadjanova[165], Lena Backlund[118,119], Bernhard T. Baune[166,167,168], Frank Bellivier[169,170], Susanne Bengesser[50], Wade H. Berrettini[171], Douglas H. R. Blackwood[61], Michael Boehnke[172], Anders D. Børglum[14,173,174], Gerome Breen[9,10], Vaughan J. Carr[175], Stanley Catts[176], Aiden Corvin[177], Nicholas Craddock[75], Udo Dannlowski[166], Dimitris Dikeos[178], Tõnu Esko[26,27,179,180], Bruno Etain[169,170], Panagiotis Ferentinos[9,46], Mark Frye[64], Janice M. Fullerton[152,153], Micha Gawlik[106], Elliot S. Gershon[42,181], Fernando S. Goes[136], Melissa J. Green[152,175], Maria Grigoroiu-Serbanescu[182], Joanna Hauser[68], Frans Henskens[59], Jan Hillert[80], Kyung Sue Hong[47], David M. Hougaard[14,48], Christina M. Hultman[20], Kristian Hveem[19,183], Nakao Iwata[24], Assen V.

Jablensky[184], Ian Jones[75], Lisa A. Jones[88], René S. Kahn[2,52], John R. Kelsoe[89], George Kirov[75], Mikael Landén[20,185], Marion Leboyer[97,98,186], Cathryn M. Lewis[9,10,187], Qingqin S. Li[188], Jolanta Lissowska[189], Christine Lochner[190], Carmel Loughland[59], Nicholas G. Martin[87,191], Carol A. Mathews[192], Fermin Mayoral[91], Susan L. McElroy[193], Andrew M. McIntosh[127,194], Francis J. McMahon[195], Ingrid Melle[6,196], Patricia Michie[59], Lili Milani[27], Philip B. Mitchell[175], Gunnar Morken[21,197], Ole Mors[14,198], Preben Bo Mortensen[12,14,38,39], Bryan Mowry[176], Bertram Müller-Myhsok[62,199,200], Richard M. Myers[124], Benjamin M. Neale[25,45,179], Caroline M. Nievergelt[89,201], Merete Nordentoft[14,202], Markus M. Nöthen[3], Michael C. O'Donovan[75], Ketil J. Oedegaard[203,204], Tomas Olsson[205], Michael J. Owen[75], Sara A. Paciga[206], Chris Pantelis[207], Carlos Pato[108], Michele T. Pato[108], George P. Patrinos[110,208,209], Roy H. Perlis[210,211], Danielle Posthuma[212,213], Josep Antoni Ramos-Quiroga[53,54,55,56], Andreas Reif[57], Eva Z. Reininghaus[50], Marta Ribasés[53,54,56,145], Marcella Rietschel[78], Stephan Ripke[25,28,45], Guy A. Rouleau[126,214], Takeo Saito[24], Ulrich Schall[59], Martin Schalling[118,119], Peter R. Schofield[152,153], Thomas G. Schulze[36,78,81,136,215], Laura J. Scott[172], Rodney J. Scott[59], Alessandro Serretti[216], Cynthia Shannon Weickert[152,175,217], Jordan W. Smoller[25,133,218], Hreinn Stefansson[32], Kari Stefansson[32,219], Eystein Stordal[220,221], Fabian Streit[78], Patrick F. Sullivan[20,222,223], Gustavo Turecki[224], Arne E. Vaaler[225], Eduard Vieta[226], John B. Vincent[102], Irwin D. Waldman[227], Thomas W. Weickert[152,175,217], Thomas Werge[14,228,229,230], Naomi R. Wray[11,231], John-Anker Zwart[18,19,33], Joanna M. Biernacka[8,64], John I. Nurnberger[232], Sven Cichon[3,4,95,96], Howard J. Edenberg[77,233], Eli A. Stahl[1,2,179,236], Andrew McQuillin[40,236], Arianna Di Florio[75,223,236], Roel A. Ophoff[23,79,117,234,236] and Ole A. Andreassen[6,7,236†]

**PGC BIP Affiliations**

1 Department of Genetics and Genomic Sciences, Icahn School of Medicine at Mount Sinai, New York, NY, USA. 2 Department of Psychiatry, Icahn School of Medicine at Mount Sinai, New York, NY, USA. 3 Institute of Human Genetics, University of Bonn, School of Medicine and University Hospital Bonn, Bonn, Germany. 4 Institute of Neuroscience and Medicine (INM-1), Research Centre Jülich, Jülich, Germany. 5 Centre for Human Genetics, University of Marburg, Marburg, Germany. 6 Division of Mental Health and Addiction, Oslo University Hospital, Oslo, Norway. 7 NORMENT, University of Oslo, Oslo, Norway. 8 Department of Health Sciences Research, Mayo Clinic, Rochester, MN, USA. 9 Social, Genetic and Developmental Psychiatry Centre, King's College London, London, UK. 10 NIHR Maudsley BRC, King's College London, London, UK. 11 Institute for Molecular Bioscience, The University of Queensland, Brisbane, Queensland, Australia. 12 iSEQ, Center for Integrative Sequencing, Aarhus University, Aarhus, Denmark. 13 Department of Biomedicine – Human Genetics, Aarhus University, Aarhus, Denmark. 14 iPSYCH, The Lundbeck Foundation Initiative for Integrative Psychiatric Research, Aarhus, Denmark. 15 Department of Psychiatry and Behavioral Sciences, SUNY Downstate Health Sciences University, Brooklyn, NY, USA. 16 VA NY Harbor Healthcare System, Brooklyn, NY, USA. 17 Research and Communication Unit for Musculoskeletal Health, Division of Clinical Neuroscience, Oslo University Hospital, Oslo, Norway. 18 Institute of Clinical Medicine, University of Oslo, Oslo, Norway. 19 K. G. Jebsen Center for Genetic Epidemiology, Department of Public Health and Nursing, Faculty of Medicine and Health Sciences, Norwegian University of Science and Technology, Trondheim, Norway. 20 Department of Medical Epidemiology and Biostatistics, Karolinska Institutet, Stockholm, Sweden. 21 Department of Mental Health, Faculty of Medicine and Health Sciences, Norwegian University of Science and Technology (NTNU), Trondheim, Norway. 22 Department of Østmarka, Division of Mental Health Care, St Olavs Hospital, Trondheim University Hospital, Trondheim, Norway. 23 Department of Psychiatry and Biobehavioral Science, Semel Institute, David Geffen School of Medicine, University of California, Los Angeles, Los Angeles, CA, USA. 24 Department of Psychiatry, School of Medicine, Fujita Health University, Toyoake, Japan. 25 Stanley Center for Psychiatric Research, Broad Institute, Cambridge, MA, USA. 26 Department of Genetics, Harvard Medical School, Boston, MA, USA. 27 Estonian Genome Center, Institute of Genomics, University of Tartu, Tartu, Estonia. 28 Department of Psychiatry and Psychotherapy, Charité - Universitätsmedizin, Berlin, Germany. 29 Department of Neuroscience, Icahn School of Medicine at Mount Sinai, New York, NY, USA. 30 Ronald M. Loeb Center for Alzheimer's Disease, Icahn School of Medicine at Mount Sinai, New York, NY, USA. 31 Estelle and

Daniel Maggin Department of Neurology, Icahn School of Medicine at Mount Sinai, New York, NY, USA. 32 deCODE Genetics/Amgen, Reykjavik, Iceland. 33 Department of Research, Innovation and Education, Division of Clinical Neuroscience, Oslo University Hospital, Oslo, Norway. 34 Samsung Advanced Institute for Health Sciences and Technology (SAIHST), Samsung Medical Center, Sungkyunkwan University, Seoul, South Korea. 35 Russian Academy of Medical Sciences, Mental Health Research Center, Moscow, Russian Federation. 36 Institute of Psychiatric Phenomics and Genomics (IPPG), University Hospital, LMU Munich, Munich, Germany. 37 Department of Psychiatry and Psychotherapy, University Hospital, LMU Munich, Munich, Germany. 38 National Centre for Register-Based Research, Aarhus University, Aarhus, Denmark. 39 Centre for Integrated Register-Based Research, Aarhus University, Aarhus, Denmark. 40 Division of Psychiatry, University College London, London, UK. 41 Department of Neuroscience, Istituto Di Ricerche Farmacologiche Mario Negri IRCCS, Milan, Italy. 42 Department of Psychiatry and Behavioral Neuroscience, University of Chicago, Chicago, IL, USA. 43 Northwestern University, Chicago, IL, USA. 44 Psychiatry, Berkshire Healthcare NHS Foundation Trust, Bracknell, UK. 45 Analytic and Translational Genetics Unit, Massachusetts General Hospital, Boston, MA, USA. 46 2nd Department of Psychiatry, Attikon General Hospital, National and Kapodistrian University of Athens, Athens, Greece. 47 Department of Psychiatry, Samsung Medical Center, School of Medicine, Sungkyunkwan University, Seoul, South Korea. 48 Center for Neonatal Screening, Department for Congenital Disorders, Statens Serum Institut, Copenhagen, Denmark. 49 Department of Psychiatry and Psychotherapy, University Hospital Carl Gustav Carus, Technische Universität Dresden, Dresden, Germany. 50 Department of Psychiatry and Psychotherapeutic Medicine, Medical University of Graz, Graz, Austria. 51 Department of Psychiatric Research, Diakonhjemmet Hospital, Oslo, Norway. 52 Psychiatry, Brain Center UMC Utrecht, Utrecht, the Netherlands. 53 Instituto de Salud Carlos III, Biomedical Network Research Centre on Mental Health (CIBERSAM), Madrid, Spain. 54 Department of Psychiatry, Hospital Universitari Vall d´Hebron, Barcelona, Spain. 55 Department of Psychiatry and Forensic Medicine, Universitat Autònoma de Barcelona, Barcelona, Spain. 56 Psychiatric Genetics Unit, Group of Psychiatry Mental Health and Addictions, Vall d´Hebron Research Institut (VHIR), Universitat Autònoma de Barcelona, Barcelona, Spain. 57 Department of Psychiatry, Psychosomatic Medicine and Psychotherapy, University Hospital Frankfurt, Frankfurt am Main, Germany. 58 Psychiatry, University of California San Francisco, San Francisco, CA, USA. 59 University of Newcastle, Newcastle, New South Wales, Australia. 60 Mood Disorders Program, Department of Psychiatry, McGill University Health Center, Montreal, Quebec, Canada. 61 Division of Psychiatry, University of Edinburgh, Edinburgh, UK. 62 Department of Translational Research in Psychiatry, Max Planck Institute of Psychiatry, Munich, Germany. 63 Department of Psychiatry, Universidad Autonoma de Nuevo Leon, Monterrey, Mexico. 64 Department of Psychiatry and Psychology, Mayo Clinic, Rochester, MN, USA. 65 Department of Laboratory Medicine and Pathology, Mayo Clinic, Rochester, MN, USA. 66 Centre for Psychiatry, Queen Mary University of London, London, UK. 67 UCL Genetics Institute, University College London, London, UK. 68 Department of Psychiatry, Laboratory of Psychiatric Genetics, Poznan University of Medical Sciences, Poznan, Poland. 69 Center for Multimodal Imaging and Genetics, Departments of Neurosciences, Radiology, and Psychiatry, University of California, San Diego, CA, USA. 70 Department of Child and Adolescent Psychiatry, Psychosomatics and Psychotherapy, University Hospital Essen, University of Duisburg-Essen, Duisburg, Germany. 71 Department of Medical Genetics, Oslo University Hospital, Oslo, Norway. 72 NORMENT, Department of Clinical Science, University of Bergen, Bergen, Norway. 73 Department of Neurology, Oslo University Hospital, Oslo, Norway. 74 NORMENT, KG Jebsen Centre for Psychosis Research, Oslo University Hospital, Oslo, Norway. 75 Medical Research Council Centre for Neuropsychiatric Genetics and Genomics, Division of Psychological Medicine and Clinical Neurosciences, Cardiff University, Cardiff, UK. 76 Academic Psychiatry, Newcastle University, Newcastle upon Tyne, UK. 77 Department of Medical and Molecular Genetics, Indiana University, Indianapolis, IN, USA. 78 Department of Genetic Epidemiology in Psychiatry, Central Institute of Mental Health, Medical Faculty Mannheim, Heidelberg University, Mannheim, Germany. 79 Center for Neurobehavioral Genetics, Semel Institute for Neuroscience and Human Behavior, Los Angeles,

CA, USA. 80 Department of Clinical Neuroscience, Karolinska Institutet, Stockholm, Sweden. 81 Department of Psychiatry and Psychotherapy, University Medical Center Göttingen, Göttingen, Germany. 82 Department of Psychiatry, Dalhousie University, Halifax, Nova Scotia, Canada. 83 Department of Psychiatry, Yale School of Medicine, New Haven, CT, USA. 84 Veterans Affairs Connecticut Healthcare System, West Haven, CT, USA. 85 Departments of Genetics and Neuroscience, Yale University School of Medicine, New Haven, CT, USA. 86 Department of Psychological Sciences, University of Missouri, Columbia, MO, USA. 87 Genetics and Computational Biology, QIMR Berghofer Medical Research Institute, Brisbane, Queensland, Australia. 88 Psychological Medicine, University of Worcester, Worcester, UK. 89 Department of Psychiatry, University of California San Diego, La Jolla, CA, USA. 90 Bioinformatics Research Centre, Aarhus University, Aarhus, Denmark. 91 Mental Health Department, University Regional Hospital, Biomedicine Institute (IBIMA), Málaga, Spain. 92 Department of Psychiatry, Seoul National University College of Medicine, Seoul, South Korea. 93 Landspitali University Hospital, Reykjavik, Iceland. 94 Department of Psychology, Eberhard Karls Universität Tübingen, Tübingen, Germany. 95 Department of Biomedicine, University of Basel, Basel, Switzerland. 96 Institute of Medical Genetics and Pathology, University Hospital Basel, Basel, Switzerland. 97 Neuropsychiatrie Translationnelle, Inserm U955, Créteil, France. 98 Faculté de Santé, Université Paris Est, Créteil, France. 99 International Max Planck Research School for Translational Psychiatry (IMPRS-TP), Munich, Germany. 100 Laboratory of Complex Trait Genomics, Department of Computational Biology and Medical Sciences, Graduate School of Frontier Sciences, The University of Tokyo, Tokyo, Japan. 101 Laboratory for Statistical and Translational Genetics, RIKEN Center for Integrative Medical Sciences, Yokohama, Japan. 102 Campbell Family Mental Health Research Institute, Centre for Addiction and Mental Health, Toronto, Ontario, Canada. 103 Neurogenetics Section, Centre for Addiction and Mental Health, Toronto, Ontario, Canada. 104 Department of Psychiatry, University of Toronto, Toronto, Ontario, Canada. 105 Institute of Medical Sciences, University of Toronto, Toronto, Ontario, Canada. 106 Department of Psychiatry, Psychosomatics and Psychotherapy, Center of Mental Health, University Hospital Würzburg, Würzburg, Germany. 107 Cell Biology, SUNY Downstate Medical Center College of Medicine, Brooklyn, NY, USA. 108 Institute for Genomic Health, SUNY Downstate Medical Center College of Medicine, Brooklyn, NY, USA. 109 ISGlobal, Barcelona, Spain. 110 Laboratory of Pharmacogenomics and Individualized Therapy, Department of Pharmacy, School of Health Sciences, University of Patras, Patras, Greece. 111 Mental Illness Research, Education and Clinical Center, Crescenz VAMC, Philadelphia, PA, USA. 112 Center for Studies of Addiction, University of Pennsylvania Perelman School of Medicine, Philadelphia, PA, USA. 113 RIKEN Center for Integrative Medical Sciences, Yokohama, Japan. 114 Psychiatry, Altrecht, Utrecht, the Netherlands. 115 Psychiatry, GGZ inGeest, Amsterdam, the Netherlands. 116 Psychiatry, VU Medisch Centrum, Amsterdam, the Netherlands. 117 Department of Psychiatry, Erasmus MC, University Medical Center Rotterdam, Rotterdam, the Netherlands. 118 Department of Molecular Medicine and Surgery, Karolinska Institutet, Stockholm, Sweden. 119 Center for Molecular Medicine, Karolinska University Hospital, Stockholm, Sweden. 120 Psychiatry, North East London NHS Foundation Trust, Ilford, UK. 121 Clinic for Psychiatry and Psychotherapy, University Hospital Cologne, Cologne, Germany. 122 Department of Psychiatry, Korea University College of Medicine, Seoul, South Korea. 123 Psychiatric and Neurodevelopmental Genetics Unit, Center for Genomic Medicine, Massachusetts General Hospital and Harvard Medical School, Boston, MA, USA. 124 HudsonAlpha Institute for Biotechnology, Huntsville, AL, USA. 125 Department of Human Genetics, McGill University, Montréal, Quebec, Canada. 126 Montreal Neurological Institute and Hospital, McGill University, Montréal, Quebec, Canada. 127 Division of Psychiatry, Centre for Clinical Brain Sciences, The University of Edinburgh, Edinburgh, UK. 128 Department of Psychiatry and Psychotherapy, University of Bonn, Bonn, Germany. 129 Clinical Biochemistry Laboratory, Attikon General Hospital, Medical School, National and Kapodistrian University of Athens, Athens, Greece. 130 Department of Clinical Neuroscience, Centre for Psychiatry Research, Karolinska Institutet, Stockholm, Sweden. 131 Systems Genetics Working Group, Department of Genetics, Stellenbosch University, Stellenbosch, South Africa. 132 Genetic Cancer Susceptibility Group, International Agency for Research on Cancer,

Lyon, France. 133 Department of Psychiatry, Massachusetts General Hospital, Boston, MA, USA. 134 Centre for Neuroimaging and Cognitive Genomics (NICOG), National University of Ireland Galway, Galway, Ireland. 135 Medical Faculty, School of Science and Technology, University Sarajevo, Sarajevo, Bosnia and Herzegovina. 136 Department of Psychiatry and Behavioral Sciences, Johns Hopkins University School of Medicine, Baltimore, MD, USA. 137 Oxford Health NHS Foundation Trust, Warneford Hospital, Oxford, UK. 138 Department of Psychiatry, University of Oxford, Warneford Hospital, Oxford, UK. 139 Department of Psychiatry and Behavioral Sciences, Emory University School of Medicine, Atlanta, GA, USA. 140 Outpatient Clinic for Bipolar Disorder, Altrecht, Utrecht, the Netherlands. 141 Department of Psychiatry, Washington University in Saint Louis, Saint Louis, MO, USA. 142 Department of Biochemistry and Molecular Biology II, Faculty of Pharmacy, University of Granada, Granada, Spain. 143 Institute of Neurosciences, Biomedical Research Center (CIBM), University of Granada, Granada, Spain. 144 Medicine, Psychiatry, Biomedical Informatics, Vanderbilt University Medical Center, Nashville, TN, USA. 145 Department of Genetics, Microbiology and Statistics, Faculty of Biology, Universitat de Barcelona, Barcelona, Spain. 146 Faculty of Medicine, Department of Psychiatry, School of Health Sciences, University of Iceland, Reykjavik, Iceland. 147 Institute of Health and Wellbeing, University of Glasgow, Glasgow, UK. 148 Psychiatry and the Behavioral Sciences, University of Southern California, Los Angeles, CA, USA. 149 Mood Disorders, PsyQ, Rotterdam, the Netherlands. 150 SAMRC Unit on Risk and Resilience in Mental Disorders, Department of Psychiatry and Neuroscience Institute, University of Cape Town, Cape Town, South Africa. 151 Department of Environmental Epidemiology, Nofer Institute of Occupational Medicine, Lodz, Poland. 152 Neuroscience Research Australia, Sydney, New South Wales, Australia. 153 School of Medical Sciences, University of New South Wales, Sydney, New South Wales, Australia. 154 Centro de Biología Molecular Severo Ochoa, Universidad Autónoma de Madrid and CSIC, Madrid, Spain. 155 Department of Psychiatry and Human Behavior, School of Medicine, University of California, Irvine, Irvine, CA, USA. 156 Psychiatry, Psychiatrisches Zentrum Nordbaden, Wiesloch, Germany. 157 Computational Sciences Center of Emphasis, Pfizer Global Research and Development, Cambridge, MA, USA. 158 Dalla Lana School of Public Health, University of Toronto, Toronto, Ontario, Canada. 159 Department of Psychological Medicine, Institute of Psychiatry, Psychology and Neuroscience, King's College London, London, UK. 160 South London and Maudsley NHS Foundation Trust, Bethlem Royal Hospital, Beckenham, UK. 161 Department of Clinical Sciences, Psychiatry, Umeå University Medical Faculty, Umeå, Sweden. 162 NORMENT, KG Jebsen Centre for Psychosis Research, Division of Mental Health and Addiction, Institute of Clinical Medicine and Diakonhjemmet Hospital, University of Oslo, Oslo, Norway. 163 National Institute of Mental Health, Klecany, Czech Republic. 164 Institute of Environmental Medicine, Karolinska Institutet, Stockholm, Sweden. 165 Institute of Pulmonology, Russian State Medical University, Moscow, Russian Federation. 166 Department of Psychiatry, University of Münster, Münster, Germany. 167 Department of Psychiatry, Melbourne Medical School, The University of Melbourne, Melbourne, Victoria, Australia. 168 The Florey Institute of Neuroscience and Mental Health, The University of Melbourne, Parkville, Victoria, Australia. 169 Université de Paris, INSERM, Optimisation Thérapeutique en Neuropsychopharmacologie, UMRS 1144, Paris, France. 170 APHP Nord, DMU Neurosciences, Département de Psychiatrie et de Médecine Addictologique, GHU Saint Louis-Lariboisière-Fernand Widal, Paris, France. 171 Psychiatry, University of Pennsylvania, Philadelphia, PA, USA. 172 Center for Statistical Genetics and Department of Biostatistics, University of Michigan, Ann Arbor, MI, USA. 173 Department of Biomedicine and the iSEQ Center, Aarhus University, Aarhus, Denmark. 174 Center for Genomics and Personalized Medicine, CGPM, Aarhus, Denmark. 175 School of Psychiatry, University of New South Wales, Sydney, New South Wales, Australia. 176 University of Queensland, Brisbane, Queensland, Australia. 177 Neuropsychiatric Genetics Research Group, Department of Psychiatry and Trinity Translational Medicine Institute, Trinity College Dublin, Dublin, Ireland. 178 1st Department of Psychiatry, Eginition Hospital, National and Kapodistrian University of Athens, Athens, Greece. 179 Medical and Population Genetics, Broad Institute, Cambridge, MA, USA. 180 Division of Endocrinology, Children's Hospital Boston, Boston, MA, USA. 181 Department of Human Genetics, University of Chicago, Chicago, IL, USA. 182 Biometric

Psychiatric Genetics Research Unit, Alexandru Obregia Clinical Psychiatric Hospital, Bucharest, Romania. 183 HUNT Research Center, Department of Public Health and Nursing, Faculty of Medicine and Health Sciences, Norwegian University of Science and Technology, Trondheim, Norway. 184 University of Western Australia, Nedlands, Western Australia, Australia. 185 Institute of Neuroscience and Physiology, University of Gothenburg, Gothenburg, Sweden. 186 Department of Psychiatry and Addiction Medicine, Assistance Publique - Hôpitaux de Paris, Paris, France. 187 Department of Medical and Molecular Genetics, King's College London, London, UK. 188 Neuroscience Therapeutic Area, Janssen Research and Development, LLC, Titusville, NJ, USA. 189 Cancer Epidemiology and Prevention, M. Sklodowska-Curie National Research Institute of Oncology, Warsaw, Poland. 190 SA MRC Unit on Risk and Resilience in Mental Disorders, Department of Psychiatry, Stellenbosch University, Stellenbosch, South Africa. 191 School of Psychology, The University of Queensland, Brisbane, Queensland, Australia. 192 Department of Psychiatry and Genetics Institute, University of Florida, Gainesville, FL, USA. 193 Research Institute, Lindner Center of HOPE, Mason, OH, USA. 194 Centre for Cognitive Ageing and Cognitive Epidemiology, University of Edinburgh, Edinburgh, UK. 195 Human Genetics Branch, Intramural Research Program, National Institute of Mental Health, Bethesda, MD, USA. 196 Division of Mental Health and Addiction, University of Oslo, Institute of Clinical Medicine, Oslo, Norway. 197 Psychiatry, St Olavs University Hospital, Trondheim, Norway. 198 Psychosis Research Unit, Aarhus University Hospital - Psychiatry, Risskov, Denmark. 199 Munich Cluster for Systems Neurology (SyNergy), Munich, Germany. 200 University of Liverpool, Liverpool, UK. 201 Research/Psychiatry, Veterans Affairs San Diego Healthcare System, San Diego, CA, USA. 202 Mental Health Services in the Capital Region of Denmark, Mental Health Center Copenhagen, University of Copenhagen, Copenhagen, Denmark. 203 Division of Psychiatry, Haukeland Universitetssjukehus, Bergen, Norway. 204 Faculty of Medicine and Dentistry, University of Bergen, Bergen, Norway. 205 Department of Clinical Neuroscience and Center for Molecular Medicine, Karolinska Institutet at Karolinska University Hospital, Solna, Sweden. 206 Human Genetics and Computational Biomedicine, Pfizer Global Research and Development, Groton, CT, USA. 207 University of Melbourne, Melbourne, Victoria, Australia. 208 Department of Pathology, College of Medicine and Health Sciences, United Arab Emirates University, Al-Ain, United Arab Emirates. 209 Zayed Center of Health Sciences, United Arab Emirates University, Al-Ain, United Arab Emirates. 210 Psychiatry, Harvard Medical School, Boston, MA, USA. 211 Division of Clinical Research, Massachusetts General Hospital, Boston, MA, USA. 212 Department of Complex Trait Genetics, Center for Neurogenomics and Cognitive Research, Amsterdam Neuroscience, Vrije Universiteit Amsterdam, Amsterdam, the Netherlands. 213 Department of Clinical Genetics, Amsterdam Neuroscience, Vrije Universiteit Medical Center, Amsterdam, the Netherlands. 214 Department of Neurology and Neurosurgery, Faculty of Medicine, McGill University, Montreal, Quebec, Canada. 215 Department of Psychiatry and Behavioral Sciences, SUNY Upstate Medical University, Syracuse, NY, USA. 216 Department of Biomedical and NeuroMotor Sciences, University of Bologna, Bologna, Italy. 217 Department of Neuroscience, SUNY Upstate Medical University, Syracuse, NY, USA. 218 Psychiatric and Neurodevelopmental Genetics Unit (PNGU), Massachusetts General Hospital, Boston, MA, USA. 219 Faculty of Medicine, University of Iceland, Reykjavik, Iceland. 220 Department of Psychiatry, Hospital Namsos, Namsos, Norway. 221 Department of Neuroscience, Norges Teknisk Naturvitenskapelige Universitet Fakultet for Naturvitenskap og Teknologi, Trondheim, Norway. 222 Department of Genetics, University of North Carolina at Chapel Hill, Chapel Hill, NC, USA. 223 Department of Psychiatry, University of North Carolina at Chapel Hill, Chapel Hill, NC, USA. 224 Department of Psychiatry, McGill University, Montreal, Quebec, Canada. 225 Department of Psychiatry, Sankt Olavs Hospital Universitetssykehuset i Trondheim, Trondheim, Norway. 226 Clinical Institute of Neuroscience, Hospital Clinic, University of Barcelona, IDIBAPS, CIBERSAM, Barcelona, Spain. 227 Department of Psychology, Emory University, Atlanta, GA, USA. 228 Institute of Biological Psychiatry, Mental Health Services, Copenhagen University Hospital, Copenhagen, Denmark. 229 Department of Clinical Medicine, University of Copenhagen, Copenhagen, Denmark. 230 Center for GeoGenetics, GLOBE Institute, University of Copenhagen, Copenhagen, Denmark. 231 Queensland Brain Institute, The University of Queensland,

Brisbane, Queensland, Australia. 232 Psychiatry, Indiana University School of Medicine, Indianapolis, IN, USA. 233 Biochemistry and Molecular Biology, Indiana University School of Medicine, Indianapolis, IN, USA. 234 Department of Human Genetics, David Geffen School of Medicine, University of California Los Angeles, Los Angeles, CA, USA. 235 These authors contributed equally: Niamh Mullins, Andreas J. Forstner. 236 These authors jointly supervised this work: Eli A. Stahl, Andrew McQuillin, Arianna Di Florio, Roel A. Ophoff, Ole A. Andreassen.

### List of PGC MDD collaborators

**PGC MDD Collaborators**

Mark J Adams 1 *
Fabian Streit 2 *
Swapnil Awasthi 3 *
Brett N Adey 4
Karmel W Choi 5, 6
V Kartik Chundru 7
Jonathan RI Coleman 4, 8
Jerome C Foo 2
Olga Giannakopoulou 9
Alisha S M Hall 2, 10
Jens Hjerling-Leffler 11
David M Howard 4
Christopher Hübel 4, 12, 13
Alex S F Kwong 1, 14
Bochao Danae Lin 15
Xiangrui Meng 9
Guiyan Ni 16
Oliver Pain 17
Gita A Pathak 18, 19
Eva C Schulte 20, 21, 22, 23
Jackson G Thorp 24
Alicia Walker 16
Shuyang Yao 25
Jian Zeng 16
Johan Zvrskovec 4, 8
Dag Aarsland 26
Ky'Era V Actkins 27
Mazda Adli 3, 28
Esben Agerbo 12, 29, 30
Mareike Aichholzer 31
Tracy M Air 32
Allison Aiello 33
Thomas D Als 30, 34, 35
Evelyn Andersson 36
Till F M Andlauer 37, 38
Volker Arolt 39
Helga Ask 40, 41
Sunita Badola 42
Clive Ballard 43
Karina Banasik 44
Nicholas J Bass 9
Aartjan T F Beekman 45
Sintia Belangero 46
Elisabeth B Binder 38, 47
Ottar Bjerkeset 48, 49
Gyda Bjornsdottir 50
Julia Boberg 36
Sigrid Børte 51, 52, 53
Emma Bränn 54
Alice Braun 55
Thorsten Brodersen 56
Søren Brunak 44
Mie T Bruun 57
Pichit Buspavanich 58, 59
Jonas Bybjerg-Grauholm 60, 61
Enda M Byrne 62
Archie Campbell 63, 64
Megan L. Campbell 65
Enrique Castelao 66
Jorge Cervilla 67, 68
Boris Chaumette 69
Chia-Yen Chen 70
Zhengming Chen 71, 72
Sven Cichon 73, 74, 75, 76
Lucía Colodro-Conde 24
Anne Corbett 43
Elizabeth C Corfield 40, 77
Baptiste Couvy-Duchesne 78
Nick Craddock 79, 80
Udo Dannlowski 39
Gail Davies 81
EJC de Geus 82
Ian J Deary 81
Franziska Degenhardt 76, 83
Abbas Dehghan 84, 85
J Raymond DePaulo 86
Michael Deuschle 87
Maria Didriksen 88
Khoa Manh Dinh 89
Nese Direk 90
Srdjan Djurovic 91, 92
Anna R Docherty 93, 94, 95
Katharina Domschke 96
Joseph Dowsett 88
Ole Kristian Drange 49, 97, 98, 99
Erin C Dunn 6, 100
Gudmundur Einarsson 50
Thalia C Eley 4
Samar S M Elsheikh 101
Jan Engelmann 102
Michael E Benros 60, 103, 104
Christian Erikstrup 89
Valentina Escott-Price 80
Chiara Fabbri 4, 105
Yu Fang 106
Sarah Finer 107
Josef Frank 2
Robert C Free 108
He Gao 109
Michael Gill 110
Maria Gilles 87
Fernando S Goes 86
Scott Douglas Gordon 24
Jakob Grove 30, 34, 35, 111
Daniel F Gudbjartsson 50, 112
Blanca Gutierrez 67, 68
Tim Hahn 39
Lynsey S Hall 80
Thomas F Hansen 44, 60, 113
Magnus Haraldsson 114
Catherina A Hartman 115
Alexandra Havdahl 40
Caroline Hayward 116
Stefanie Heilmann-Heimbach 76
Stefan Herms 74, 76
Ian B Hickie 117
Henrik Hjalgrim 118
Per Hoffmann 74, 76
Georg Homuth 119
Carsten Horn 120
Jouke-Jan Hottenga 82
David M Hougaard 60, 61
Iiris Hovatta 121
Qin Qin Huang 7
Floris Huider 82
Karen A Hunt 122
Marcus Ising 123
Erkki Isometsä 124
Rick Jansen 45
Yunxuan Jiang 125
Ian Jones 80
Lisa A Jones 126
Lina Jonsson 127
Robert Karlsson 25
Siegfried Kasper 128
Kenneth S Kendler 129
Ronald C Kessler 130
Stefan Kloiber 101, 123, 131, 132
James A Knowles 133
Nastassja Koen 65

Julia Kraft 55
Henry R Kranzler 134, 135
Kristi Krebs 136
Theodora Kunovac Kallak 137
Zoltán Kutalik 138, 139, 140
Elisa Lahtela 141
Margit Hørup Larsen 88
Eric J Lenze 142
Daniel F Levey 143, 144
Melissa Lewins 1
Glyn Lewis 9
Liming Li 145, 146
Kuang Lin 71
Penelope A Lind 24
Donald J MacIntyre 1, 147, 148
Dean F MacKinnon 86
Hermine HM Maes 149, 150
Wolfgang Maier 151
Victoria S Marshe 101, 152
Hamdi Mbarek 82
Peter McGuffin 4
Sarah E Medland 24
Susanne Meinert 39, 153
Susan Mikkelsen 89
Christina Mikkelsen 88, 154
Yuri Milaneschi 45
Iona Y Millwood 71, 72
Brittany L Mitchell 24
Esther Molina 67, 155
Francis M Mondimore 86
Preben Bo Mortensen 12, 29, 30
Benoit H Mulsant 101, 131
Joonas Naamanka 121
Jake M Najman 156
Matthias Nauck 157, 158
Igor Nenadić 159
Kasper R Nielsen 160
Ilja M Nolte 161
Merete Nordentoft 60, 103, 104
Markus M Nöthen 76
Mette Nyegaard 30, 162, 163, 164
Michael C O'Donovan 80
Asmundur Oddsson 50
Catherine M Olsen 165, 166
Hogni Oskarsson 167
Sisse Rye Ostrowski 88, 168
Vanessa K Ota 46
Michael J Owen 80
Richard Packer 169
Teemu Palviainen 141
Pedro M Pan 170
Carlos N Pato 171
Michele T Pato 171
Nancy L Pedersen 25
Ole Birger Pedersen 172
Roseann E Peterson 129, 173
Wouter J Peyrot 45
James B Potash 86
Martin Preisig 66
Jorge A Quiroz 174
Charles F Reynolds III 175
John P Rice 142
Giovanni A Salum 176
Robert A Schoevers 177, 178
Andrew Schork 30, 179, 180
Thomas G Schulze 2, 21, 86, 181, 182
Tabea S Send 87
Jianxin Shi 183
Engilbert Sigurdsson 114
Kritika Singh 27
Grant C B Sinnamon 184
Lea Sirignano 2
Olav B Smeland 185, 186
Daniel J Smith 187
Erik Sørensen 88
Sundararajan Srinivasan 188
Hreinn Stefansson 50
Kari Stefansson 50, 189
Dan J. Stein 190
Frederike Stein 191
André Tadic 102, 192
Henning Teismann 193
Alexander Teumer 194
Anita Thapar 80, 195
Pippa A Thomson 64
Lise Wegner Thørner 88
Apostolia Topaloudi 196
Ioanna Tzoulaki 84, 85, 197
Monica Uddin 198
André G Uitterlinden 199
Henrik Ullum 88, 200, 201
Daniel Umbricht 202
Robert J Ursano 203
Sandra Van der Auwera 204
David A van Heel 122
Albert M van Hemert 205
Abirami Veluchamy 188
Alexander Viktorin 25
Henry Völzke 194
Agaz Wani 198
G Bragi Walters 50
Robin G Walters 71, 72
Sylvia Wassertheil-Smoller 206
Myrna M Weissman 207, 208
Jürgen Wellmann 193
David C Whiteman 165
Derek Wildman 198
Gonneke Willemsen 82
Alexander T Williams 169
Bendik S Winsvold 51, 52, 209
Stephanie H Witt 2
Ying Xiong 25
Lea Zillich 2
John-Anker Zwart 51, 52, 53
23andMe Research Team 125
Estonian Biobank Research Team 136
HUNT All-In Psychiatry 210
China Kadoorie Biobank Collaborative Group 211
Genes & Health Research Team 212
Ole A Andreassen 185, 186, 213
Bernhard T Baune 214, 215, 216
Klaus Berger 193
Dorret I Boomsma 82
Anders D Børglum 30, 34, 35
Gerome Breen 4, 8
Na Cai 217, 218, 219
Hilary Coon 94
William E Copeland 220
Byron Creese 43
Lea K Davis 27
Eske M Derks 24
Enrico Domenici 221
Paul Elliott 84, 85, 197, 222
Andreas J Forstner 73, 76
Micha Gawlik 223
Joel Gelernter 19, 143, 224
Hans J Grabe 204

Steven P Hamilton 225
Kristian Hveem 226, 227, 228
Catherine John 169, 229
Jaakko Kaprio 141
Tilo Kircher 159
Marie-Odile Krebs 230
Karoline Kuchenbaecker 9, 71
Mikael Landén 25, 127
Kelli Lehto 136
Douglas F Levinson 231
Qingqin S Li 232
Klaus Lieb 102
Yi Lu 25
Susanne Lucae 123
Jurjen J Luykx 15, 233
Patrik K Magnusson 25
Nicholas G Martin 24
Hilary C Martin 7
Andrew McQuillin 9
Christel M Middeldorp 62, 234
Lili Milani 136
Ole Mors 30, 235
Daniel J Müller 101, 131, 132, 236
Bertram Müller-Myhsok 38, 237, 238
Albertine J Oldehinkel 115
Sara A Paciga 239
Colin NA Palmer 188
Peristera Paschou 196
Brenda WJH Penninx 45
Roy H Perlis 5, 6, 240
Giorgio Pistis 66
Renato Polimanti 18, 19
David J Porteous 64
Danielle Posthuma 241, 242
Ted Reichborn-Kjennerud 40
Andreas Reif 31
Frances Rice 80, 243
Roland Ricken 3
Marcella Rietschel 2
Margarita Rivera 67, 244
Christian Rück 245
Catherine Schaefer 246
Srijan Sen 106, 247
Alessandro Serretti 105
Alkistis Skalkidou 137
Jordan W Smoller 5, 248, 249
Frederike Stein 191
Murray B Stein 250, 251, 25
Patrick F Sullivan 25, 253
Martin Tesli 40
Thorgeir E Thorgeirsson 50
Henning Tiemeier 254, 255
Nicholas J Timpson 14
Rudolf Uher 256
Jens R Wendland 42
Thomas Werge 60, 179, 201, 257, 258
Naomi R Wray 16, 259 **
Stephan Ripke 3, 248 **
Cathryn M Lewis 4, 260 **
Andrew M McIntosh 1, 261 **

* Joint Lead Authors
** Joint Last Authors

**PGC MDD Affiliations**

1, Division of Psychiatry, University of Edinburgh, Edinburgh, UK
2, Department of Genetic Epidemiology in Psychiatry, Central Institute of Mental Health, Medical Faculty Mannheim, Heidelberg University, Mannheim, BW, DE
3, Department of Psychiatry and Psychotherapy, Charité – Universitätsmedizin Berlin, Berlin, BE, DE
4, Social, Genetic and Developmental Psychiatry Centre, King's College London, London, UK
5, Department of Psychiatry, Massachusetts General Hospital, Boston, MA, US
6, Department of Psychiatry, Harvard Medical School, Boston, MA, US
7, Human Genetics, Wellcome Sanger Institute, Hinxton, UK
8, NIHR Maudsley Biomedical Research Centre, King's College London, London, UK
9, Division of Psychiatry, University College London, London, UK
10, Department of Clinical Medicine, Aarhus University, Aarhus, DK
11, Department of Medical Biochemistry and Biophysics, Karolinska Institutet, Stockholm, SE
12, National Centre for Register-based Research, Aarhus University, Aarhus, DK
13, Department of Pediatric Neurology, Charité – Universitätsmedizin Berlin, Berlin, BE, DE
14, MRC Integrative Epidemiology Unit, University of Bristol, Bristol, UK
15, Department of Psychiatry and Neuropsychology, School for Mental Health and Neuroscience, Maastricht University Medical Centre, Maastricht, NL
16, Institute for Molecular Bioscience, University of Queensland, Brisbane, QLD, AU
17, Maurice Wohl Clinical Neuroscience Institute, Department of Basic and Clinical Neuroscience, King's College London, London, UK
18, Veterans Affairs Connecticut Healthcare System, West Haven, CT, US
19, Department of Psychiatry, Yale University School of Medicine, New Haven, CT, US
20, Department of Psychiatry, University of Munich, Munich, BY, DE
21, Institute of Psychiatric Phenomics and Genomics, University of Munich, Munich, BY, DE
22, Department of Psychiatry and Psychotherapy, University Hospital Bonn, Medical Faculty, University of Bonn, Bonn, DE
23, Institute of Human Genetics, University Hospital Bonn, Medical Faculty, University of Bonn, Bonn, DE
24, Mental Health and Neuroscience, QIMR Berghofer Medical Research Institute, Brisbane, QLD, AU
25, Department of Medical Epidemiology and Biostatistics, Karolinska Institutet, Stockholm, SE
26, Old Age Psychiatry, King's College London, London, UK
27, Department of Medicine, Division of Genetic Medicine, Vanderbilt University Medical Center, Nashville, TN, US
28, Department of Psychiatry and Psychotherapy, Fliedner Klinik Berlin, Berlin, BE, DE
29, Centre for Integrated Register-based Research, Aarhus University, Aarhus, DK
30, iPSYCH, The Lundbeck Foundation Initiative for Integrative Psychiatric Research, Aarhus, DK
31, Department of Psychiatry, Psychosomatic Medicine and Psychotherapy, Goethe University Frankfurt - University Hospital, Frankfurt am Main, DE
32, Discipline of Psychiatry, University of Adelaide, Adelaide, SA, AU
33, Department of Epidemiology, Columbia University Mailman School of Public Health, New York, NY, US
34, Department of Biomedicine and Centre for Integrative Sequencing, iSEQ, Aarhus University, Aarhus, DK
35, Center for Genomics and Personalized Medicine, Aarhus University, Aarhus, DK
36, Department of Clinical Neuroscience, Karolinska Institutet,, SE
37, Department of Neurology, Klinikum rechts der Isar, Technical University of Munich, Munich, BY, DE
38, Department of Translational Research in Psychiatry, Max Planck Institute of Psychiatry, Munich, BY, DE
39, Institute for Translational Psychiatry, University of Münster, Münster, NRW, DE

40, Department of Mental Disorders, Norwegian Institute of Public Health, Oslo, NO
41, PROMENTA Research Center, Department of Psychology, University of Oslo, Oslo, NO
42, Research and Development, Takeda Pharmaceutical Company Limited, Cambridge, MA, US
43, Faculty of Health and Life Sciences, University of Exeter, Exeter, UK
44, Novo Nordisk Center for Protein Research, Department of Health Sciences, University of Copenhagen, Copenhagen, DK
45, Department of Psychiatry, Amsterdam Public Health and Amsterdam Neuroscience, Amsterdam UMC, Vrije Universiteit Amsterdam, Amsterdam, NL
46, Morphology and Genetics, Universidade Federal de Sao Paulo, Sao Paulo, SP, BR
47, Department of Psychiatry and Behavioral Sciences, Emory University School of Medicine, Atlanta, GA, US
48, Faculty of Nursing and Health Sciences, NORD University, Levanger, NO
49, Department of Mental Health, Faculty of Medicine and Health Sciences, Norwegian University of Science and Technology (NTNU), Trondheim, TRD, NO
50, deCODE Genetics / Amgen, Reykjavik, IS
51, K. G. Jebsen Center for Genetic Epidemiology, Department of Public Health and Nursing, Faculty of Medicine and Health Sciences, Norwegian University of Science and Technology (NTNU), Trondheim, TRD, NO
52, Department of Research and Innovation, Division of Clinical Neuroscience, Oslo University Hospital, Oslo, NO
53, Institute of Clinical Medicine, Faculty of Medicine, University of Oslo, Oslo, NO
54, Institute of Environmental Medicine, Unit of Integrative Epidemiology, Karolinska Institutet, Stockholm, SE
55, Department of Psychiatry and Psychotherapy, Charité – Universitätsmedizin Berlin, Berlin, DE
56, Department of Clinical Immunology, Roskilde University/Næstved Hospital, Roskilde, DK
57, Department of Clinical Immunology, Odense University Hospital, Odense, DK
58, Department of Psychiatry, Psychotherapy and Psychosomatics, Brandenburg Medical School Theodor Fontane, Neuruppin, BB, DE
59, Department of Psychiatry and Psychotherapy, Gender Research in Medicine, Institute of Sexology and Sexual Medicine, Charité – Universitätsmedizin Berlin, Berlin, BE, DE
60, iPSYCH, The Lundbeck Foundation Initiative for Integrative Psychiatric Research, Copenhagen, DK
61, Center for Neonatal Screening, Department for Congenital Disorders, Statens Serum Institut, Copenhagen, DK
62, Child Health Research Centre, University of Queensland, Brisbane, QLD, AU
63, Centre for Medical Informatics, Usher Institute, University of Edinburgh, Edinburgh, UK
64, Centre for Genomic & Experimental Medicine, Institute for Genetics and Cancer, University of Edinburgh, Edinburgh, UK
65, Department of Psychiatry and Mental Health, University of Cape Town, Cape Town, SA
66, Department of Psychiatry, Lausanne University Hospital and University of Lausanne, Prilly, VD, CH
67, Instituto de Investigación Biosanitaria ibs.GRANADA, Granada, ES
68, Department of Psychiatry, Faculty of Medicine and Institute of Neurosciences, Biomedical Research Centre (CIBM), University of Granada, Granada, ES
69, Université de Paris Cité, INSERM U1266, Institute of Psychiatry and Neuroscience of Paris, GHU Paris Psychiatry and Neuroscience, Paris, FR
70, Translational Biology, Biogen, Cambridge, MA, US
71, Nuffield Department of Population Health, University of Oxford, Oxford, UK
72, MRC Population Health Research Unit, University of Oxford, Oxford, UK
73, Institute of Neuroscience and Medicine (INM-1), Research Center Juelich, Juelich, DE
74, Human Genomics Research Group, Department of Biomedicine, University of Basel, Basel, CH
75, Institute of Medical Genetics and Pathology, University Hospital Basel, University of Basel, Basel, CH

76, Institute of Human Genetics, University of Bonn, School of Medicine & University Hospital Bonn, Bonn, DE
77, Nic Waals Institute, Lovisenberg Diakonale Hospital, Oslo, NO
78, Centre for Advanced Imaging, University of Queensland, Saint Lucia, QLD, AU
79, Psychological Medicine, Cardiff University, Cardiff, WLS, UK
80, Centre for Neuropsychiatric Genetics and Genomics, Cardiff University, Cardiff, WLS, UK
81, The Lothian Birth Cohorts, University of Edinburgh, Edinburgh, UK
82, Department of Biological Psychology & Amsterdam Public Health Research Institute, Vrije Universiteit Amsterdam, Amsterdam, NL
83, Department of Child and Adolescent Psychiatry, Psychosomatics and Psychotherapy, University Hospital Essen, Unversity of Duisburg-Essen, Duisburg, DE
84, MRC Centre for Environment and Health, School of Public Health, Imperial College London, London, UK
85, Imperial College Dementia Research Institute, Imperial College London, London, UK
86, Department of Psychiatry and Behavioral Sciences, Johns Hopkins University School of Medicine, Baltimore, MD, US
87, Department of Psychiatry and Psychotherapy, Research Group Stress Related Disorders, Central Institute of Mental Health, Medical Faculty Mannheim, Heidelberg University, Mannheim, BW, DE
88, Department of Clinical Immunology, Copenhagen University Hospital, Rigshospitalet, Copenhagen, CPH, DK
89, Department of Clinical Immunology, Aarhus University Hospital, Aarhus, DK
90, Department of Psychiatry, Istanbul University, Istanbul, TR
91, Department of Medical Genetics, Oslo University Hospital, Oslo, OSL, NO
92, NORMENT, Department of Clinical Science, University of Bergen, Bergen, NO
93, Virginia Institute for Psychiatric & Behavioral Genetics, Virginia Commonwealth University, Richmond, VA, US
94, Psychiatry Department / Huntsman Mental Health Institute, University of Utah School of Medicine, Salt Lake City, UT, US
95, Center for Genomic Research, University of Utah School of Medicine, Salt Lake City, UT, US
96, Department of Psychiatry and Psychotherapy, Medical Center, University of Freiburg, Faculty of Medicine, University of Freiburg, Freiburg, DE
97, Division of Mental Health Care, St. Olavs Hospital, Trondheim University Hospital, Trondheim, TRD, NO
98, Department of Psychiatry, Sørlandet Hospital, Kristiansand, AG, NO
99, University of Oslo, NORMENT Centre, Institute of Clinical Medicine, Oslo, OSL, NO
100, Center for Genomic Medicine, Massachusetts General Hospital, Boston, MA, US
101, Centre for Addiction and Mental Health, Toronto, ON, CA
102, Department of Psychiatry and Psychotherapy, University Medical Center of the Johannes Gutenberg University Mainz, Mainz, DE
103, Mental Health Center Copenhagen, Mental Health Services Capital Region of Denmark, Copenhagen, DK
104, Faculty of Health Science, Department of Clinical Medicine, University of Copenhagen, Copenhagen, DK
105, Department of Biomedical and Neuromotor Sciences, University of Bologna, Bologna, IT
106, Michigan Neuroscience Institute, University of Michigan, Ann Arbor, MI, US
107, Wolfson Institute of Population Health, Queen Mary University of London, London, UK
108, School of Computing and Mathematical Sciences, University of Leicester, Leicester, UK
109, Department of Epidemiology and Biostatistics, Imperial College London, London, UK
110, Discipline of Psychiatry, School of Medicine, Trinity College Dublin, Dublin, IE
111, Bioinformatics Research Centre, Aarhus University, Aarhus, DK
112, School of Engineering, University of Iceland, Reykjavik, IS

113, Danish Headache Centre, Department of Neurology, Rigshospitalet, Glostrup, DK
114, Faculty of Medicine, Department of Psychiatry, University of Iceland, Reykjavik, IS
115, Department of Psychiatry, University of Groningen, University Medical Center Groningen, Groningen, NL
116, MRC Human Genetics Unit, Institute for Genetics and Cancer, University of Edinburgh, Edinburgh, UK
117, Brain and Mind Centre, University of Sydney, Sydney, NSW, AU
118, Department of Epidemiology Research, Statens Serum Institut, Copenhagen, DK
119, Interfaculty Institute for Genetics and Functional Genomics, Department of Functional Genomics, University Medicine Greifswald, Greifswald, MV, DE
120, Roche Pharmaceutical Research and Early Development, Pharmaceutical Sciences, Roche Innovation Center Basel, F. Hoffmann-La Roche Ltd, Basel, CH
121, SleepWell Research Program and Department of Psychology and Logopedics, University of Helsinki, Helsinki, FI
122, Blizard Institute, Barts and the London School of Medicine and Dentistry, Queen Mary University of London, London, UK
123, Max Planck Institute of Psychiatry, Munich, BY, DE
124, Department of Psychiatry, University of Helsinki, Helsinki, FI
125, 23andMe Research Team, 23andMe, Inc., Sunnyvale, CA, US
126, Department of Psychological Medicine, University of Worcester, Worcester, UK
127, Institution of Neuroscience and Physiology, University of Gothenburg, Gothenburg, SE
128, Department of Psychiatry and Psychotherapy, Medical University of Vienna, Vienna, AT
129, Department of Psychiatry, Virginia Commonwealth University, Richmond, VA, US
130, Health Care Policy, Harvard Medical School, Boston, MA, US
131, Department of Psychiatry, University of Toronto, Toronto, ON, CA
132, Department of Pharmacology & Toxicology, University of Toronto, Toronto, ON, CA
133, Department of Genetics, Rutgers University, Piscataway, NJ, US
134, Department of Psychiatry, Perelman School of Medicine, University of Pennsylvania, Philadelphia, PA, US
135, Mental Illness Research, Education and Clinical Center, Crescenz VA Medical Center, Philadelphia, PA, US
136, Estonian Genome Centre, Institute of Genomics, University of Tartu, Tartu, EE
137, Department of Women's and Children's Health, Uppsala University, Uppsala, SE
138, Department of Epidemiology and Health Systems, Center for Primary Care and Public Health, Lausanne, VD, CH
139, Swiss Institute of Bioinformatics, Lausanne, VD, CH
140, Department of Computational Biology, University of Lausanne, Lausanne, VD, CH
141, Institute for Molecular Medicine Finland - FIMM, University of Helsinki, Helsinki, FI
142, Department of Psychiatry, Washington University School of Medicine in St. Louis, St. Louis, MO, US
143, Psychiatry, Veterans Affairs Connecticut Healthcare System, West Haven, CT, US
144, Department of Psychiatry, Yale University, New Haven, CT, US
145, Department of Epidemiology and Biostatistics, School of Public Health, Peking University, Beijing, CN
146, Peking University Center for Public Health and Epidemic Preparedness & Response, Peking University, Beijing, CN
147, Mental Health, NHS 24, Glasgow, UK
148, Royal Edinburgh Hospital, NHS Lothian, Edinburgh, UK
149, Department of Human and Molecular Genetics, Virginia Commonwealth University, Richmond, VA, USA

150, Virginia Institute for Psychiatric and Behavioral Genetics, Virginia Commonwealth University, Richmond, VA, USA
151, Department of Psychiatry and Psychotherapy, University of Bonn, Bonn, DE
152, Center for Translational and Computational Neuroimmunology, Columbia University Medical Center, New York, NY, US
153, Institute for Translational Neuroscience, University of Münster, Münster, NRW, DE
154, Novo Nordisk Foundation Center for Basic Metabolic Research, Faculty of Health Science, Copenhagen University, Copenhagen, DK
155, Department of Nursing, Faculty of Health Sciences and Institute of Neurosciences, Biomedical Research Centre (CIBM), University of Granada, Granada, ES
156, School of Public Health, University of Queensland, Brisbane, QLD, AU
157, DZHK (German Centre for Cardiovascular Research), Partner Site Greifswald, Greifswald, MV, DE
158, Institute of Clinical Chemistry and Laboratory Medicine, University Medicine Greifswald, Greifswald, MV, DE
159, Department of Psychiatry, University of Marburg, Marburg, DE
160, Department of Clinical Immunology, Aalborg University Hospital, Aalborg, DK
161, Department of Epidemiology, University of Groningen, University Medical Center Groningen, Groningen, NL
162, Department of Health, Science and Technology, Aalborg University, Aalborg, DK
163, Centre for Integrative Sequencing, iSEQ, Aarhus University, Aarhus, DK
164, Department of Biomedicine-Human Genetics, Aarhus University, Aarhus, DK
165, Population Health, QIMR Berghofer Medical Research Institute, Brisbane, QLD, AU
166, The Fraser Institute, Faculty of Medicine, University of Queensland, Brisbane, QLD, AU
167, Humus, Reykjavik, IS
168, Department of Clinical Medicine, University of Copenhagen, Copenhagen, CPH, DK
169, Department of Population Health Sciences, University of Leicester, Leicester, UK
170, Department of Psychiatry, Universidade Federal de Sao Paulo, Sao Paulo, SP, BR
171, Department of Psychiatry, Rutgers University, Piscataway, NJ, US
172, Department of Clinical Immunology, Zealand University Hospital, Køge, DK
173, Department of Psychiatry and Behavioral Sciences, SUNY Downstate Health Sciences University, Brooklyn, NY, US
174, NMD Pharma, Lexington, MA, US
175, Psychiatry, University of Pittsburgh Medical Centre, Pittsburgh, PA, US
176, Psychiatry, Universidade Federal do Rio Grande do Sul, Porto Alegre, BR
177, Department of Psychiatry, University Medical Center Groningen, Groningen, NL
178, Research School of Behavioural and Cognitive Neurosciences (BCN), University of Groningen, Groningen, NL
179, Institute of Biological Psychiatry, Mental Health Center Sct. Hans, Mental Health Services Capital Region of Denmark, Copenhagen, DK
180, Neurogenomics Division, The Translational Genomics Research Institute (TGEN), Phoenix, AZ, US
181, Human Genetics Branch, NIMH Division of Intramural Research Programs, Bethesda, MD, US
182, Department of Psychiatry and Psychotherapy, University Medical Center Göttingen, Goettingen, NI, DE
183, Division of Cancer Epidemiology and Genetics, National Cancer Institute, Bethesda, MD, US
184, School of Medicine and Dentistry, James Cook University, Townsville, QLD, AU
185, Division of Mental Health and Addiction, Oslo University Hospital, Oslo, OSL, NO
186, NORMENT, Institute of Clinical Medicine, University of Oslo, Oslo, OSL, NO
187, Institute of Health and Wellbeing, University of Glasgow, Glasgow, UK
188, Division of Population Health and Genomics, Ninewells Hospital and School of Medicine, University of Dundee, Dundee, UK
189, Faculty of Medicine, University of Iceland, Reykjavik, IS

190, SAMRC Unit on Risk & Resilience in Mental Disorders, Department of Psychiatry and Mental Health, University of Cape Town, Cape Town, SA
191, Department of Psychiatry and Psychotherapy, University of Marburg, Marburg, HE, DE
192, Department of Psychiatry, Psychotherapy and Psychosomatics, Dr. Fontheim Mentale Gesundheit, Liebenburg, DE
193, Institute of Epidemiology and Social Medicine, University of Münster, Münster, NRW, DE
194, Institute for Community Medicine, University Medicine Greifswald, Greifswald, MV, DE
195, Wolfson Centre for Young People's Mental Health, Division of Psychological Medicine and Clinical Neurosciences, Cardiff University, Cardiff, WLS, UK
196, Department of Biological Sciences, Purdue University, West Lafayette, IN, US
197, Imperial College BHF Centre for Research Excellence, Imperial College London, London, UK
198, Genomics Program, University of South Florida College of Public Health, Tampa, FL, US
199, Department of Internal Medicine, Erasmus University Medical Center Rotterdam, Rotterdam, NL
200, Management Section, Statens Serum Institut, Copenhagen, DK
201, Department of Clinical Medicine, University of Copenhagen, Copenhagen, DK
202, Xperimed LLC, Basel, CH
203, Psychiatry, USUHS, Bethesda, US
204, Department of Psychiatry and Psychotherapy, University Medicine Greifswald, Greifswald, MV, DE
205, Department of Psychiatry, Leiden University Medical Center, Leiden, NL
206, Department of Epidemiology and Population Health, Albert Einstein College of Medicine, Bronx, NY, US
207, Department of Psychiatry, Columbia University College of Physicians and Surgeons, New York, NY, US
208, Division of Epidemiology, New York State Psychiatric Institute, New York, NY, US
209, Department of Neurology, Oslo University Hospital, Oslo, NO
210, HUNT All-In Psychiatry
211, China Kadoorie Biobank Collaborative Group
212, Genes & Health Research Team
213, KG Jebsen Centre for Neurodevelopmental Research, University of Oslo, Oslo, OSL, NO
214, Department of Psychiatry, University of Melbourne, Melbourne, VIC, AU
215, Florey Institute of Neuroscience and Mental Health, University of Melbourne, Melbourne, VIC, AU
216, Department of Psychiatry, University of Münster, Münster, NRW, DE
217, Computational Health Centre, Helmholtz Zentrum München, Neuherberg, DE
218, School of Medicine, Technical University of Munich, Munich, BY, DE
219, Helmholtz Pioneer Campus, Helmholtz Zentrum München, Neuherberg, DE
220, Department of Psychiatry, University of Vermont, Burlington, VT, US
221, Department of Cellular, Computational and Integrative Biology, Università degli Studi di Trento, Trento, IT
222, Imperial College Biomedical Research Centre, Imperial College London, London, UK
223, Department of Psychiatry, Psychosomatics and Psychotherapy, Julius-Maximilians-Universität Würzburg, Würzburg, DE
224, Department of Genetics, Department of Neuroscience, Yale University School of Medicine, New Haven, CT, US
225, Psychiatry, Kaiser Permanente Northern California, San Francisco, CA, US
226, K. G. Jebsen Center for Genetic Epidemiology, Department of Public Health and Nursing, Faculty of Medicine and Health Sciences, Norwegian University of Science and Technology (NTNU), Trondheim, NO
227, HUNT Research Center, Department of Public Health and Nursing, Faculty of Medicine and Health Sciences, Norwegian University of Science and Technology (NTNU), Trondheim, NO

228, Department of Research, Innovation and Education, St. Olavs Hospital, Trondheim University Hospital, Trondheim, NO
229, NIHR Leicester Biomedical Research Centre, Glenfield Hospital, Leicester, UK
230, Pathophysiology of Psychiatric Diseases, INSERM, Univ Paris Cité, GHU Paris, Paris, FR
231, Department of Psychiatry & Behavioral Sciences, Stanford University, Stanford, CA, US
232, Neuroscience Therapeutic Area, Janssen Research and Development, LLC, Titusville, NJ, US
233, Second Opinion Outpatient Clinic, GGNet Mental Health, Warnsveld, NL
234, Child and Youth Mental Health Service, Children's Health Queensland Hospital and Health Service, Brisbane, QLD, AU
235, Psychosis Research Unit, Aarhus University Hospital-Psychiatry, Aarhus, DK
236, Department of Psychiatry, Psychosomatics and Psychotherapy, University Hospital of Würzburg, Würzburg, DE
237, Munich Cluster for Systems Neurology (SyNergy), Munich, BY, DE
238, University of Liverpool, Liverpool, UK
239, Human Genetics and Computational Biomedicine, Pfizer Global Research and Development, Groton, CT, US
240, Centre for Quantitative Health, Massachusetts General Hospital, Boston, MA, US
241, Child and Adolescent Psychiatry, Amsterdam UMC, Vrije Universiteit Amsterdam, Amsterdam, NL
242, Complex Trait Genetics, Vrije Universiteit Amsterdam, Amsterdam, NL
243, Wolfson Centre for Young People's Mental Health, Division of Psychological Medicine and Clinical Neurosciences, Cardiff University, Cardiff, UK
244, Department of Biochemistry and Molecular Biology II, Faculty of Pharmacy and Institute of Neurosciences, Biomedical Research Centre (CIBM), University of Granada, Granada, ES
245, Department of Clinical Neuroscience, Karolinska Institutet, Stockholm, SE
246, Division of Research, Kaiser Permanente Northern California, Oakland, CA, US
247, Department of Psychiatry, University of Michigan, Ann Arbor, MI, US
248, Stanley Center for Psychiatric Research, Broad Institute of MIT and Harvard, Cambridge, MA, US
249, Psychiatric and Neurodevelopmental Genetics Unit, Massachusetts General Hospital, Boston, MA, US
250, Psychiatry, UCSD School of Medicine, La Jolla, CA, US
251, Public Health, UCSD School of Public Health, La Jolla, CA, US
252, Psychiatry, Veterans Affairs San Diego Healthcare System, San Diego, CA, US
253, Departments of Genetics and Psychiatry, University of North Carolina at Chapel Hill, Chapel Hill, NC, US
254, Child and Adolescent Psychiatry, Erasmus University Medical Center Rotterdam, Rotterdam, NL
255, Social and Behavioral Science, Harvard T.H. Chan School of Public Health, Boston, MA, US
256, Psychiatry, Dalhousie University, Halifax, NS, CA
257, Institute of Biological Psychiatry, Mental Health Center Sct. Hans, Copenhagen University Hospital, Mental Health Services, Copenhagen, DK
258, GLOBE Institute, Lundbeck Foundation Centre for Geogenetics, University of Copenhagen, Copenhagen, DK
259, Queensland Brain Institute, University of Queensland, Brisbane, QLD, AU
260, Department of Medical & Molecular Genetics, King's College London, London, UK
261, Institute for Genomics and Cancer, University of Edinburgh, Edinburgh, UK

### Supplementary Table captions

**Table S1. Overview of PGC training data.** The GWAS training data consisted of case-control GWAS summary statistics for SCZ (*N*=53,386 cases and 77,258 controls), BIP (*N*=41,917 cases and 371,549 controls), and MDD (*N*=173,140 cases and 331,433 controls. Panel A: We provide the sample sizes (Neff) of the PGC training data and the number of SNPs included to compute the case-control PRS for main QC (INFO>0.9 and MAF>0.10) and for looser QC (INFO>0.3 and MAF>0.01) of which the results are presented in Figure SX and Table SX. Panel B: We show the case-control cohorts excluded from the training data. Specifically, independency between the training data and test data was attained one case-control cohort at the time. Specifically, for SCZ case-control cohort X, the SCZ case-control PRS were based on the SCZ training data excluding cohort X, the BIP case-control PRS were based on the full BIP training data, and the MDD case-control PRS were based on the full MDD training data (and analogue for the BIP case-control cohorts and MDD case-control cohorts). We tested for sample overlap of controls between the MDD controls (used as diagnostic category 'control' in this study) and BIP and SCZ controls; when such overlap was found, the respective SCZ/BIP cohort was excluded from the SCZ/BIP training data to compute the SCZ/BIP PRS in the respective MDD cases and controls.

**Table S2. Overview of PGC test data.** The test data consisted of 11,460 individuals of European ancestry (2,865 of each of SCZ, BIP, MDD, control), subdivided into four cohorts that were matched with respect to country and genotyping platform (GER (N=3,368), UK1 (N=1,136), UK2 (N=6,080), USA (N=876). This Table shows the original case-control cohorts from the PGC working groups forming these matches (sample sizes between brackets). Panel A shows the data as used in the main analyses described in this paper; Panel B shows the data based on additional subject QC (see Methods); Panel C shows the full data available after the match with respect to country and genotyping platform. Note that the data in Panel A and Panel B are a random subset of Panel C such data are restricted to 25% of each of SCZ, BIP, MDD and control. Note that mdd0 yields the diagnostic category 'controls' in this paper (bip0 and scz0 are not used as controls as these may contain MDD cases).

**Table S3. Numerical values of Figure 1: Calibration and accuracy of DDx-PRS and Marginal-PRS in simulations.** A detailed description is provided in the caption of Figure 1.

**Table S4. Accuracy of case vs. control (resp. rest) comparisons of DDx-PRS, Marginal-PRS and standard case-control PRS in simulations.** We report the accuracy (AUC) of case vs. rest and case vs. control, for schizophrenia (SCZ), bipolar disorder (BIP) and major depressive disorder (MDD) for three methods: DDx-PRS, Marginal-PRS and standard case-control PRS (i.e., as outputted by PRS-CS). These secondary analyses show that the power to distinguish SCZ vs. control (resp. BIP vs. control and MDD vs. control) was comparable for DDx-PRS, Marginal-PRS and the standard case-control PRS. As expected, case vs. rest accuracy was lower than case vs. control accuracy for SCZ (AUC of 0.678±0.001 for SCZ vs. rest, 0.745±0.002 for SCZ vs. control for DDx-PRS) and BIP (0.629±0.002 for BIP vs. rest, 0.701±0.002 for BIP vs. control)—but slightly higher than case vs. control accuracy for MDD (0.618±0.002 for MDD vs. rest, 0.610±0.002 for MDD vs. control), because MDD cases are genetically more similar to controls than to SCZ cases and BIP cases.

**Table S5. Numerical values of Figure S1: Calibration and accuracy of DDx-PRS and Marginal-PRS for comparisons across all 6 pairs of diagnostic categories in simulations.** A detailed description is provided in the caption of Figure S1.

**Table S6. Numerical values of Figure S2: Calibration slopes of DDx-PRS and Marginal-PRS in simulations.** A detailed description is provided in the caption of Figure S2.

**Table S7. Numerical values of Figure S3: Calibration and accuracy of DDx-PRS and Marginal-PRS at double training sample size in simulations.** A detailed description is provided in the caption of Figure S3.

**Table S8. Numerical values of Figure S4: Calibration and accuracy of DDx-PRS and Marginal-PRS with smaller genetic correlation SCZ-BIP in simulations.** A detailed description is provided in the caption of Figure S4.

**Table S9. Numerical values of Figure S5: Calibration and accuracy of DDx-PRS and Marginal-PRS with no overlap of controls in training data in simulations.** A detailed description is provided in the caption of Figure S5.

**Table S10. Numerical values of Figure 3: Calibration and accuracy of DDx-PRS and Marginal-PRS in PGC data.** A detailed description is provided in the caption of Figure 3.

**Table S11. Accuracy of case vs. control (resp. rest) comparisons of DDx-PRS, Marginal-PRS and standard case-control PRS in PGC data.** We report the accuracy (AUC) of case vs. rest and case vs. control, for schizophrenia (SCZ), bipolar disorder (BIP) and major depressive disorder (MDD) for three methods: DDx-PRS, Marginal-PRS and standard case-control PRS (i.e., as outputted by PRS-CS). These secondary analyses show that the power to distinguish SCZ vs. control (resp. BIP vs. control and MDD vs. control) was comparable for DDx-PRS, Marginal-PRS and the standard case-control PRS. As expected, case vs. rest accuracy was lower than case vs. control accuracy for SCZ (AUC of 0.655±0.010 for SCZ vs. rest, 0.728±0.016 for SCZ vs. control for DDx-PRS), BIP (0.639±0.013 for BIP vs. rest, 0.720±0.013 for BIP vs. control), and MDD (0.591±0.009 for MDD vs. rest, 0.618±0.017 for MDD vs. control.

**Table S12. Numerical values of Figure S8: Calibration and accuracy of DDx-PRS and Marginal-PRS for comparisons across all 6 pairs of diagnostic categories in PGC data.** A detailed description is provided in the caption of Figure S8.

**Table S13. Numerical values of Figure S9: Calibration slopes of DDx-PRS and Marginal-PRS in PGC data.** A detailed description is provided in the caption of Figure S9.

**Table S14. Numerical values of Figure S10: Calibration and accuracy of DDx-PRS and Marginal-PRS for different QC in PGC data.** A detailed description is provided in the caption of Figure S10.

**Table S15. Numerical values of Figure 5: Calibration and accuracy of DDx-PRS, DDx-PRS-tuned and Direct-tuned in PGC data.** A detailed description is provided in the caption of Figure 5.

**Table S16. Numerical values of Figure S11: Calibration and accuracy of DDx-PRS, DDx-PRS-tuned and Direct-tuned in simulations.** A detailed description is provided in the caption of Figure S11.

**Table S17. Numerical values of Figure S12: Calibration and accuracy of DDx-PRS-tuned and Direct-tuned at smaller tuning sample sizes in PGC data.** A detailed description is provided in the caption of Figure S12.

**Table S18. Numerical values of Figure S13: Calibration and accuracy of DDx-PRS-tuned and Direct-tuned using within-cohort tuning data in PGC data.** A detailed description is provided in the caption of Figure S13.

**Table S19. Numerical values of Figure 7: Projections of clinical utility at larger training sample sizes.** A detailed description is provided in the caption of Figure 7.

**Table S20. True vs. predicted diagnosis probabilities in deciles of predicted diagnosis probability of DDx-PRS in PGC data.** This Table accompanies main Figure 7 and reports the true vs. the predicted diagnosis probabilities in deciles of predicted diagnosis probability. This secondary Table shows that the true and predicted diagnosis probabilities are close. Note that the absolute difference between the true and predicted diagnosis probabilities are very close to the ICIs reported in main Figure 4, as expected as the ICI measure the average absolute difference between the predicted and true probabilities, weighted by the density of predicted probabilities (thus reflects the average error of the predicted probabilities).

**Table S21. Numerical values of Figure S14: True diagnosis probability per decile of predicted diagnosis probability of DDx-PRS, DDx-PRS-tuned and Direct-tuned in PGC data.** A detailed description is provided in the caption of Figure S14.

**Table S22. Numerical values of Figure S15: True diagnosis probability per decile of predicted diagnosis probability of DDx-PRS in PGC data and in simulations at the exact empirical values.** A detailed description is provided in the caption of Figure S15.

**Table S23. Variances, covariances and means across disorders of liabilities and case-control PRS (overall and for each configuration of liabilities).** The variances/covariances and means are displayed for the case-control PRS and liabilities for SCZ/BIP/MDD, for the full population and for every configuration of liabilities (see main Table 1). The approximations for the full population are based GWAS results from the PGC case-control training data, S-LDSC, cross-trait LDSC, and case-control PRS computed in 1000G; the results for every configuration of liabilities are based on analytical computations (see the description of step 3 of DDx-PRS in the Method section for details).

**Table S24. Correlation of case-control PRS in 1000G data based on training data with different cohorts held-out.** Of the training data, the test data was left out one cohort at a time (Table S1), resulting in slightly different training data for the test cohorts. Here we show the correlation of the case-control PRS based on these slightly different training data sets in 1000G. The average correlation of the PRS is 0.96 for SCZ, 0.95 for BIP and 0.96 for MDD; the minimum correlation of the PRS is 0.94

for SCZ, 0.93 for BIP and 0.95 for MDD. The high correlation between these case-control PRS validates the approach to exclude one test cohort at the time to obtain independency between training data and test data. (We note the slight heterogeneity thus introduced in the test data could results in slight conservative bias.)

**Table S25. R2 of case-control PRS in SCZ (resp. BIP) case vs. matched SCZ (resp. BIP) controls & SCZ (resp. BIP) case vs. MDD controls.** We report the $r^2$ the case-control PRS in contrasting the SCZ cases (resp. BIP cases) to their matched SCZ controls (resp. BIP controls) based on the original PGC studies, and the $r^2$ of contrasting the SCZ cases (res. BIP cases) to the matched MDD controls (as applied as diagnostic category 'controls' in this study). These $r^2$ are close thereby confirming that a valid match was made for the merged data used in this paper.

### Supplementary Figures

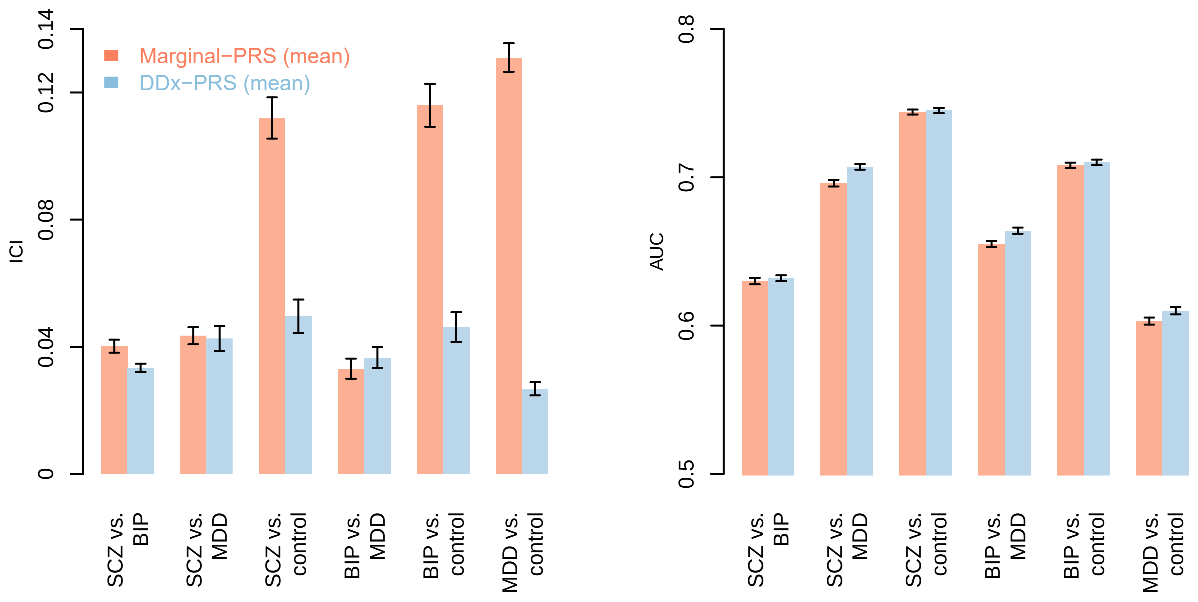

**Figure S1. Calibration and accuracy of DDx-PRS and Marginal-PRS for comparisons across all 6 pairs of diagnostic categories in simulations.** This Figure is analogue to main Figure 1 and reports the calibration (Integrated Calibration Index; ICI) and accuracy (area under the ROC curve; AUC) for the comparisons of schizophrenia (SCZ) vs. bipolar disorder (BIP), SCZ vs. major depressive disorder (MDD), SCZ vs. control, BIP vs. MDD, BIP vs. control and MDD vs. control, for two methods: Marginal-PRS (red) and DDx-PRS (blue). Results are based on 50 simulation replicates; mean values are displayed in light red and light blue. Error bars denote standard errors. Numerical results are reported in Table S5. These secondary analyses show that DDx-PRS attained substantially better calibration and slightly higher accuracy than Marginal-PRS for these 6 comparisons; this is in line with the findings of the four comparisons of one diagnostic category vs. rest (main Figure 1).

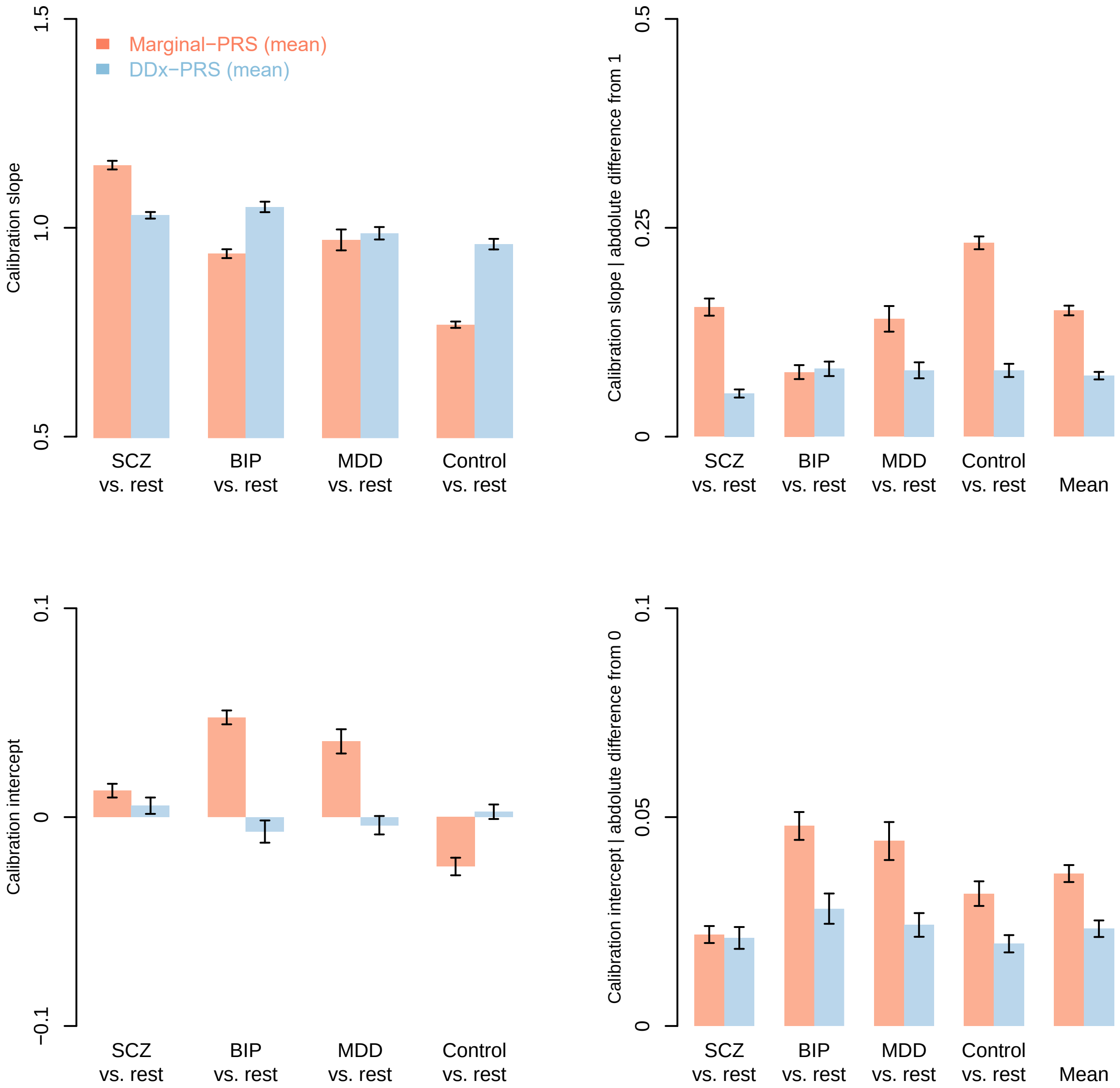

**Figure S2. Calibration slopes of DDx-PRS and Marginal-PRS in simulations.** This Figure accompanies main Figure 1 and reports the calibration slopes and intercepts for the comparisons of schizophrenia (SCZ) vs. rest, bipolar disorder (BIP) vs. rest, major depressive disorder (MDD) vs. rest, and control vs. rest, for two methods: Marginal-PRS (red) and DDx-PRS (blue). Results are based on 50 simulation replicates; mean values are displayed in light red and light blue. Error bars denote standard errors. Perfect calibration would attain a slope of 1 and intercept of 0. Numerical results are reported in Table S6. These secondary analyses show that DDx-PRS attained substantially better calibration than Marginal-PRS with a calibration slope closer to 1 and calibration intercept closer to 0; this is in line with the finding from the ICI (main Figure 1).

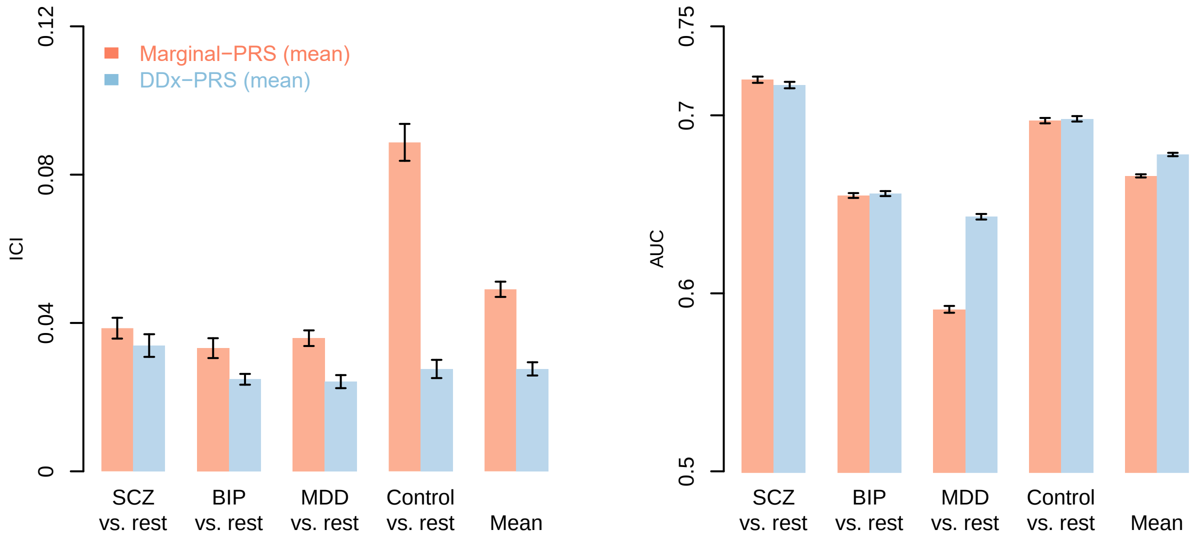

**Figure S3. Calibration and accuracy of DDx-PRS and Marginal-PRS at double training sample size in simulations.** This Figure is analogue to main Figure 1 and reports results at double training sample size in simulations. We report the calibration (Integrated Calibration Index; ICI) and accuracy (area under the ROC curve; AUC) for the comparisons of schizophrenia (SCZ) vs. rest, bipolar disorder (BIP) vs. rest, major depressive disorder (MDD) vs. rest, control vs. rest and the mean across these four comparisons, for two methods: Marginal-PRS (red) and DDx-PRS (blue). Results are based on 50 simulation replicates; mean values are displayed in light red and light blue. Error bars denote standard errors. Numerical results are reported in Table S7. These secondary analyses show that DDx-PRS attained substantially better calibration than Marginal-PRS (with ICI values similar to main Figure 1a) and slightly higher accuracy than Marginal-PRS (with AUC values larger than main Figure 1b for both methods, as expected).

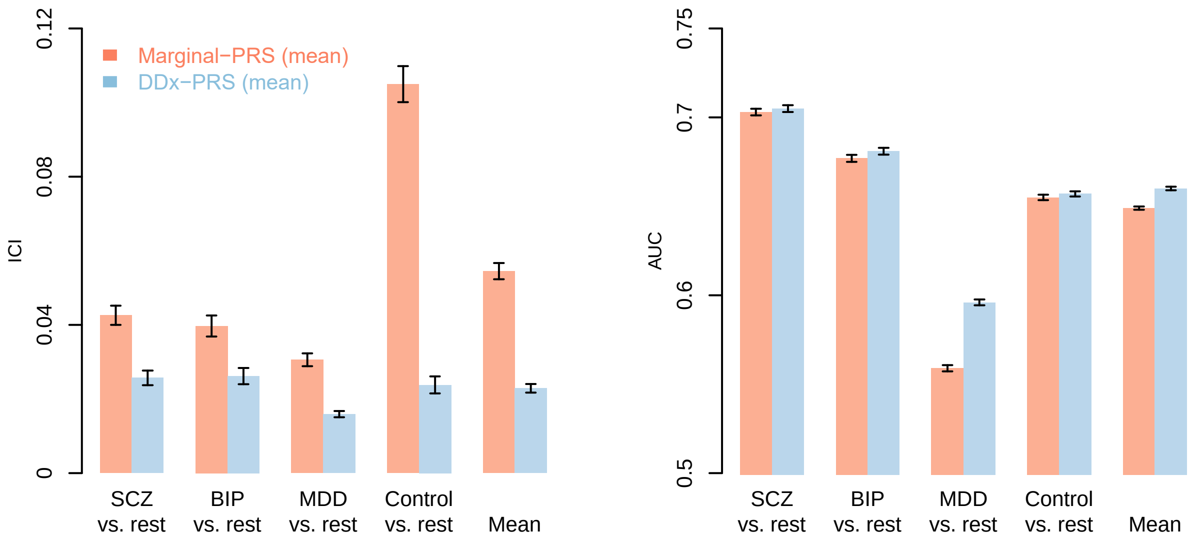

**Figure S4. Calibration and accuracy of DDx-PRS and Marginal-PRS with smaller genetic correlation SCZ-BIP in simulations.** This Figure is analogue to main Figure 1 and reports results at a smaller genetic correlation SCZ-BIP of 0.3 (instead of 0.7 in main Figure 1) in simulations. We report the calibration (Integrated Calibration Index; ICI) and accuracy (area under the ROC curve; AUC) for the comparisons of schizophrenia (SCZ) vs. rest, bipolar disorder (BIP) vs. rest, major depressive disorder (MDD) vs. rest, control vs. rest and the mean across these four comparisons, for two methods: Marginal-PRS (red) and DDx-PRS (blue). Results are based on 50 simulation replicates; mean values are displayed in light red and light blue. Error bars denote standard errors. Numerical results are reported in Table S8. These secondary analyses show that DDx-PRS attained better calibration and slightly higher accuracy than Marginal-PRS, with both methods attaining higher accuracy than in our main simulations due to better discrimination between SCZ and BIP (main Figure 1).

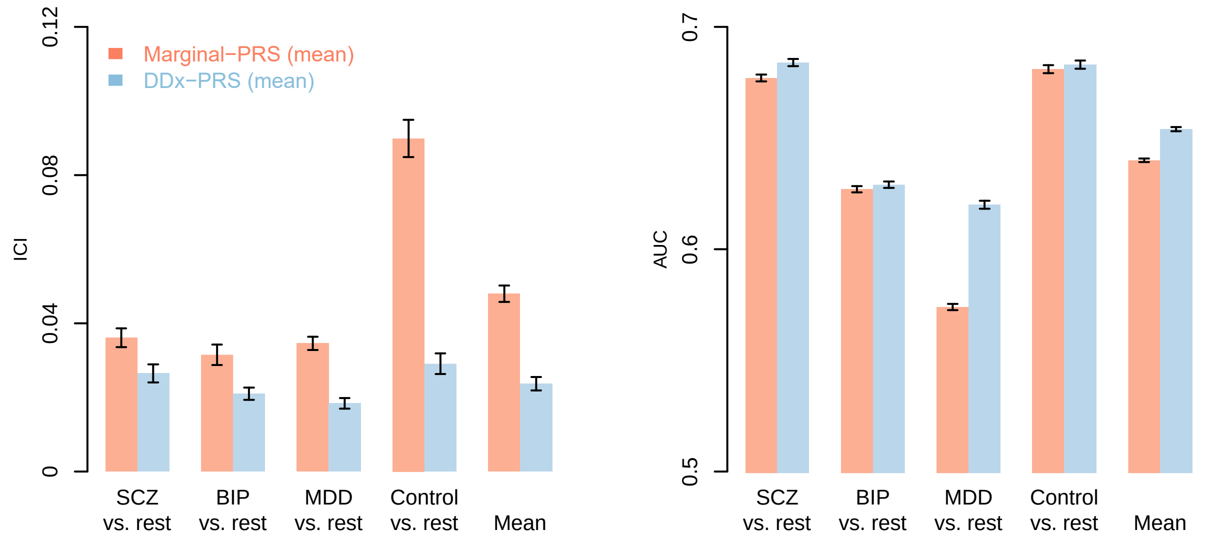

**Figure S5. Calibration and accuracy of DDx-PRS and Marginal-PRS with no overlap of controls in training data in simulations.** This Figure is analogue to main Figure 1 and reports results with no overlap of controls in training data in simulations. We report the calibration (Integrated Calibration Index; ICI) and accuracy (area under the ROC curve; AUC) for the comparisons of schizophrenia (SCZ) vs. rest, bipolar disorder (BIP) vs. rest, major depressive disorder (MDD) vs. rest, control vs. rest and the mean across these four comparisons, for two methods: Marginal-PRS (red) and DDx-PRS (blue). Results are based on 50 simulation replicates; mean values are displayed in light red and light blue. Error bars denote standard errors. Numerical results are reported in Table S9. These secondary analyses show that whether there is overlap among controls in training data (main Figure 1) or no overlap (this Figure) has almost no impact on the results.

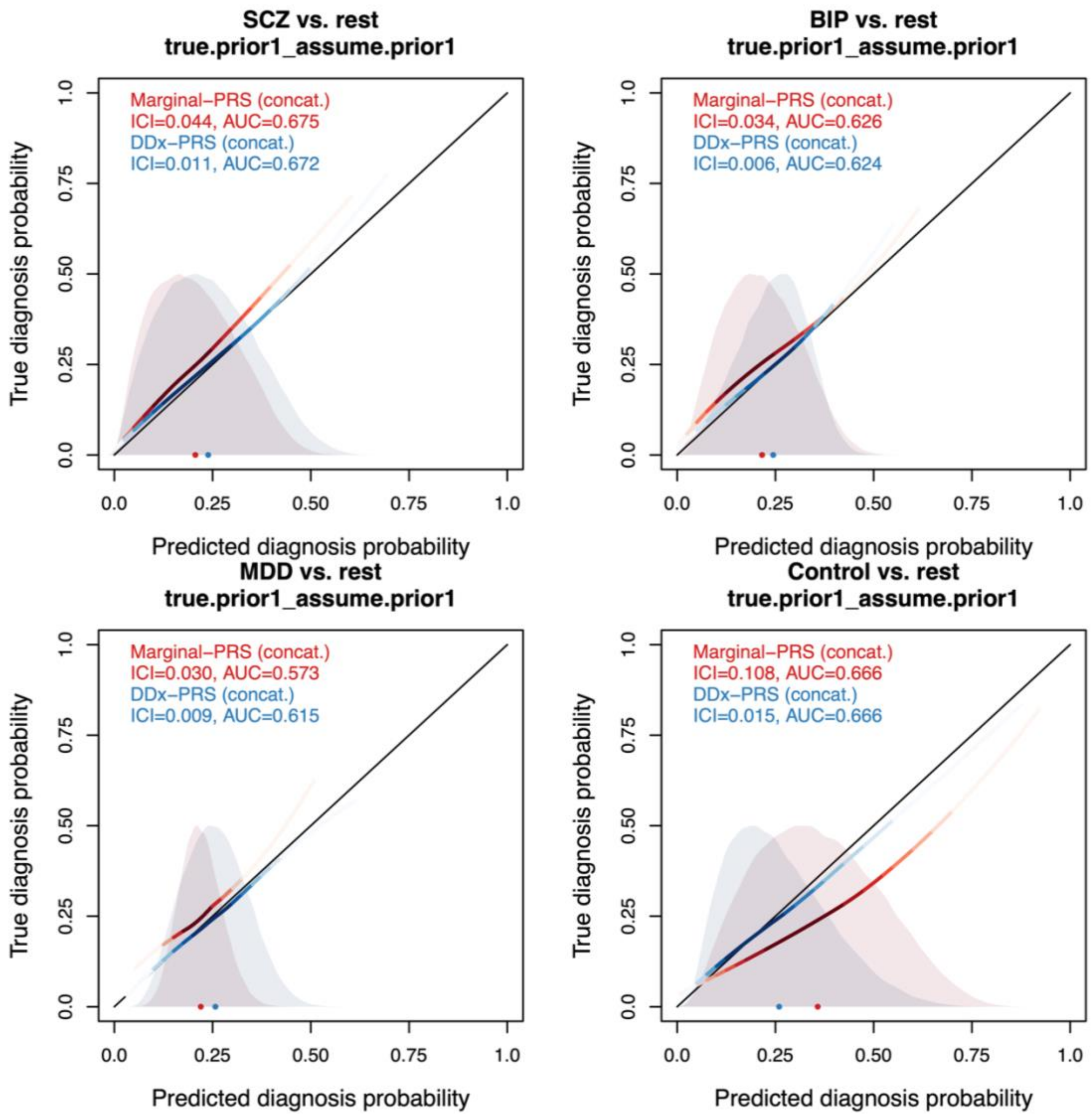

**Figure S6. Calibration curves of DDx-PRS and Marginal-PRS for different prior probabilities in simulations. Panel A.** This Figure is analogue to main Figure 2 and reports results for different assumed and true prior probabilities; this Panel reports true.prior1_assume.prior1 (see below; note that true.prior1_assume.prior1 equals main Figure2). Every Figure panel reports the true diagnosis probability vs. the predicted diagnosis probability for the comparisons of schizophrenia (SCZ) vs. rest, bipolar disorder (BIP) vs. rest, major depressive disorder (MDD) vs. rest, and control vs. rest, for two methods: Marginal-PRS (red) and DDx-PRS (blue). Results are based on concatenated data across 50 simulation replicates. The calibration curves are plotted using a locally estimated scatterplot smoothing (loess)-based smoothing function. The Integrated Calibration Index (ICI) equals the average absolute difference between the calibration curve and the line y=x (plotted in black), weighted by the density of the predicted diagnosis probabilities (displayed in the shaded histograms and in the color-intensity of the calibration curves). AUC, area under the ROC curve. The priors are:

prior1: SCZ=25%, BIP=25%, MDD=25%, control=25%

prior2: SCZ=40%, BIP=20%, MDD=20%, control=20%

prior3: SCZ=20%, BIP=40%, MDD=20%, control=20%

prior4: SCZ=20%, BIP=20%, MDD=40%, control=20%

prior5: SCZ=20%, BIP=20%, MDD=20%, control=40%

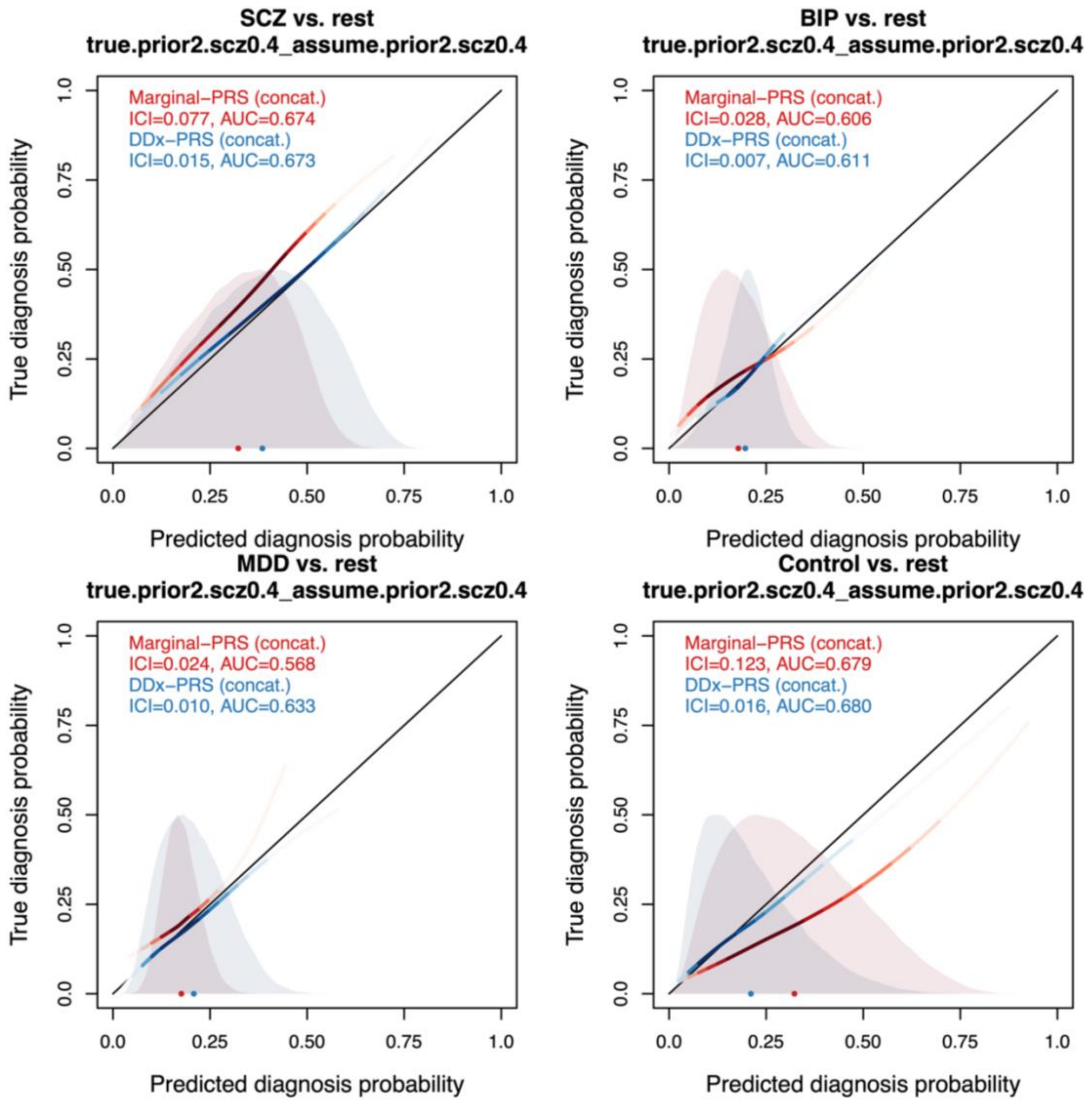

**Figure S6. Calibration curves of DDx-PRS and Marginal-PRS for different prior probabilities in simulations. Panel B.** This Figure is analogue to main Figure 2 and reports results for different assumed and true prior probabilities; this Panel reports true.prior2_assume.prior2 (see below). Every Figure panel reports the true diagnosis probability vs. the predicted diagnosis probability for the comparisons of schizophrenia (SCZ) vs. rest, bipolar disorder (BIP) vs. rest, major depressive disorder (MDD) vs. rest, and control vs. rest, for two methods: Marginal-PRS (red) and DDx-PRS (blue). Results are based on concatenated data across 50 simulation replicates. The calibration curves are plotted using a locally estimated scatterplot smoothing (loess)-based smoothing function. The Integrated Calibration Index (ICI) equals the average absolute difference between the calibration curve and the line y=x (plotted in black), weighted by the density of the predicted diagnosis probabilities (displayed in the shaded histograms and in the color-intensity of the calibration curves). AUC, area under the ROC curve. The priors are:

prior1: SCZ=25%, BIP=25%, MDD=25%, control=25%
prior2: SCZ=40%, BIP=20%, MDD=20%, control=20%
prior3: SCZ=20%, BIP=40%, MDD=20%, control=20%
prior4: SCZ=20%, BIP=20%, MDD=40%, control=20%
prior5: SCZ=20%, BIP=20%, MDD=20%, control=40%

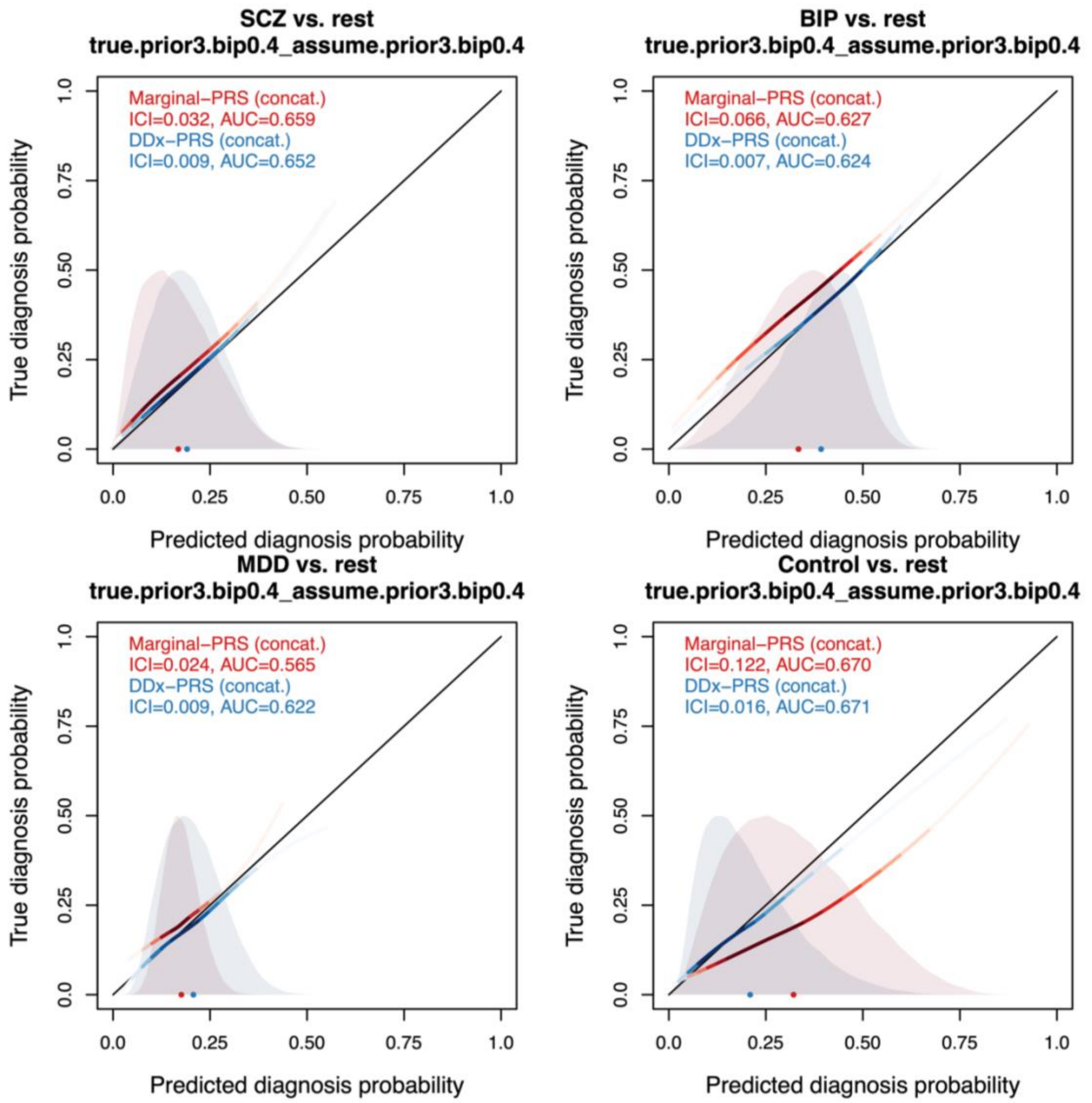

**Figure S6. Calibration curves of DDx-PRS and Marginal-PRS for different prior probabilities in simulations. Panel C.** This Figure is analogue to main Figure 2 and reports results for different assumed and true prior probabilities; this Panel reports true.prior3_assume.prior3 (see below). Every Figure panel reports the true diagnosis probability vs. the predicted diagnosis probability for the comparisons of schizophrenia (SCZ) vs. rest, bipolar disorder (BIP) vs. rest, major depressive disorder (MDD) vs. rest, and control vs. rest, for two methods: Marginal-PRS (red) and DDx-PRS (blue). Results are based on concatenated data across 50 simulation replicates. The calibration curves are plotted using a locally estimated scatterplot smoothing (loess)-based smoothing function. The Integrated Calibration Index (ICI) equals the average absolute difference between the calibration curve and the line y=x (plotted in black), weighted by the density of the predicted diagnosis probabilities (displayed in the shaded histograms and in the color-intensity of the calibration curves). AUC, area under the ROC curve. The priors are:

prior1: SCZ=25%, BIP=25%, MDD=25%, control=25%
prior2: SCZ=40%, BIP=20%, MDD=20%, control=20%
prior3: SCZ=20%, BIP=40%, MDD=20%, control=20%
prior4: SCZ=20%, BIP=20%, MDD=40%, control=20%
prior5: SCZ=20%, BIP=20%, MDD=20%, control=40%

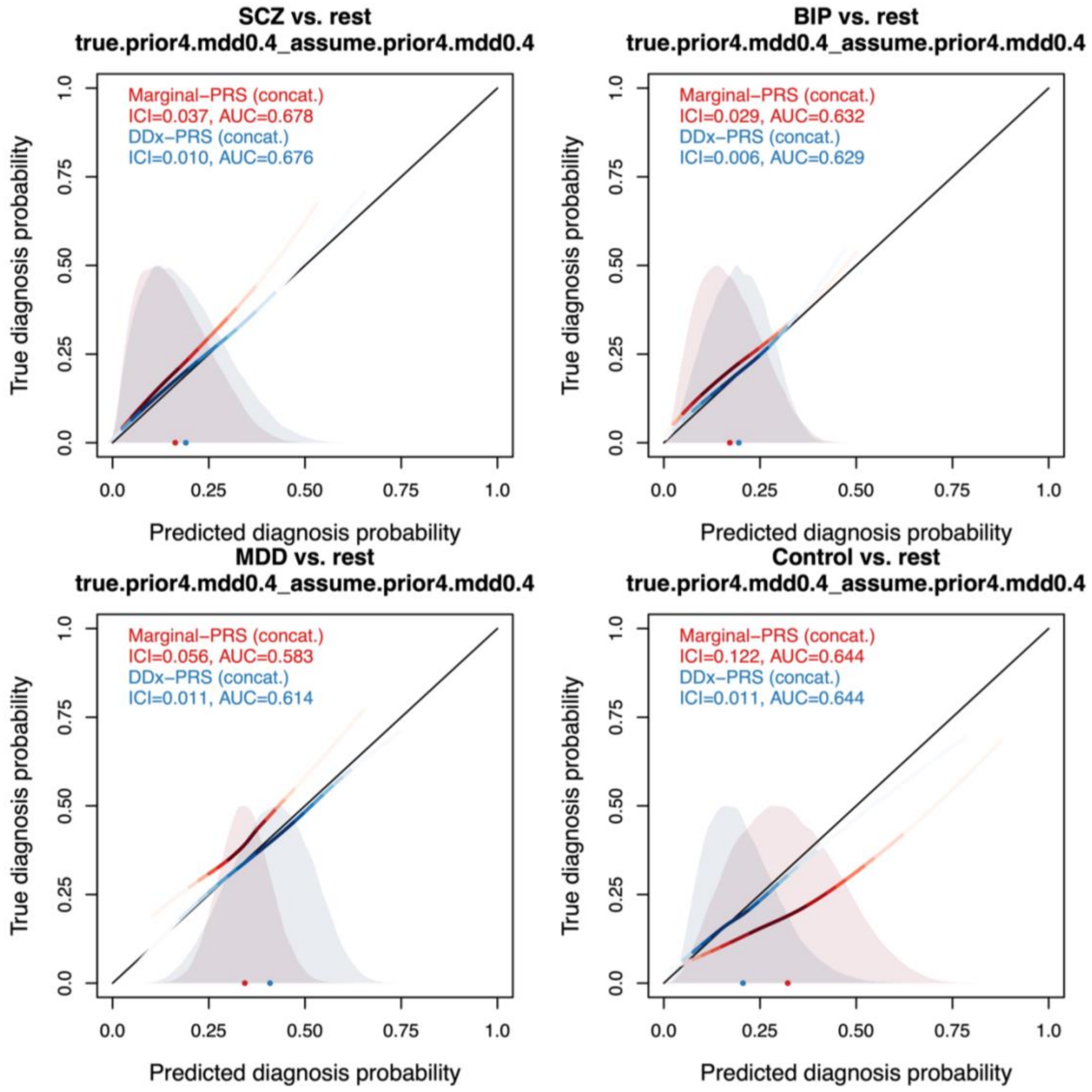

**Figure S6. Calibration curves of DDx-PRS and Marginal-PRS for different prior probabilities in simulations. Panel D.** This Figure is analogue to main Figure 2 and reports results for different assumed and true prior probabilities; this Panel reports true.prior4_assume.prior4 (see below). Every Figure panel reports the true diagnosis probability vs. the predicted diagnosis probability for the comparisons of schizophrenia (SCZ) vs. rest, bipolar disorder (BIP) vs. rest, major depressive disorder (MDD) vs. rest, and control vs. rest, for two methods: Marginal-PRS (red) and DDx-PRS (blue). Results are based on concatenated data across 50 simulation replicates. The calibration curves are plotted using a locally estimated scatterplot smoothing (loess)-based smoothing function. The Integrated Calibration Index (ICI) equals the average absolute difference between the calibration curve and the line y=x (plotted in black), weighted by the density of the predicted diagnosis probabilities (displayed in the shaded histograms and in the color-intensity of the calibration curves). AUC, area under the ROC curve. The priors are:

prior1: SCZ=25%, BIP=25%, MDD=25%, control=25%
prior2: SCZ=40%, BIP=20%, MDD=20%, control=20%
prior3: SCZ=20%, BIP=40%, MDD=20%, control=20%
prior4: SCZ=20%, BIP=20%, MDD=40%, control=20%
prior5: SCZ=20%, BIP=20%, MDD=20%, control=40%

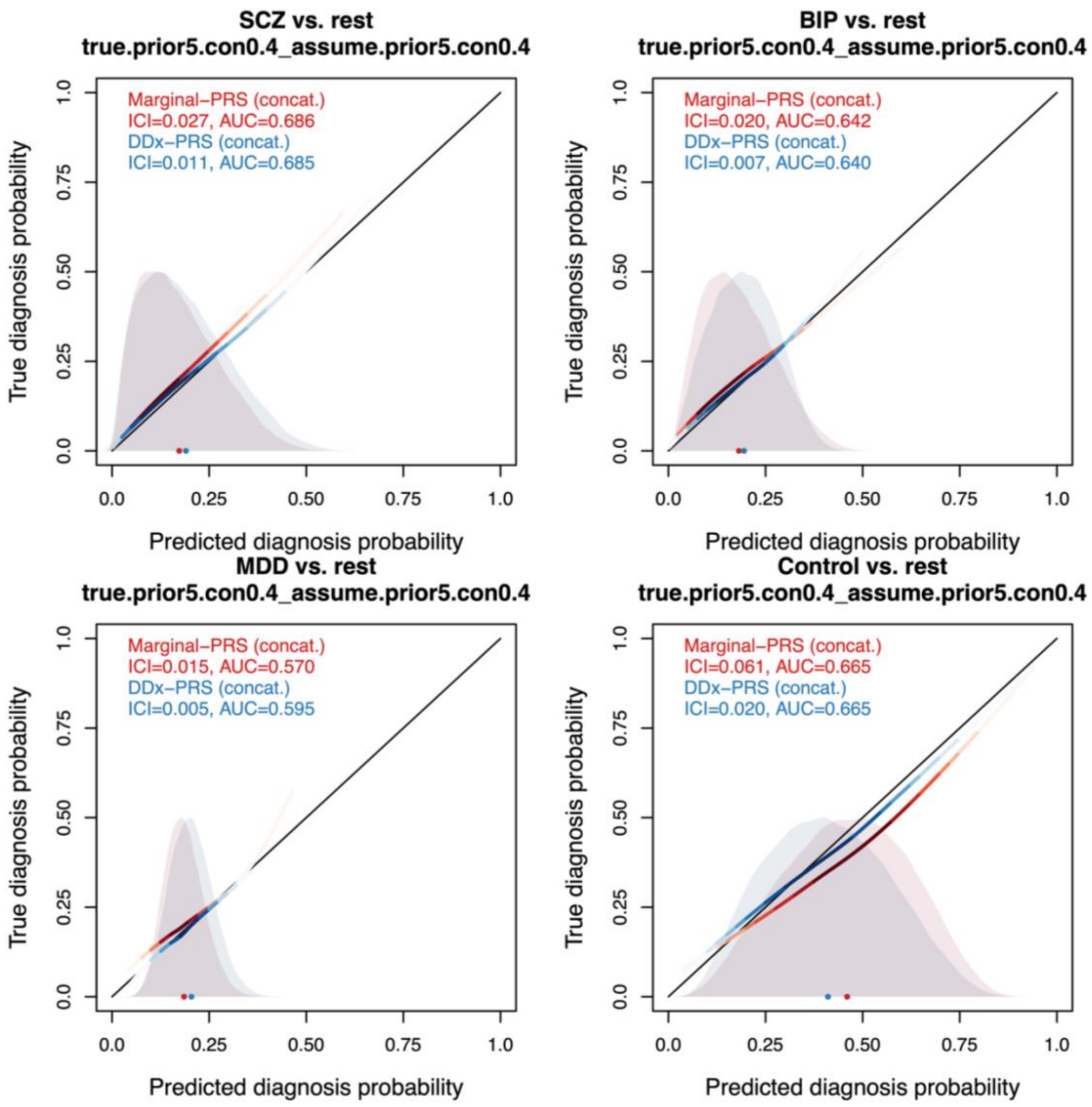

**Figure S6. Calibration curves of DDx-PRS and Marginal-PRS for different prior probabilities in simulations. Panel E.** This Figure is analogue to main Figure 2 and reports results for different assumed and true prior probabilities; this Panel reports true.prior5_assume.prior5 (see below). Every Figure panel reports the true diagnosis probability vs. the predicted diagnosis probability for the comparisons of schizophrenia (SCZ) vs. rest, bipolar disorder (BIP) vs. rest, major depressive disorder (MDD) vs. rest, and control vs. rest, for two methods: Marginal-PRS (red) and DDx-PRS (blue). Results are based on concatenated data across 50 simulation replicates. The calibration curves are plotted using a locally estimated scatterplot smoothing (loess)-based smoothing function. The Integrated Calibration Index (ICI) equals the average absolute difference between the calibration curve and the line y=x (plotted in black), weighted by the density of the predicted diagnosis probabilities (displayed in the shaded histograms and in the color-intensity of the calibration curves). AUC, area under the ROC curve. The priors are:

prior1: SCZ=25%, BIP=25%, MDD=25%, control=25%
prior2: SCZ=40%, BIP=20%, MDD=20%, control=20%
prior3: SCZ=20%, BIP=40%, MDD=20%, control=20%
prior4: SCZ=20%, BIP=20%, MDD=40%, control=20%
prior5: SCZ=20%, BIP=20%, MDD=20%, control=40%

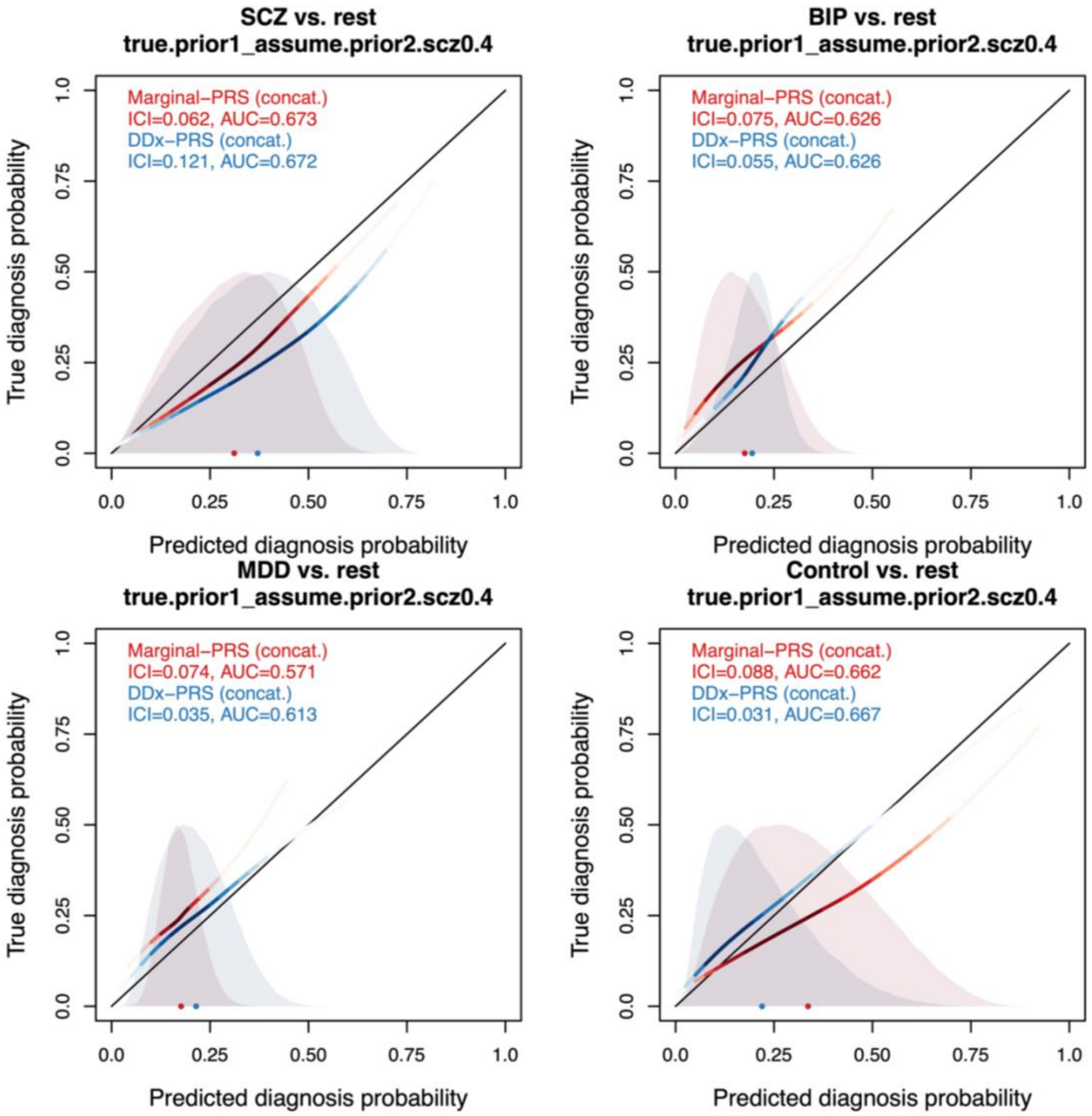

**Figure S6. Calibration curves of DDx-PRS and Marginal-PRS for different prior probabilities in simulations. Panel F.** This Figure is analogue to main Figure 2 and reports results for different assumed and true prior probabilities; this Panel reports true.prior1_assume.prior2 (see below). Every Figure panel reports the true diagnosis probability vs. the predicted diagnosis probability for the comparisons of schizophrenia (SCZ) vs. rest, bipolar disorder (BIP) vs. rest, major depressive disorder (MDD) vs. rest, and control vs. rest, for two methods: Marginal-PRS (red) and DDx-PRS (blue). Results are based on concatenated data across 50 simulation replicates. The calibration curves are plotted using a locally estimated scatterplot smoothing (loess)-based smoothing function. The Integrated Calibration Index (ICI) equals the average absolute difference between the calibration curve and the line y=x (plotted in black), weighted by the density of the predicted diagnosis probabilities (displayed in the shaded histograms and in the color-intensity of the calibration curves). AUC, area under the ROC curve. The priors are:

prior1: SCZ=25%, BIP=25%, MDD=25%, control=25%
prior2: SCZ=40%, BIP=20%, MDD=20%, control=20%
prior3: SCZ=20%, BIP=40%, MDD=20%, control=20%
prior4: SCZ=20%, BIP=20%, MDD=40%, control=20%
prior5: SCZ=20%, BIP=20%, MDD=20%, control=40%

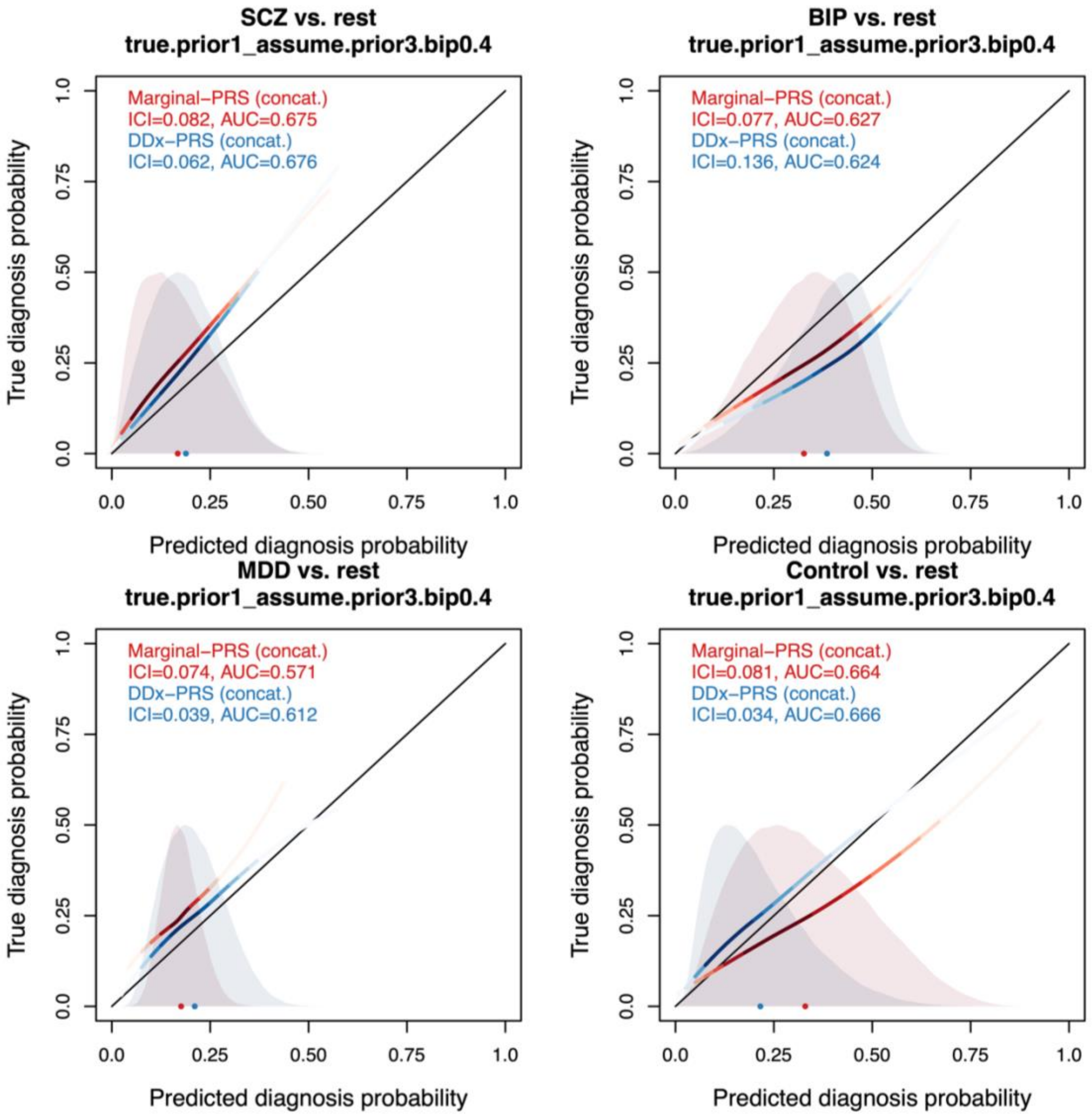

**Figure S6. Calibration curves of DDx-PRS and Marginal-PRS for different prior probabilities in simulations. Panel G.** This Figure is analogue to main Figure 2 and reports results for different assumed and true prior probabilities; this Panel reports true.prior1_assume.prior3 (see below). Every Figure panel reports the true diagnosis probability vs. the predicted diagnosis probability for the comparisons of schizophrenia (SCZ) vs. rest, bipolar disorder (BIP) vs. rest, major depressive disorder (MDD) vs. rest, and control vs. rest, for two methods: Marginal-PRS (red) and DDx-PRS (blue). Results are based on concatenated data across 50 simulation replicates. The calibration curves are plotted using a locally estimated scatterplot smoothing (loess)-based smoothing function. The Integrated Calibration Index (ICI) equals the average absolute difference between the calibration curve and the line y=x (plotted in black), weighted by the density of the predicted diagnosis probabilities (displayed in the shaded histograms and in the color-intensity of the calibration curves). AUC, area under the ROC curve. The priors are:

prior1: SCZ=25%, BIP=25%, MDD=25%, control=25%
prior2: SCZ=40%, BIP=20%, MDD=20%, control=20%
prior3: SCZ=20%, BIP=40%, MDD=20%, control=20%
prior4: SCZ=20%, BIP=20%, MDD=40%, control=20%
prior5: SCZ=20%, BIP=20%, MDD=20%, control=40%

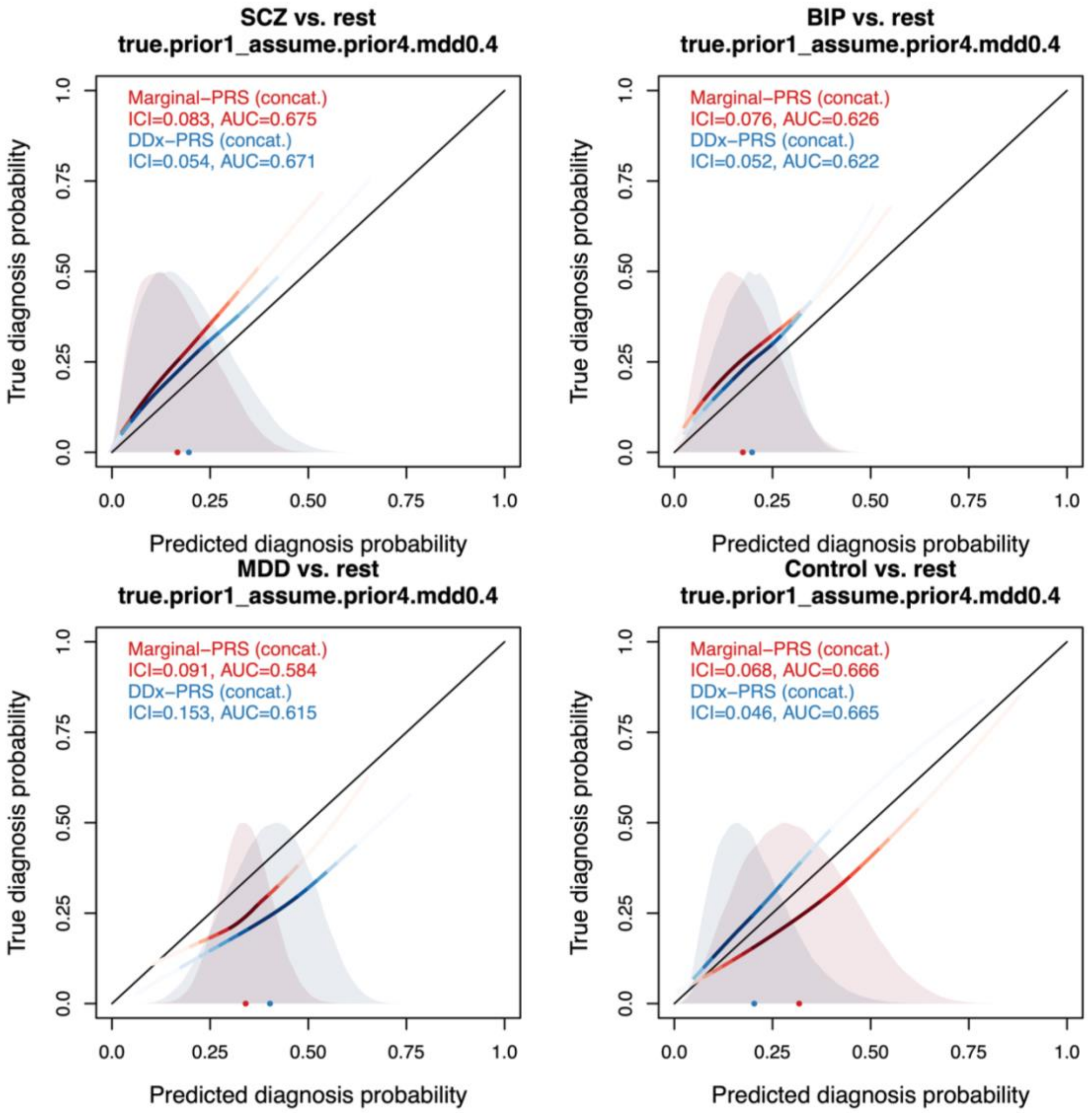

**Figure S6. Calibration curves of DDx-PRS and Marginal-PRS for different prior probabilities in simulations. Panel H.** This Figure is analogue to main Figure 2 and reports results for different assumed and true prior probabilities; this Panel reports true.prior1_assume.prior4 (see below). Every Figure panel reports the true diagnosis probability vs. the predicted diagnosis probability for the comparisons of schizophrenia (SCZ) vs. rest, bipolar disorder (BIP) vs. rest, major depressive disorder (MDD) vs. rest, and control vs. rest, for two methods: Marginal-PRS (red) and DDx-PRS (blue). Results are based on concatenated data across 50 simulation replicates. The calibration curves are plotted using a locally estimated scatterplot smoothing (loess)-based smoothing function. The Integrated Calibration Index (ICI) equals the average absolute difference between the calibration curve and the line y=x (plotted in black), weighted by the density of the predicted diagnosis probabilities (displayed in the shaded histograms and in the color-intensity of the calibration curves). AUC, area under the ROC curve. The priors are:

prior1: SCZ=25%, BIP=25%, MDD=25%, control=25%
prior2: SCZ=40%, BIP=20%, MDD=20%, control=20%
prior3: SCZ=20%, BIP=40%, MDD=20%, control=20%
prior4: SCZ=20%, BIP=20%, MDD=40%, control=20%
prior5: SCZ=20%, BIP=20%, MDD=20%, control=40%

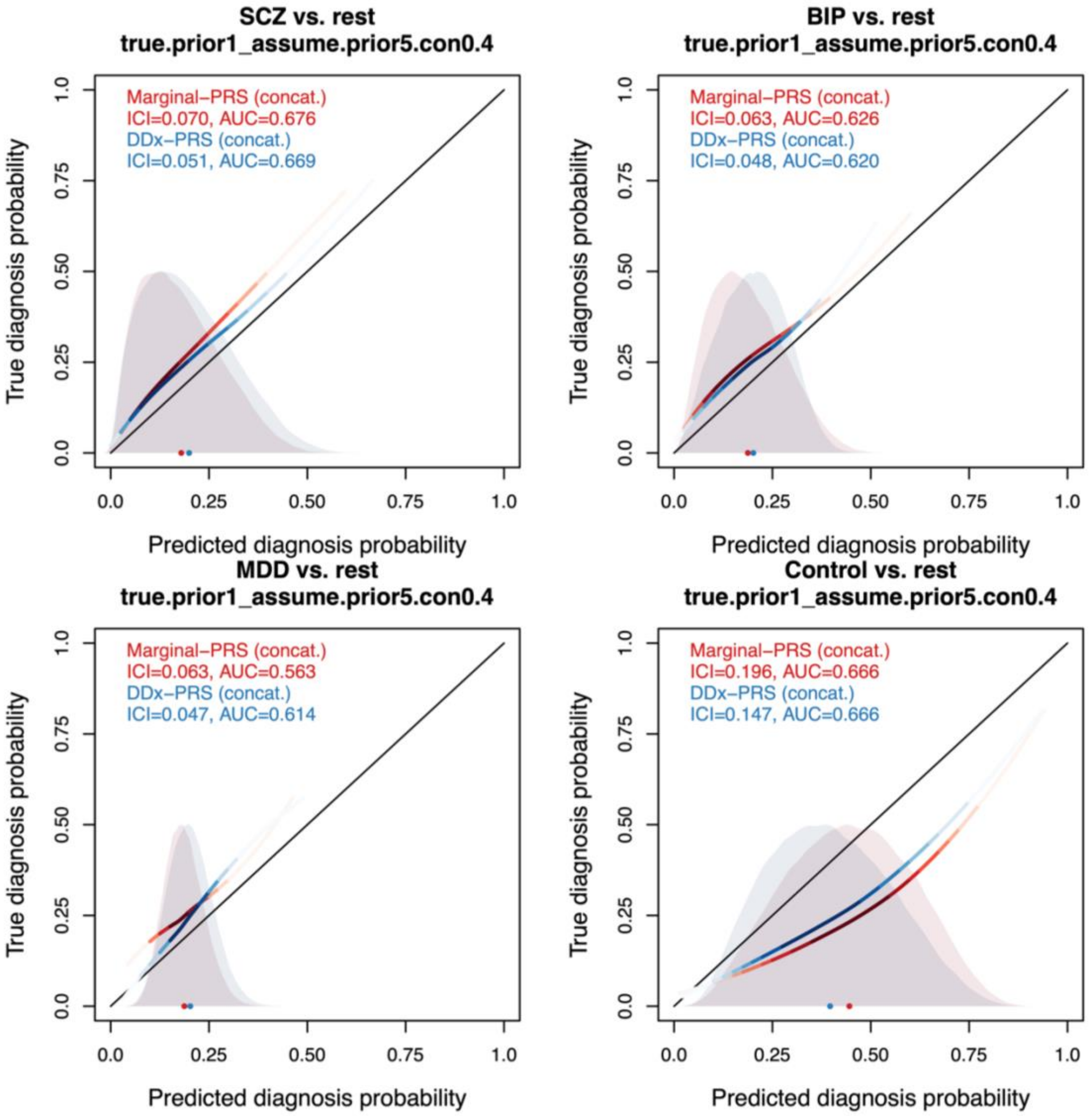

**Figure S6. Calibration curves of DDx-PRS and Marginal-PRS for different prior probabilities in simulations. Panel I.** This Figure is analogue to main Figure 2 and reports results for different assumed and true prior probabilities; this Panel reports true.prior1_assume.prior5 (see below). Every Figure panel reports the true diagnosis probability vs. the predicted diagnosis probability for the comparisons of schizophrenia (SCZ) vs. rest, bipolar disorder (BIP) vs. rest, major depressive disorder (MDD) vs. rest, and control vs. rest, for two methods: Marginal-PRS (red) and DDx-PRS (blue). Results are based on concatenated data across 50 simulation replicates. The calibration curves are plotted using a locally estimated scatterplot smoothing (loess)-based smoothing function. The Integrated Calibration Index (ICI) equals the average absolute difference between the calibration curve and the line y=x (plotted in black), weighted by the density of the predicted diagnosis probabilities (displayed in the shaded histograms and in the color-intensity of the calibration curves). AUC, area under the ROC curve. The priors are:

prior1: SCZ=25%, BIP=25%, MDD=25%, control=25%
prior2: SCZ=40%, BIP=20%, MDD=20%, control=20%
prior3: SCZ=20%, BIP=40%, MDD=20%, control=20%
prior4: SCZ=20%, BIP=20%, MDD=40%, control=20%
prior5: SCZ=20%, BIP=20%, MDD=20%, control=40%

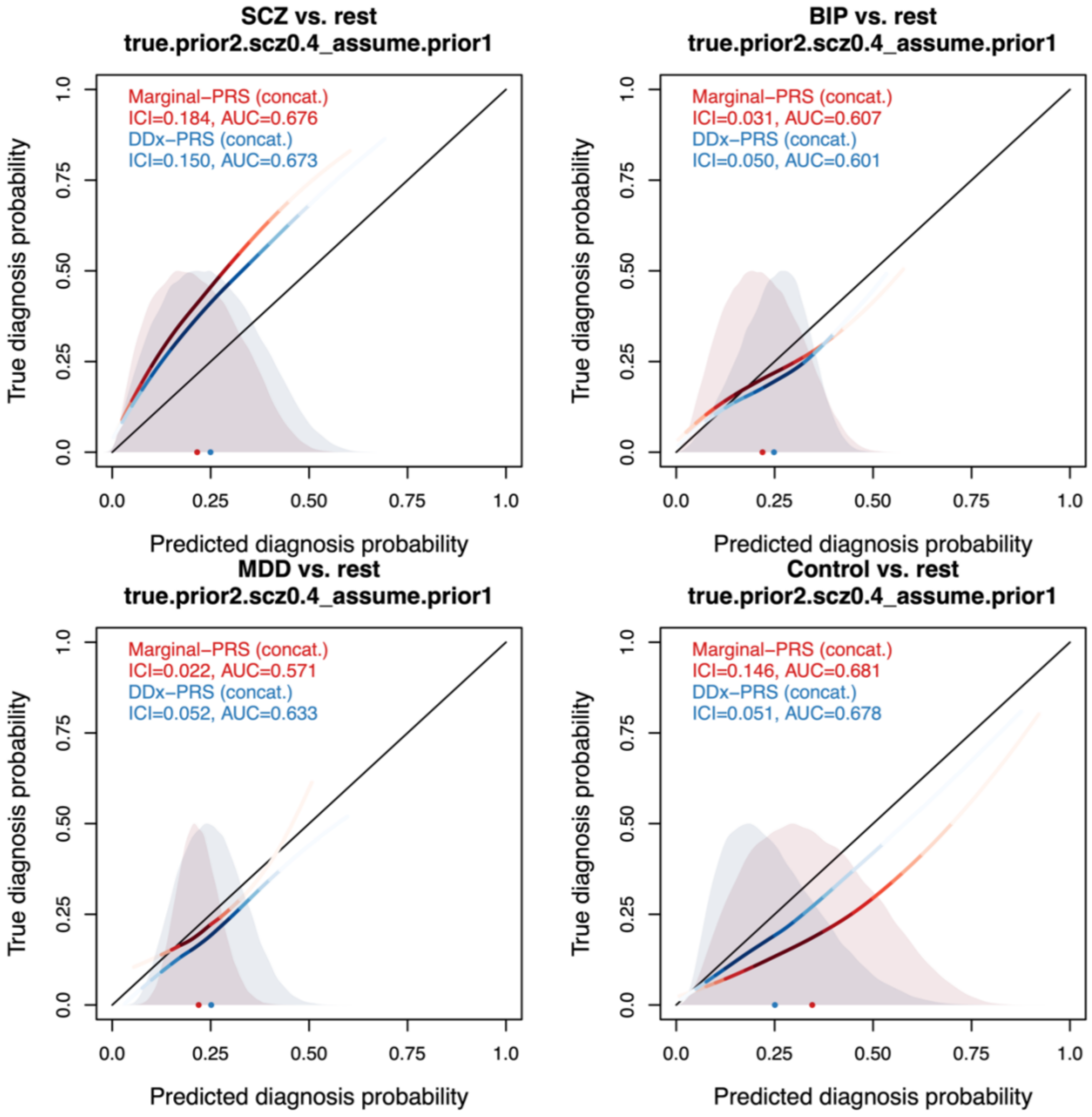

**Figure S6. Calibration curves of DDx-PRS and Marginal-PRS for different prior probabilities in simulations. Panel J.** This Figure is analogue to main Figure 2 and reports results for different assumed and true prior probabilities; this Panel reports true.prior2_assume.prior1 (see below). Every Figure panel reports the true diagnosis probability vs. the predicted diagnosis probability for the comparisons of schizophrenia (SCZ) vs. rest, bipolar disorder (BIP) vs. rest, major depressive disorder (MDD) vs. rest, and control vs. rest, for two methods: Marginal-PRS (red) and DDx-PRS (blue). Results are based on concatenated data across 50 simulation replicates. The calibration curves are plotted using a locally estimated scatterplot smoothing (loess)-based smoothing function. The Integrated Calibration Index (ICI) equals the average absolute difference between the calibration curve and the line y=x (plotted in black), weighted by the density of the predicted diagnosis probabilities (displayed in the shaded histograms and in the color-intensity of the calibration curves). AUC, area under the ROC curve. The priors are:

prior1: SCZ=25%, BIP=25%, MDD=25%, control=25%
prior2: SCZ=40%, BIP=20%, MDD=20%, control=20%
prior3: SCZ=20%, BIP=40%, MDD=20%, control=20%
prior4: SCZ=20%, BIP=20%, MDD=40%, control=20%
prior5: SCZ=20%, BIP=20%, MDD=20%, control=40%

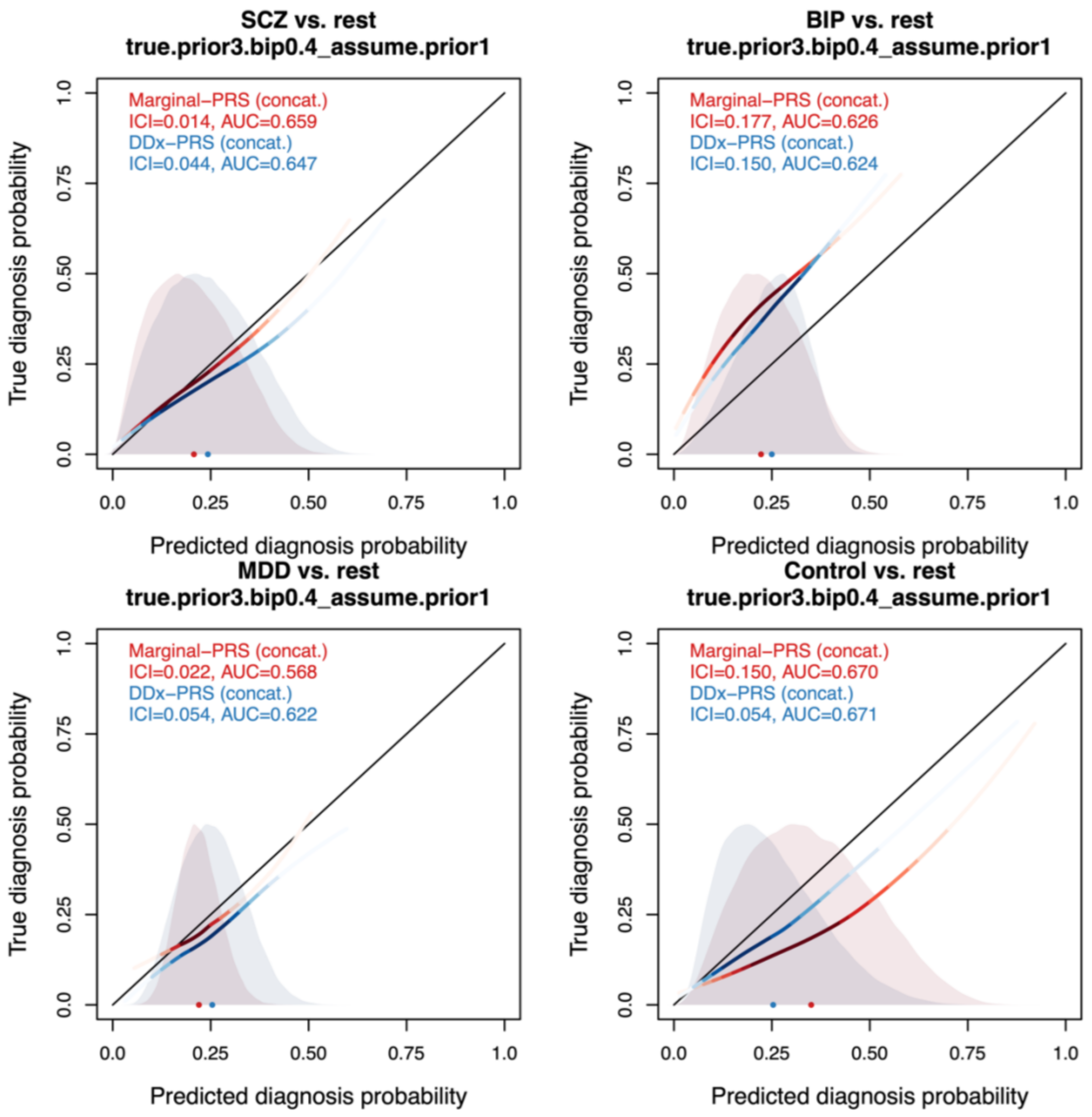

**Figure S6. Calibration curves of DDx-PRS and Marginal-PRS for different prior probabilities in simulations. Panel K.** This Figure is analogue to main Figure 2 and reports results for different assumed and true prior probabilities; this Panel reports true.prior3_asume.prior1 (see below). Every Figure panel reports the true diagnosis probability vs. the predicted diagnosis probability for the comparisons of schizophrenia (SCZ) vs. rest, bipolar disorder (BIP) vs. rest, major depressive disorder (MDD) vs. rest, and control vs. rest, for two methods: Marginal-PRS (red) and DDx-PRS (blue). Results are based on concatenated data across 50 simulation replicates. The calibration curves are plotted using a locally estimated scatterplot smoothing (loess)-based smoothing function. The Integrated Calibration Index (ICI) equals the average absolute difference between the calibration curve and the line y=x (plotted in black), weighted by the density of the predicted diagnosis probabilities (displayed in the shaded histograms and in the color-intensity of the calibration curves). AUC, area under the ROC curve. The priors are:

prior1: SCZ=25%, BIP=25%, MDD=25%, control=25%
prior2: SCZ=40%, BIP=20%, MDD=20%, control=20%
prior3: SCZ=20%, BIP=40%, MDD=20%, control=20%
prior4: SCZ=20%, BIP=20%, MDD=40%, control=20%
prior5: SCZ=20%, BIP=20%, MDD=20%, control=40%

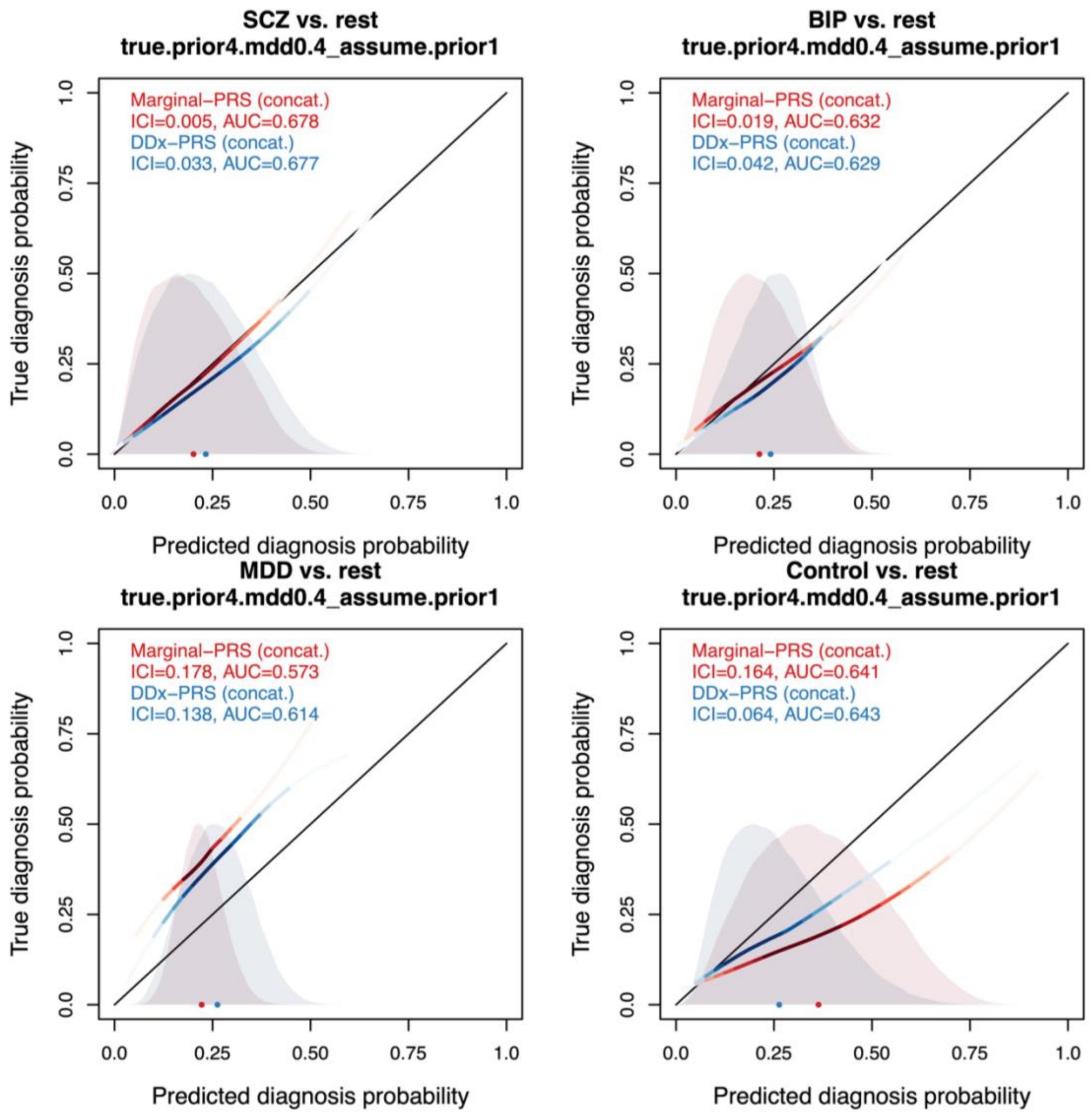

**Figure S6. Calibration curves of DDx-PRS and Marginal-PRS for different prior probabilities in simulations. Panel L.** This Figure is analogue to main Figure 2 and reports results for different assumed and true prior probabilities; this Panel reports true.prior4_assume.prior1 (see below). Every Figure panel reports the true diagnosis probability vs. the predicted diagnosis probability for the comparisons of schizophrenia (SCZ) vs. rest, bipolar disorder (BIP) vs. rest, major depressive disorder (MDD) vs. rest, and control vs. rest, for two methods: Marginal-PRS (red) and DDx-PRS (blue). Results are based on concatenated data across 50 simulation replicates. The calibration curves are plotted using a locally estimated scatterplot smoothing (loess)-based smoothing function. The Integrated Calibration Index (ICI) equals the average absolute difference between the calibration curve and the line y=x (plotted in black), weighted by the density of the predicted diagnosis probabilities (displayed in the shaded histograms and in the color-intensity of the calibration curves). AUC, area under the ROC curve. The priors are:

prior1: SCZ=25%, BIP=25%, MDD=25%, control=25%
prior2: SCZ=40%, BIP=20%, MDD=20%, control=20%
prior3: SCZ=20%, BIP=40%, MDD=20%, control=20%
prior4: SCZ=20%, BIP=20%, MDD=40%, control=20%
prior5: SCZ=20%, BIP=20%, MDD=20%, control=40%

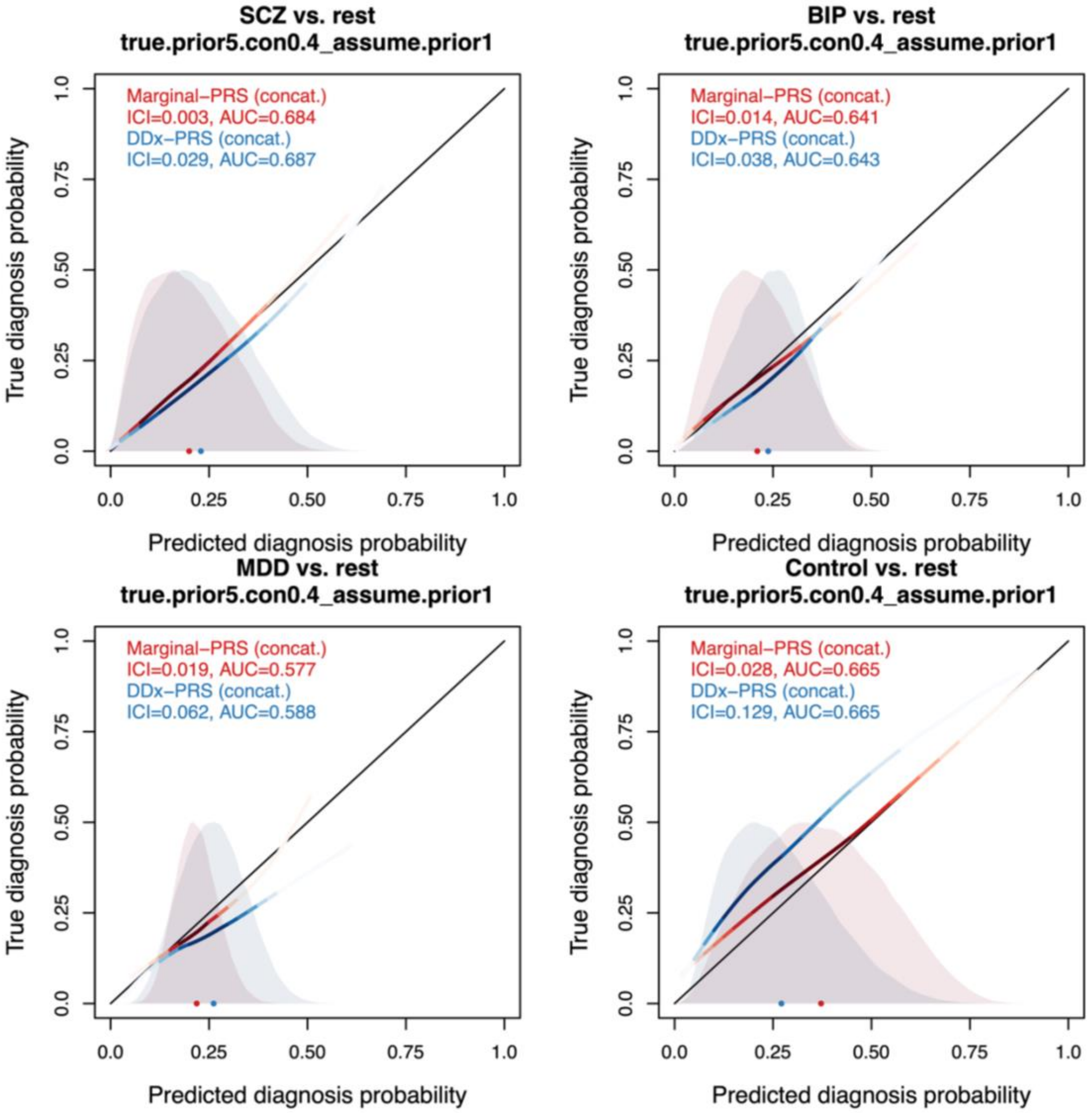

**Figure S6. Calibration curves of DDx-PRS and Marginal-PRS for different prior probabilities in simulations. Panel M.** This Figure is analogue to main Figure 2 and reports results for different assumed and true prior probabilities; this Panel reports true.prior5_assume.prior1 (see below). Every Figure panel reports the true diagnosis probability vs. the predicted diagnosis probability for the comparisons of schizophrenia (SCZ) vs. rest, bipolar disorder (BIP) vs. rest, major depressive disorder (MDD) vs. rest, and control vs. rest, for two methods: Marginal-PRS (red) and DDx-PRS (blue). Results are based on concatenated data across 50 simulation replicates. The calibration curves are plotted using a locally estimated scatterplot smoothing (loess)-based smoothing function. The Integrated Calibration Index (ICI) equals the average absolute difference between the calibration curve and the line y=x (plotted in black), weighted by the density of the predicted diagnosis probabilities (displayed in the shaded histograms and in the color-intensity of the calibration curves). AUC, area under the ROC curve. The priors are:

prior1: SCZ=25%, BIP=25%, MDD=25%, control=25%
prior2: SCZ=40%, BIP=20%, MDD=20%, control=20%
prior3: SCZ=20%, BIP=40%, MDD=20%, control=20%
prior4: SCZ=20%, BIP=20%, MDD=40%, control=20%
prior5: SCZ=20%, BIP=20%, MDD=20%, control=40%

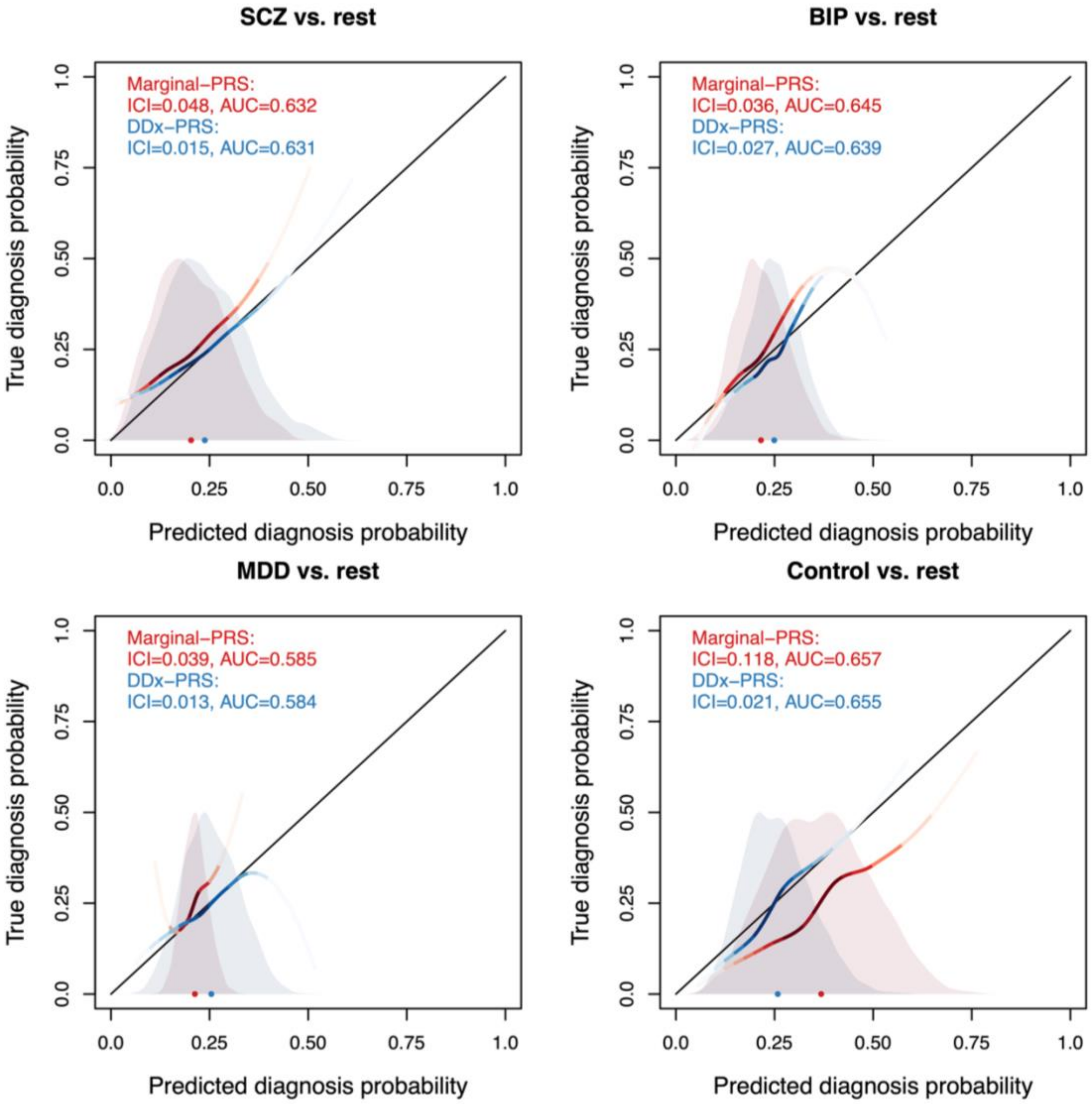

**Figure S7. Calibration and accuracy of DDx-PRS and Marginal-PRS in individual cohorts in PGC data. Panel A – cohort GER.** This Figure is analogue to main Figure 4 and reports the calibration curves for the four PGC cohorts. Every panel reports the true diagnosis probability vs. the predicted diagnosis probability for the comparisons of schizophrenia (SCZ) vs. rest, bipolar disorder (BIP) vs. rest, major depressive disorder (MDD) vs. rest, and control vs. rest, for two methods: Marginal-PRS (red) and DDx-PRS (blue). The calibration curves are plotted using a locally estimated scatterplot smoothing (loess)-based smoothing function. Results are based on the PGC test cohort GER (N=3,368, with 842 for each of SCZ, BIP, MDD and control). The Integrated Calibration Index (ICI) equals the average absolute difference between the calibration curve and the line y=x (plotted in black), weighted by the density of the predicted diagnosis probabilities (displayed in the shaded histograms and in the color-intensity of the calibration curves). AUC, area under the ROC curve. These secondary analyses show that similar results were observed for all four cohorts individually as for the mean results in Figure 3.

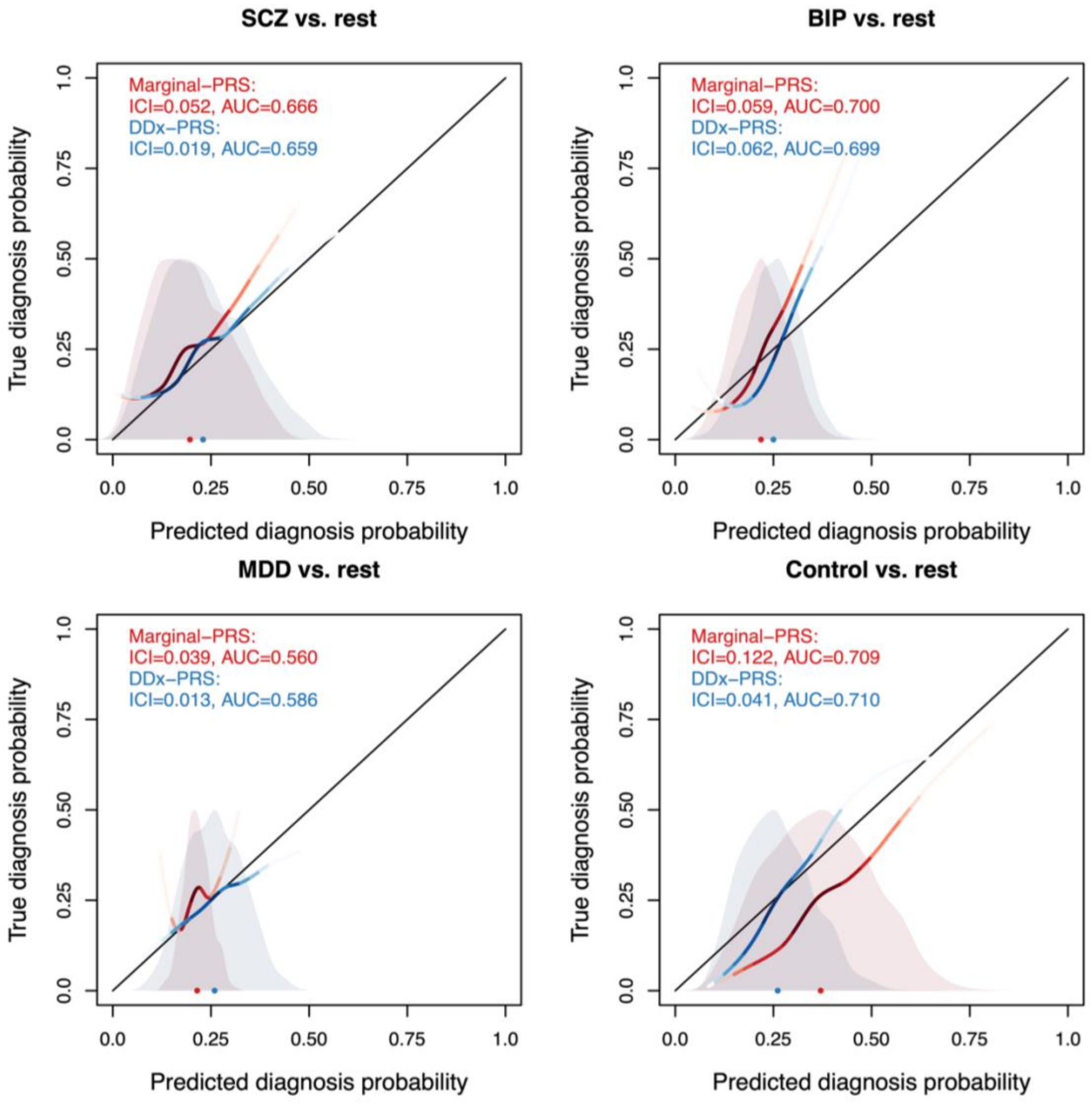

**Figure S7. Calibration and accuracy of DDx-PRS and Marginal-PRS in individual cohorts in PGC data. Panel B – cohort UK1.** This Figure is analogue to main Figure 4 and reports the calibration curves for the four PGC cohorts. Every panel reports the true diagnosis probability vs. the predicted diagnosis probability for the comparisons of schizophrenia (SCZ) vs. rest, bipolar disorder (BIP) vs. rest, major depressive disorder (MDD) vs. rest, and control vs. rest, for two methods: Marginal-PRS (red) and DDx-PRS (blue). The calibration curves are plotted using a locally estimated scatterplot smoothing (loess)-based smoothing function. Results are based on the PGC test cohort UK1 (N=1,136, with 284 for each of SCZ, BIP, MDD and control). The Integrated Calibration Index (ICI) equals the average absolute difference between the calibration curve and the line y=x (plotted in black), weighted by the density of the predicted diagnosis probabilities (displayed in the shaded histograms and in the color-intensity of the calibration curves). AUC, area under the ROC curve. These secondary analyses show that similar results were observed for all four cohorts individually as for the mean results in Figure 3.

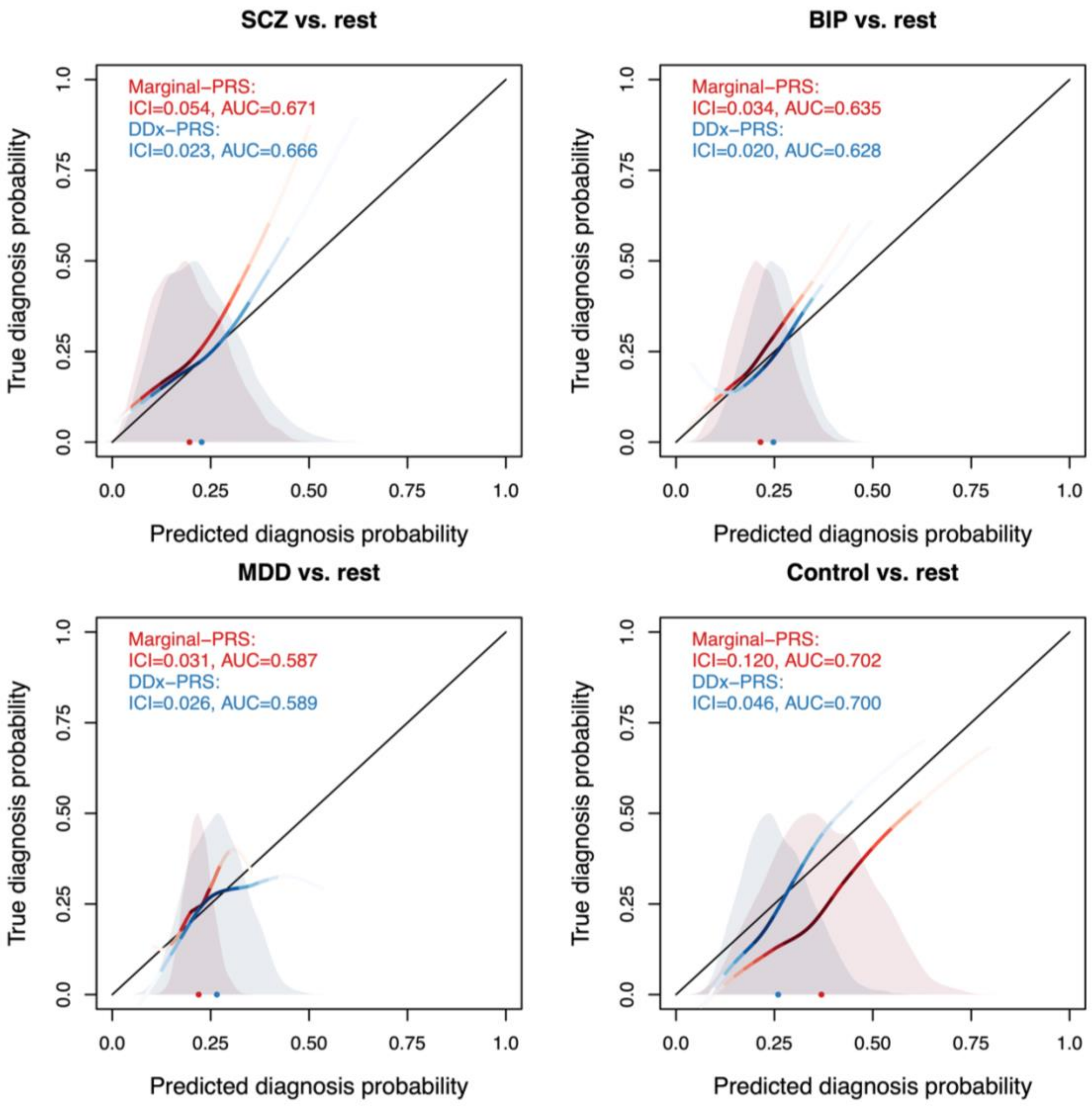

**Figure S7. Calibration and accuracy of DDx-PRS and Marginal-PRS in individual cohorts in PGC data. Panel C – UK2.** This Figure is analogue to main Figure 4 and reports the calibration curves for the four PGC cohorts. Every panel reports the true diagnosis probability vs. the predicted diagnosis probability for the comparisons of schizophrenia (SCZ) vs. rest, bipolar disorder (BIP) vs. rest, major depressive disorder (MDD) vs. rest, and control vs. rest, for two methods: Marginal-PRS (red) and DDx-PRS (blue). The calibration curves are plotted using a locally estimated scatterplot smoothing (loess)-based smoothing function. Results are based on the PGC test cohort UK2 (N=6,080, with 520 for each of SCZ, BIP, MDD and control). The Integrated Calibration Index (ICI) equals the average absolute difference between the calibration curve and the line y=x (plotted in black), weighted by the density of the predicted diagnosis probabilities (displayed in the shaded histograms and in the color-intensity of the calibration curves). AUC, area under the ROC curve. These secondary analyses show that similar results were observed for all four cohorts individually as for the mean results in Figure 3.

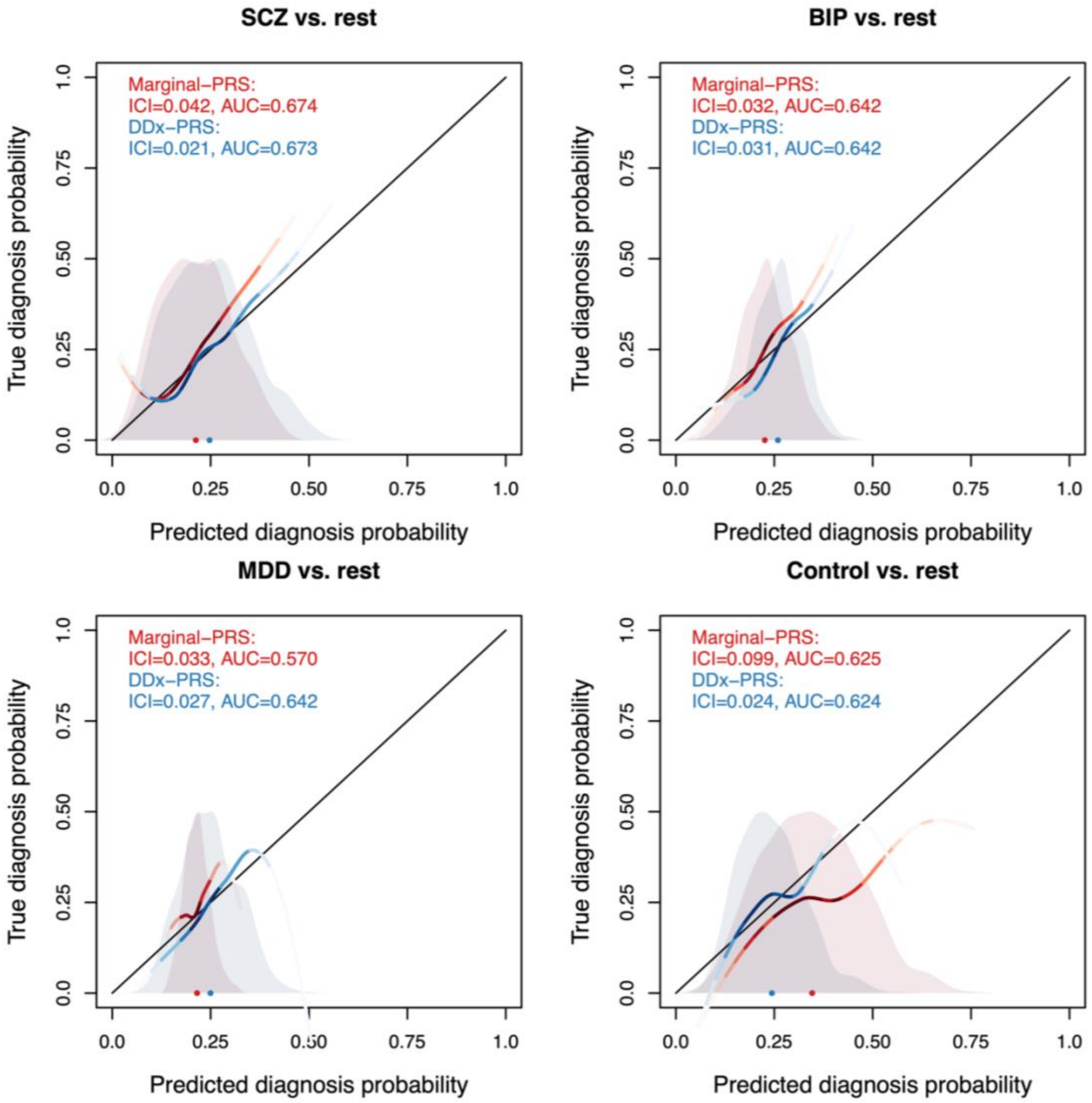

**Figure S7. Calibration and accuracy of DDx-PRS and Marginal-PRS in individual cohorts in PGC data. Panel D – USA.** This Figure is analogue to main Figure 4 and reports the calibration curves for the four PGC cohorts. Every panel reports the true diagnosis probability vs. the predicted diagnosis probability for the comparisons of schizophrenia (SCZ) vs. rest, bipolar disorder (BIP) vs. rest, major depressive disorder (MDD) vs. rest, and control vs. rest, for two methods: Marginal-PRS (red) and DDx-PRS (blue). The calibration curves are plotted using a locally estimated scatterplot smoothing (loess)-based smoothing function. Results are based on the PGC test cohort USA (N=876, with 219 for each of SCZ, BIP, MDD and control). The Integrated Calibration Index (ICI) equals the average absolute difference between the calibration curve and the line y=x (plotted in black), weighted by the density of the predicted diagnosis probabilities (displayed in the shaded histograms and in the color-intensity of the calibration curves). AUC, area under the ROC curve. These secondary analyses show that similar results were observed for all four cohorts individually as for the mean results in Figure 3.

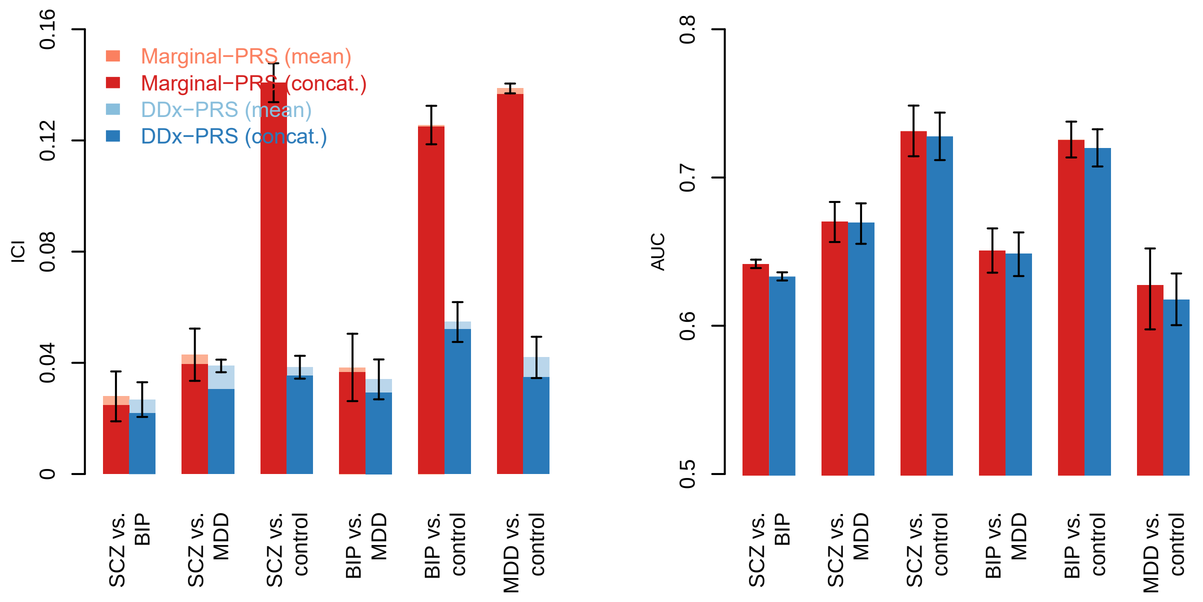

**Figure S8. Calibration and accuracy of DDx-PRS and Marginal-PRS for comparisons across all 6 pairs of diagnostic categories in PGC data.** This Figure accompanies main Figure 3 and reports the calibration (Integrated Calibration Index; ICI) and accuracy (area under the ROC curve; AUC) for the comparisons of schizophrenia (SCZ) vs. bipolar disorder (BIP), SCZ vs. major depressive disorder (MDD), SCZ vs. control, BIP vs. MDD, BIP vs. control and MDD vs. control, for two methods: Marginal-PRS (red) and DDx-PRS (blue). The training data consisted of case-control GWAS summary statistics for SCZ, BIP and MDD; the test data consisted of independent PGC samples subdivided in four cohorts that were matched with respect to country and genotyping platform (total N=11,460, with 2,865 for each of SCZ, BIP, MDD and control). The mean values across these four test cohorts are displayed in light red and light blue, and results based on data concatenated across these four test cohorts are displayed in dark red and dark blue. Error bars denote standard errors. Numerical results are reported in Table S12. These secondary analyses show DDx-PRS yielded better calibration than Marginal-PRS and similar prediction accuracy across all 6 pairs of diagnostic categories, in line with the 4 comparisons of one diagnostic category vs. rest displayed in main Figure 3.

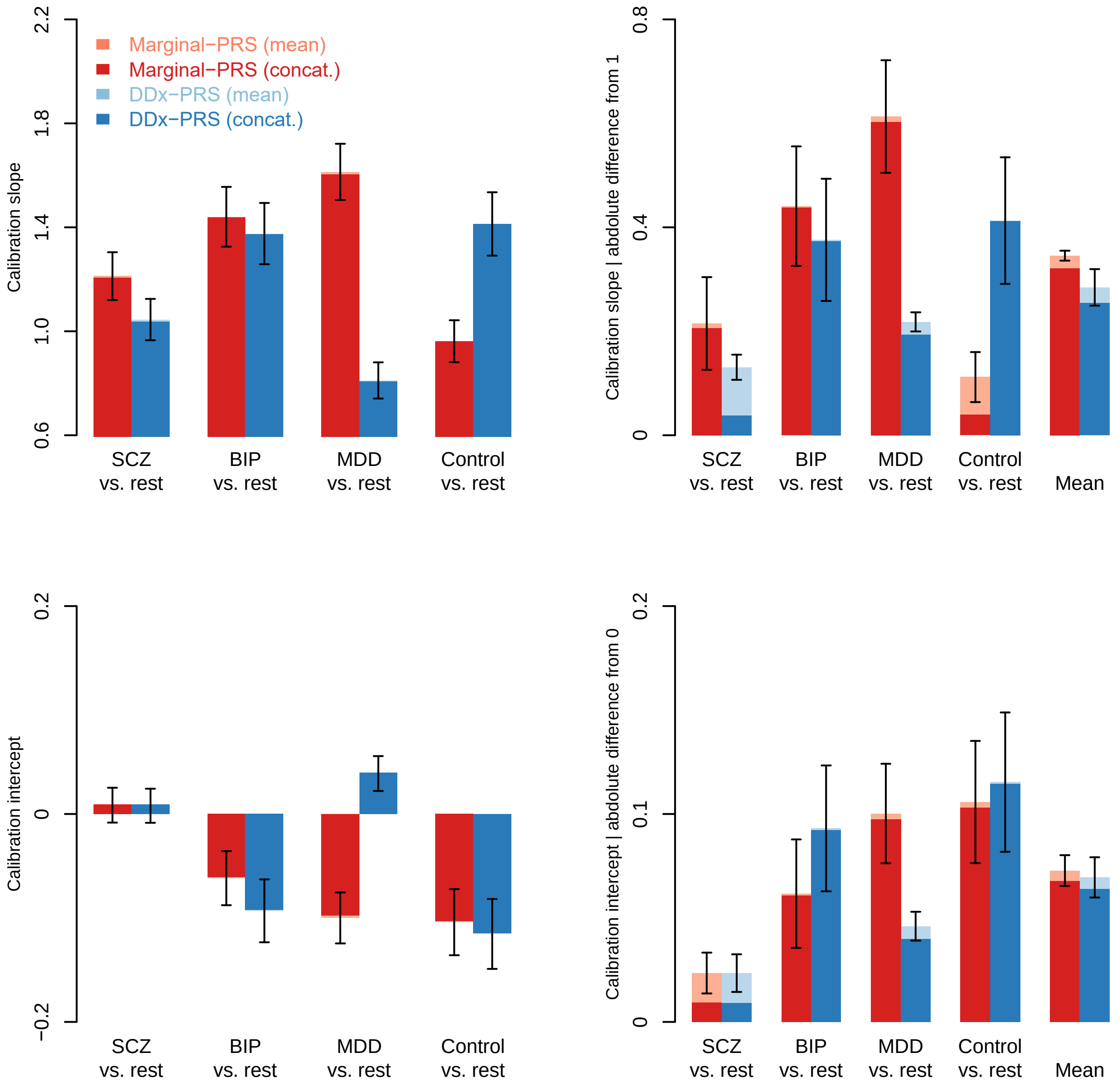

**Figure S9. Calibration slopes of DDx-PRS and Marginal-PRS in PGC data.** This Figure accompanies main Figure 3 and reports the calibration slopes and intercepts for the comparisons of schizophrenia (SCZ) vs. rest, bipolar disorder (BIP) vs. rest, major depressive disorder (MDD) vs. rest, and control vs. rest, for two methods: Marginal-PRS (red) and DDx-PRS (blue). The training data consisted of case-control GWAS summary statistics for SCZ, BIP and MDD; the test data consisted of independent PGC samples subdivided in four cohorts that were matched with respect to country and genotyping platform (total N=11,460, with 2,865 for each of SCZ, BIP, MDD and control). The mean values across these four test cohorts are displayed in light red and light blue, and results based on data concatenated across these four test cohorts are displayed in dark red and dark blue. Error bars denote standard errors. Perfect calibration would attain a slope of 1 and intercept of 0. Numerical results are reported in Table S13. These secondary analyses show that DDx-PRS attained slightly better calibration than Marginal-PRS when considering the calibration slope and intercept instead of the ICI (main Figure 3). We note that the calibration slope shows more variance between comparisons than the ICI, because the slope is impacted by sparse subregions with poor calibration while the ICI is much more robust to this (also see Austin et al. 2019 Statistics in Medicine).

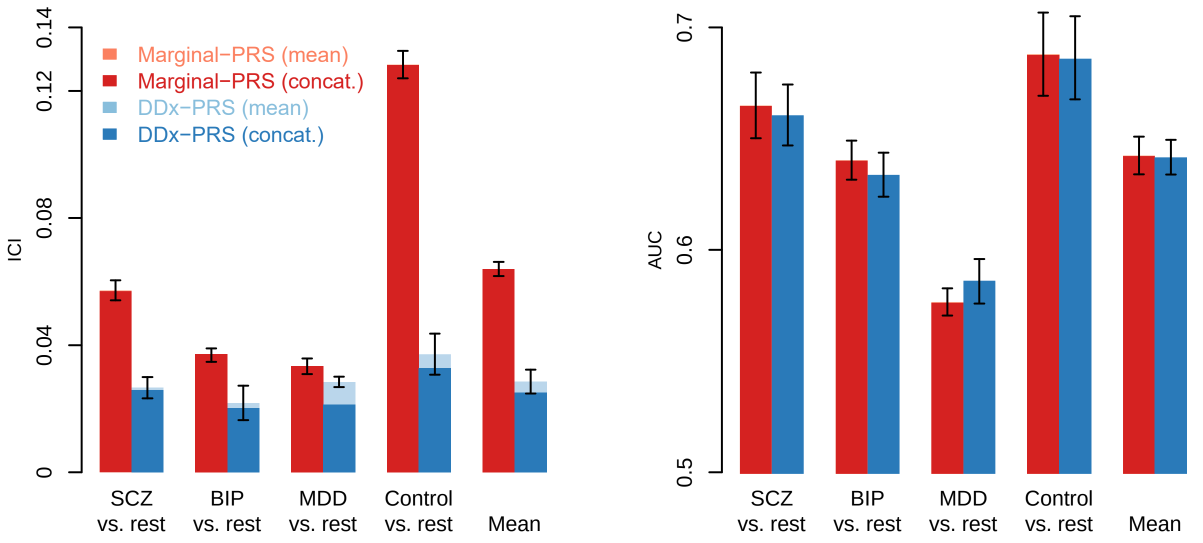

**Figure S10. Calibration and accuracy of DDx-PRS and Marginal-PRS for different QC in PGC data. Panel A – looser SNP QC.** This Figure is analogue to main Figure 3 with SNP QC of INFO>0.3 and MAF>0.01 (compared to INFO>0.9 and MAF>0.10 displayed in Figure 3). We report the calibration (Integrated Calibration Index; ICI) and accuracy (area under the ROC curve; AUC) for the comparisons of schizophrenia (SCZ) vs. rest, bipolar disorder (BIP) vs. rest, major depressive disorder (MDD) vs. rest, control vs. rest and the mean across these four comparisons, for two methods: Marginal-PRS (red) and DDx-PRS (blue). The training data consisted of case-control GWAS summary statistics for SCZ, BIP and MDD; the test data consisted of independent PGC samples subdivided in four cohorts that were matched with respect to country and genotyping platform (total N=11,460, with 2,865 for each of SCZ, BIP, MDD and control). The mean values across these four test cohorts are displayed in light red and light blue, and results based on data concatenated across these four test cohorts are displayed in dark red and dark blue. Error bars denote standard errors. Numerical results are reported in Table S14. These secondary analyses show that arbitrary choices in QC had no impact on the presented results.

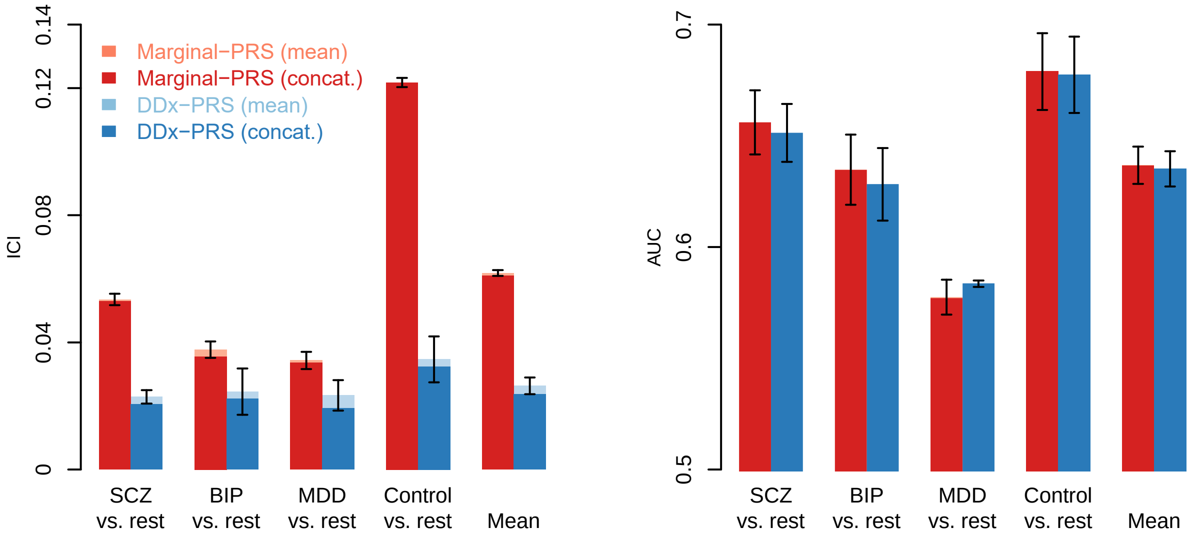

**Figure S10. Calibration and accuracy of DDx-PRS and Marginal-PRS for different QC in PGC data. Panel B – stricter subject QC.** This Figure is analogue to main Figure 3 with additional stricter subject QC (see Methods). We report the calibration (Integrated Calibration Index; ICI) and accuracy (area under the ROC curve; AUC) for the comparisons of schizophrenia (SCZ) vs. rest, bipolar disorder (BIP) vs. rest, major depressive disorder (MDD) vs. rest, control vs. rest and the mean across these four comparisons, for two methods: Marginal-PRS (red) and DDx-PRS (blue). The training data consisted of case-control GWAS summary statistics for SCZ, BIP and MDD; the test data consisted of independent PGC samples subdivided in four cohorts that were matched with respect to country and genotyping platform (total N=10,952, with 2,738 for each of SCZ, BIP, MDD and control; total N main analyses = 11,460). The mean values across these four test cohorts are displayed in light red and light blue, and results based on data concatenated across these four test cohorts are displayed in dark red and dark blue. Error bars denote standard errors. Numerical results are reported in Table S14. These secondary analyses show that arbitrary choices in QC had no impact on the presented results.

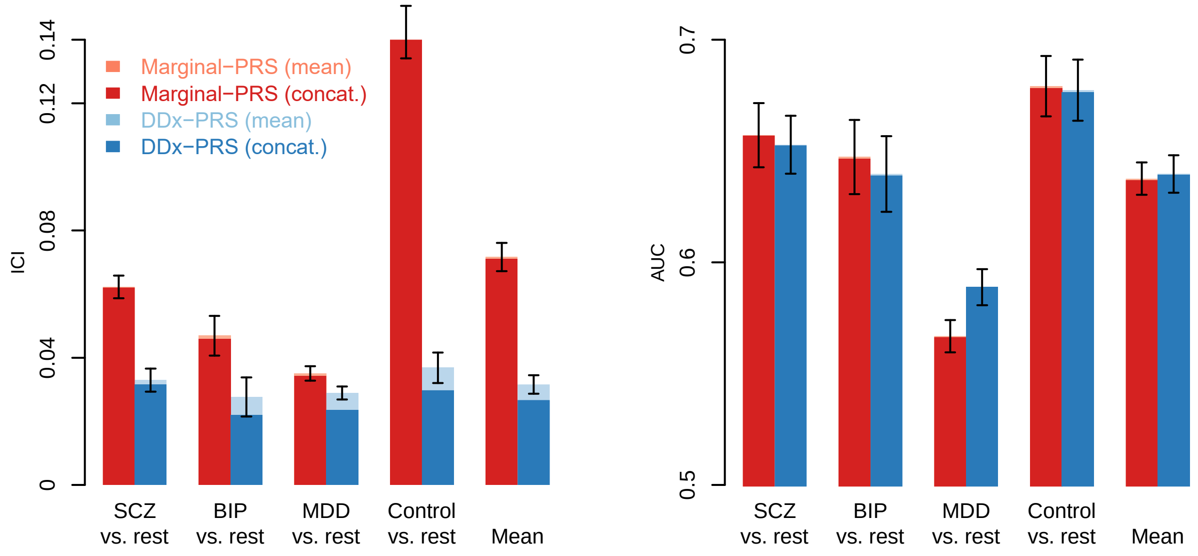

**Figure S10. Calibration and accuracy of DDx-PRS and Marginal-PRS for different QC in PGC data. Panel C – looser SNP QC and stricter subject QC.** This Figure is analogue to main Figure 3 with SNP QC of INFO>0.3 and MAF>0.01 (compared to INFO>0.9 and MAF>0.10 displayed in Figure 3) and additional stricter subject QC (see Methods). We report the calibration (Integrated Calibration Index; ICI) and accuracy (area under the ROC curve; AUC) for the comparisons of schizophrenia (SCZ) vs. rest, bipolar disorder (BIP) vs. rest, major depressive disorder (MDD) vs. rest, control vs. rest and the mean across these four comparisons, for two methods: Marginal-PRS (red) and DDx-PRS (blue). The training data consisted of case-control GWAS summary statistics for SCZ, BIP and MDD; the test data consisted of independent PGC samples subdivided in four cohorts that were matched with respect to country and genotyping platform (total N=10,952, with 2,738 for each of SCZ, BIP, MDD and control; total N main analyses = 11,460). The mean values across these four test cohorts are displayed in light red and light blue, and results based on data concatenated across these four test cohorts are displayed in dark red and dark blue. Error bars denote standard errors. Numerical results are reported in Table S14. These secondary analyses show that arbitrary choices in QC had no impact on the presented results.

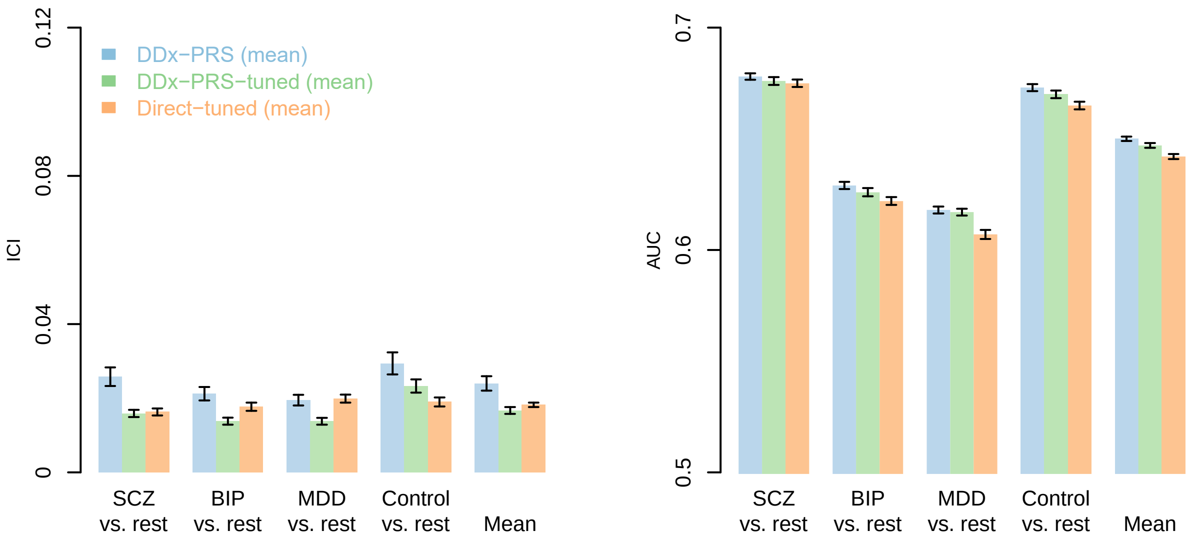

**Figure S11. Calibration and accuracy of DDx-PRS, DDx-PRS-tuned and Direct-tuned in simulations.** We report the calibration (Integrated Calibration Index; ICI) and accuracy (area under the ROC curve; AUC) for the comparisons of schizophrenia (SCZ) vs. rest, bipolar disorder (BIP) vs. rest, major depressive disorder (MDD) vs. rest, control vs. rest and the mean across these four comparisons, for three methods: DDx-PRS (blue), DDx-PRS-tuned (green) and Direct-tuned (orange). Results are based on 50 simulation replicates; mean values are displayed in light red and light blue. Simulated tuning data sets comprised of N=200 of each of SCZ, BIP, MDD and control; other simulation parameters are the same as in the main simulations reported in Figure 1. Error bars denote standard errors. Numerical results are reported in Table S16. These secondary analyses show that DDx-PRS, DD-PRS-tuned and Direct-tuned yielded similar performance in simulations, which is in line with the results based on PGC data presented in main Figure 5.

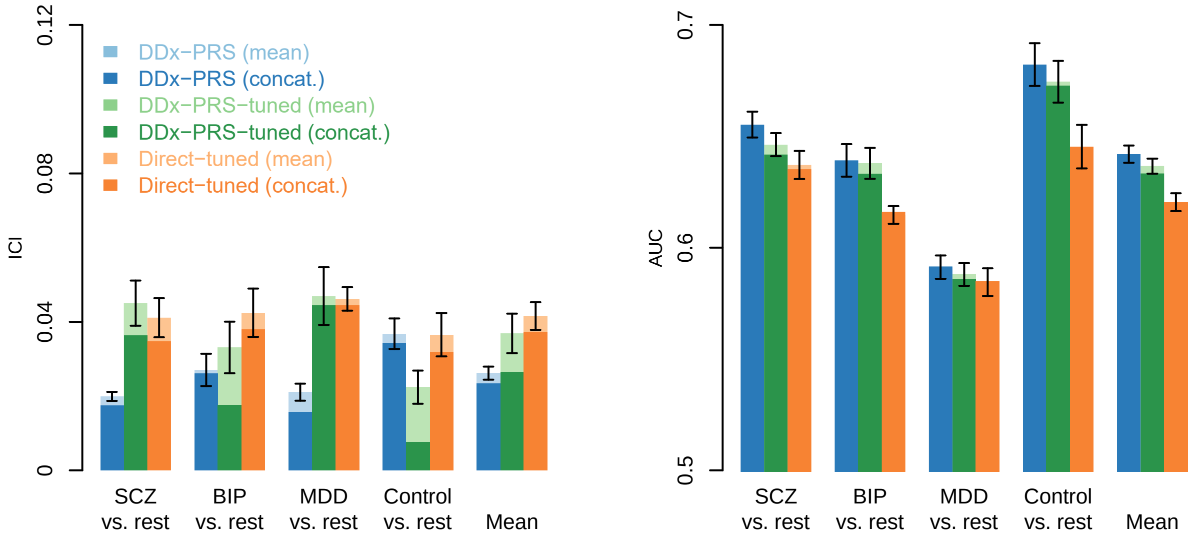

**Figure S12. Calibration and accuracy of DDx-PRS-tuned and Direct-tuned at smaller tuning sample sizes in PGC data. Panel A – N tuning 50 for each of SCZ, BIP, MDD control.** This Figure is analogue to main Figure 5 (which reports results for N tuning 200 for each of SCZ, BIP, MDD control) and reports the calibration (Integrated Calibration Index; ICI) and accuracy (area under the ROC curve; AUC) for the comparisons of schizophrenia (SCZ) vs. rest, bipolar disorder (BIP) vs. rest, major depressive disorder (MDD) vs. rest, control vs. rest and the mean across these four comparisons, for three methods: DDx-PRS (blue), DDx-PRS-tuned (green) and Direct-tuned (orange). The training data consisted of case-control GWAS summary statistics for SCZ, BIP and MDD; the test data consisted of independent PGC samples subdivided in four cohorts that were matched with respect to country and genotyping platform (total N=11,460, with 2,865 for each of SCZ, BIP, MDD and control). For tuning for DDx-PRS-tuned and Direct-tuned, 4x3=12 tuning/test cohort-pairs were analysed with tuning data restricted to 50 samples from each of SCZ, BIP, MDD and control (total $N_{tuning}$=200). The mean values across these four test cohorts (DDx-PRS) resp. 12 tuning/test cohort-pairs (DDx-PRS-tuned and Direct-tuned) are displayed in light blue, light green and light orange, and results based on data concatenated across these four test cohorts resp. 12 tuning/test cohort-pairs are displayed in dark blue, dark green and dark orange. Error bars denote standard errors. Numerical results are reported in Table S17. These secondary analyses show that the results were only slightly impacted by smaller tuning sample size, compared to Figure 5.

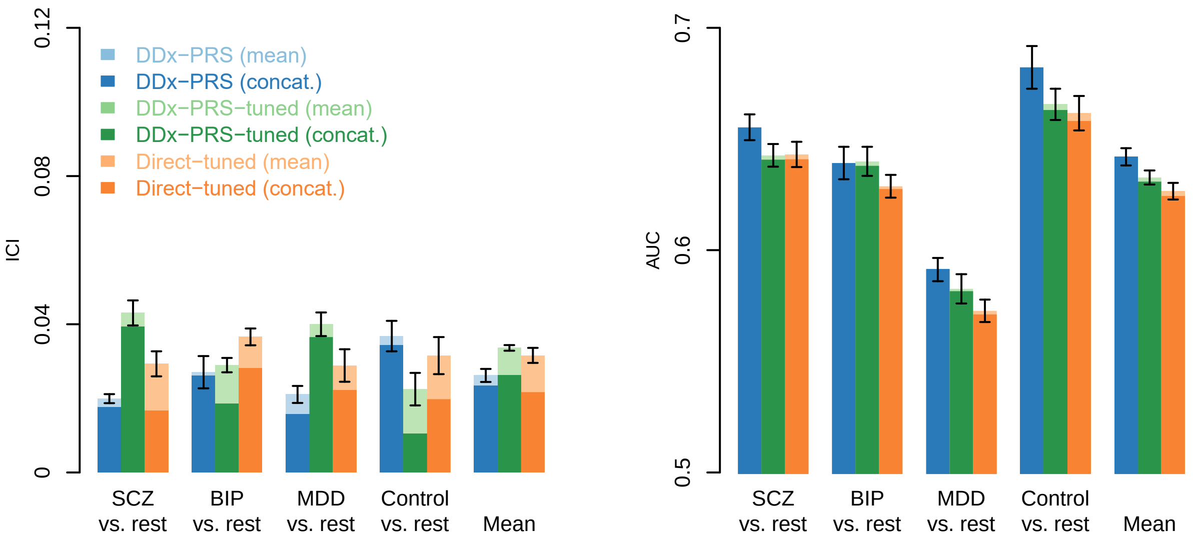

**Figure S12. Calibration and accuracy of DDx-PRS-tuned and Direct-tuned at smaller tuning sample sizes in PGC data. Panel B – N tuning 100 for each of SCZ, BIP, MDD control.** This Figure is analogue to main Figure 5 (which reports results for N tuning 200 for each of SCZ, BIP, MDD control) and reports the calibration (Integrated Calibration Index; ICI) and accuracy (area under the ROC curve; AUC) for the comparisons of schizophrenia (SCZ) vs. rest, bipolar disorder (BIP) vs. rest, major depressive disorder (MDD) vs. rest, control vs. rest and the mean across these four comparisons, for three methods: DDx-PRS (blue), DDx-PRS-tuned (green) and Direct-tuned (orange). The training data consisted of case-control GWAS summary statistics for SCZ, BIP and MDD; the test data consisted of independent PGC samples subdivided in four cohorts that were matched with respect to country and genotyping platform (total N=11,460, with 2,865 for each of SCZ, BIP, MDD and control). For tuning for DDx-PRS-tuned and Direct-tuned, 4x3=12 tuning/test cohort-pairs were analysed with tuning data restricted to 10 samples from each of SCZ, BIP, MDD and control (total $N_{tuning}$=400). The mean values across these four test cohorts (DDx-PRS) resp. 12 tuning/test cohort-pairs (DDx-PRS-tuned and Direct-tuned) are displayed in light blue, light green and light orange, and results based on data concatenated across these four test cohorts resp. 12 tuning/test cohort-pairs are displayed in dark blue, dark green and dark orange. Error bars denote standard errors. Numerical results are reported in Table S17. These secondary analyses show that the results were only slightly impacted by smaller tuning sample size, compared to Figure 5.

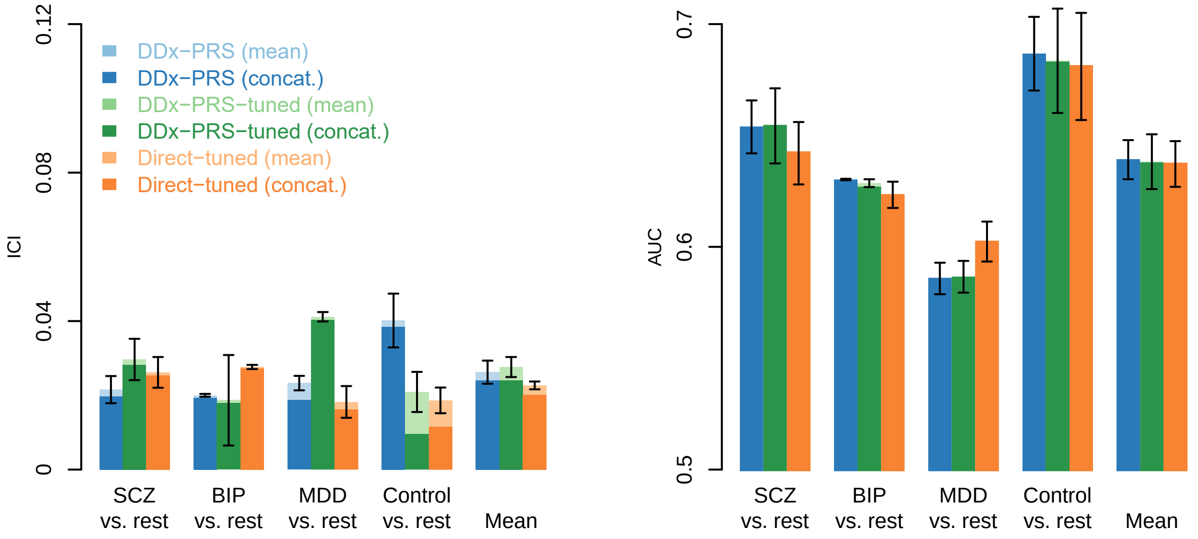

**Figure S13. Calibration and accuracy of DDx-PRS-tuned and Direct-tuned using within-cohort tuning data in PGC data.** This Figure is analogue to main Figure 5 and based on within-cohort tuning (compared to cross-cohort tuning reported in main Figure 5). We report the calibration (Integrated Calibration Index; ICI) and accuracy (area under the ROC curve; AUC) for the comparisons of schizophrenia (SCZ) vs. rest, bipolar disorder (BIP) vs. rest, major depressive disorder (MDD) vs. rest, control vs. rest and the mean across these four comparisons, for three methods: DDx-PRS (blue), DDx-PRS-tuned (green) and Direct-tuned (orange). The training data consisted of case-control GWAS summary statistics for SCZ, BIP and MDD; the test data consisted of independent PGC samples subdivided in two cohorts that were matched with respect to country and genotyping platform (GER (N=3,368) and UK2 (N=6,080); sample-sizes of UK1 and USA were too small for within-cohort tuning). For tuning for DDx-PRS-tuned and Direct-tuned, tuning data were randomly selected at 200 samples from each of SCZ, BIP, MDD and control (total $N_{tuning}$=800); the test data comprised of the remaining samples (i.e. 2,568 for GER and 5,280 for UK2). The mean values across these two test cohorts are displayed in light blue, light green and light orange, and results based on data concatenated across these two test cohorts are displayed in dark blue, dark green and dark orange. Error bars denote standard errors. Numerical results are reported in Table S18. These secondary analyses show that the performance of DDx-PRS-tuned and Direct-tuned improved only very slightly when using within-cohort tuning instead of cross-cohort tuning (main Figure 5), and that results for DDx-PRS, DDx-PRS-tuned and Direct-tuned remained comparable.

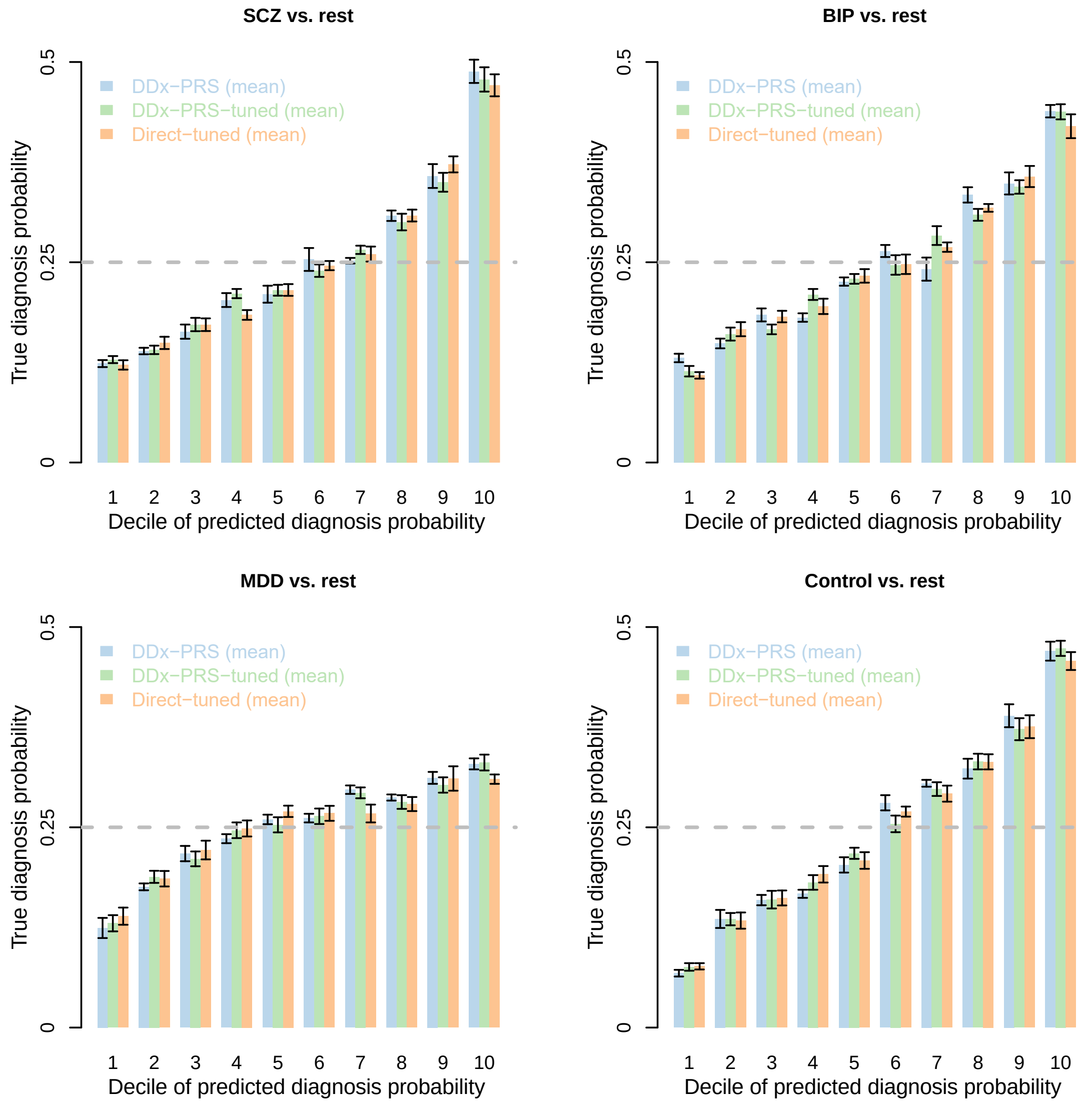

**Figure S14. True diagnosis probability per decile of predicted diagnosis probability of DDx-PRS, DDx-PRS-tuned and Direct-tuned in PGC data.** This Figure accompanies main Figure 7 and reports the true diagnosis probability per decile of predicted diagnosis probability for the comparisons of schizophrenia (SCZ) vs. rest, bipolar disorder (BIP) vs. rest, major depressive disorder (MDD) vs. rest, and control vs. rest, for three method: DDx-PRS (blue), DDx-PRS-tuned (green) and Direct-tuned (orange). The training data consisted of case-control GWAS summary statistics for SCZ, BIP and MDD; the test data consisted of independent PGC samples subdivided in four cohorts that were matched with respect to country and genotyping platform (total N=11,460, with 2,865 for each of SCZ, BIP, MDD and control). For tuning for DDx-PRS-tuned and Direct-tuned, 4x3=12 tuning/test cohort-pairs were analysed with tuning data restricted to 200 samples from each of SCZ, BIP, MDD and control (total $N_{\text{tuning}}$=800). The mean values across these four test cohorts (DDx-PRS) resp. 12 tuning/test cohort-pairs (DDx-PRS-tuned and Direct-tuned) are displayed in light blue, light green and light orange. Error bars denote standard errors. Numerical results are reported in Table S21. These secondary analyses show that the results are similar for DDx-PRS, DDx-PRS-tuned and Direct-tuned.

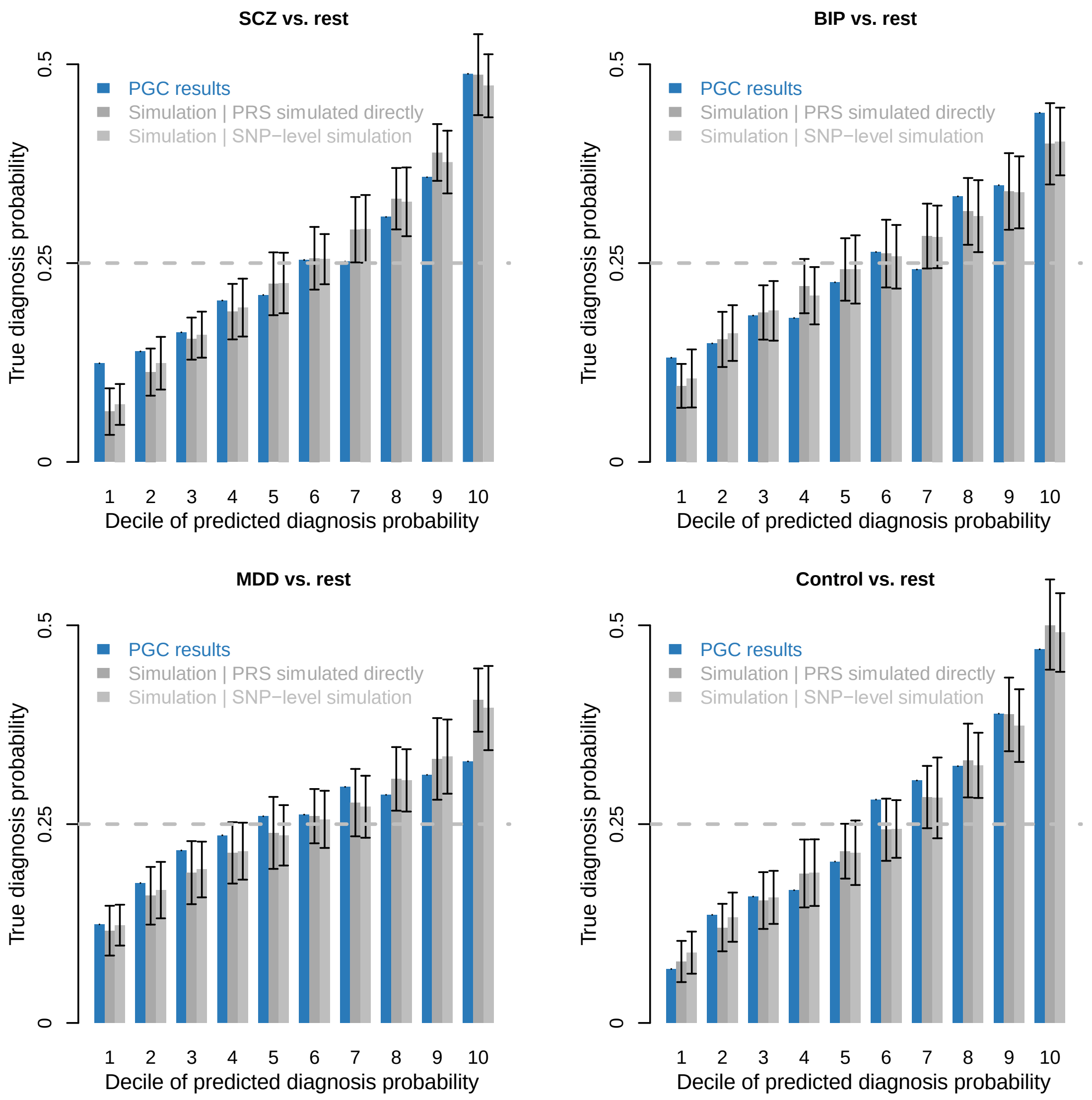

**Figure S15. True diagnosis probability per decile of predicted diagnosis probability of DDx-PRS in PGC data and in simulations at the exact empirical values.** We report the true diagnosis probability per decile of predicted diagnosis probability for the comparisons of schizophrenia (SCZ) vs. rest, bipolar disorder (BIP) vs. rest, major depressive disorder (MDD) vs. rest, and control vs. rest, for one method: DDx-PRS. Empirical results in PGC data are reported in blue, and present the results based on concatenated data (see Figure S14). Results based on simulating PRS directly are displayed in dark grey and results based on simulating SNP-level data are reported in light grey; error bars of simulation denote 1.96 times the SE across 50 simulation runs. Parameters of simulations are set to the exact empirical values of % of SNP-heritability captured by case-control PRS (i.e., 42% for SCZ, 47% for BIP, and 44% for MDD); all other parameters are the same as in the main simulations. Numerical results are reported in Table S22. These secondary analyses show that simulation results were close to the empirical results based on PGC data, which suggests that the simulations can be used to approximate projections of clinical utility at larger training sample sizes.
